# Improving epidemic nowcasting with auxiliary information and geographic networks

**DOI:** 10.64898/2026.11.21.26368654

**Authors:** Ruiqi Lyu, Alistair Turcan, Bryan Wilder

## Abstract

Monitoring infectious disease trends requires timely information on disease activity, but direct measurements suffer reporting delays, hindering public health response. Numerous less-direct measurements are collected rapidly, although their relationship to disease can change with behavior, reporting, and data availability. Here, we present GLCast, a statistical nowcasting framework that effectively uses auxiliary information to estimate delayed surveillance measurements, without requiring any verification beforehand that these data will improve performance. At each retraining window, GLCast automatically selects useful data streams through adaptive local regression and further learns dynamic epidemic neighbor graphs to aggregate models across. We evaluate GLCast on six years of weekly influenza-like illness with retail sales, three years of daily COVID-19 hospital admissions with search and survey signals, and seven recent influenza, COVID-19, and RSV surveillance targets with environmental, travel and online-attention streams. Against methods without auxiliary information, GLCast lowers mean prediction error by 25.1% for historical influenza-like illness and 12.9% for historical COVID-19 admissions against the strongest baselines, improving 45 of 45 and 49 of 51 evaluated jurisdictions, respectively, with further performance gains in critical upswing periods. In the seven modern targets, GLCast achieves the lowest overall error and maintains strong performance when multiple diseases experience overlapping upswings. Interpreting how GLCast uses additional information, we find that the features and neighbors selected are indeed dynamic over time and heterogeneous across different states. Interestingly, outbreaks appear to be change points where both feature and neighbor importance will dramatically shift. GLCast tends to assign new importance to auxiliary information at the peak of an outbreak, disregards neighbors during outbreaks, and heavily relies on neighbors during down periods.

**Significance:** Public health response depends on timely measures of disease activity, but direct surveillance reports often arrive late. Retail purchases, symptom searches, surveys, weather, and online attention offer earlier information whose usefulness changes across locations and epidemic phases. We introduce GLCast, a framework that repeatedly selects useful streams and learns which regional models can improve local estimates. Across nine applications, it improves nowcasting while revealing changing use of auxiliary information and geographic borrowing. Outbreaks are associated with shifts in stream importance, cross-state information is useful during down periods, and less useful during upswings. These interpretable outputs can help researchers identify promising surveillance signals and regional relationships.

## 1. Introduction

Public health response depends on accurate monitoring of disease activity, which is typically measured directly through case counts, hospitalizations, and deaths (1–3). These direct measures, however, face hurdles at every stage of the testing, clinical, laboratory, and reporting pipeline, and the resulting delays are often days to weeks, leaving decision-makers working from incomplete measurements (4–7). During the COVID-19 emergency, for instance, U.S. hospitals were required to report hospitalization and capacity data to the Department of Health and Human Services at high frequency, and CMS introduced conditions of participation and monetary penalties to enforce compliance, a move strongly challenged by the American Hospital Association (8, 9). Such enforcement is difficult to sustain, and most circulating viruses, including influenza, RSV, and SARS-CoV-2 today, do not have the same attention as COVID-19 did during the emergency phase and report cases with less frequency. This inherent imperfection of surveillance data necessitates nowcasting: estimating current disease activity from incomplete measurements, in contrast to forecasting, which predicts future disease activity (10, 11).

Previous work has often performed nowcasting using the history and provisional observations of the target data stream to predict current counts (5, 12–14). These methods model reporting delays, backfill, or onset-to-report distributions in the target stream itself. A second line of work estimates transmission intensity directly, through reproduction numbers and growth rates (15– 17), or combines deconvolution of delayed observations with sensor fusion across multiple indicators (18–20). A complementary literature integrates auxiliary data streams, including medical claims, symptom surveys, search trends, mobility, wastewater, and wearables, to improve real-time tracking, forecasting, hotspot prediction, or outbreak detection (4, 21–28). Search and other internet-derived signals in particular have been used extensively for influenza nowcasting and for real-time estimation of emerging outbreaks (29–38). Closest to our setting, Shen et al. used versioned medical-claims data to nowcast reported COVID-19 hospitalizations from two disease-specific claims indicators and selected temporal lags (39). Several influential approaches thus use a small set of engineered indicators or explicitly screen candidate predictors before fitting (30, 33, 39). Related multistream approaches have also combined health and behavioral signals with spatial or epidemic-network structure for early warning, outbreak detection, and nowcasting (33, 40, 41). Because epidemic activity is coupled across locations by mobility and other spatial interactions, geographic network structure can provide complementary information beyond local signals.

A less explored direction is how to use large collections of auxiliary data streams to support nowcasting when the target surveillance data are delayed, without requiring manual curation or preprocessing to identify useful data streams. This is particularly challenging as only some auxiliary streams are expected to be useful for a given pathogen, location, or time period, and their usefulness may change as behavior, testing, reporting practices, or data availability shift. Digital surveillance systems based on search behavior, for instance, degraded when the relationship between search activity and disease incidence changed over time (42, 43); care-seeking and healthcare delivery shifted abruptly during the COVID-19 pandemic (44); public attitudes and behavior moved rapidly as well (45, 46); and transmission itself responds to interventions, behavior, and population susceptibility in ways that vary over time and place (47–49). Integrating these data therefore requires dynamically filtering and weighting sources according to their current predictive value.

To effectively leverage these data for nowcasting, we introduce GLCast, a statistical nowcasting framework that estimates disease activity from a library of auxiliary streams. At each training window, GLCast re-selects informative data streams without needing to specify their usefulness beforehand. GLCast then extends this philosophy to the regional level, dynamically learning useful regions with similar epidemic dynamics to aggregate models across and further improve nowcasting estimates. We evaluate GLCast on six years of weekly influenza-like illness with retail sales, three years of daily COVID-19 hospital admissions with search and survey signals, and seven recent influenza, COVID-19, and RSV surveillance targets with context and online-attention streams. Against methods without auxiliary information, GLCast lowers mean prediction error by 25.1% for historical influenza-like illness and 12.9% for historical COVID-19 admissions against the strongest baselines, improving 45 of 45 and 49 of 51 evaluated jurisdictions, respectively, with further performance gains in critical upswing periods. In the seven modern targets, GLCast achieves the lowest overall error and maintains strong performance when multiple diseases experience overlapping upswings. Interpreting how GLCast uses additional information, we find that the features and neighbors selected are indeed dynamic over time and heterogeneous across different states and diseases. Interestingly, outbreaks appear to be change points where both feature and neighbor importance will dramatically shift. GLCast tends to assign new importance to auxiliary information at the peak of an outbreak, disregards neighbors during outbreaks, and heavily relies on neighbors during down periods.

## Results

### GLCast overview

GLCast is a general statistical framework for estimating a delayed epidemic surveillance stream from its own history, auxiliary data streams, and models fitted in other geographic regions. It consists of three steps (Fig. 1). First, for a given region and time point, GLCast constructs predictors from recent target and auxiliary measurements. Second, it searches this corpus for useful streams with a group-structured regression, treating the lags of each stream as related features. The weight structure allows entire streams and lags to be excluded if they are deemed uninformative. Third, GLCast screens other regions for similar predictive relationships, selects up to five donor models, and blends their predictions with the local model. The selected streams and regional weights change as models are refitted, allowing the frame-work to respond to changes in the observed information environment (42, 50). For full details see Materials and Methods.

**Fig. 1.**
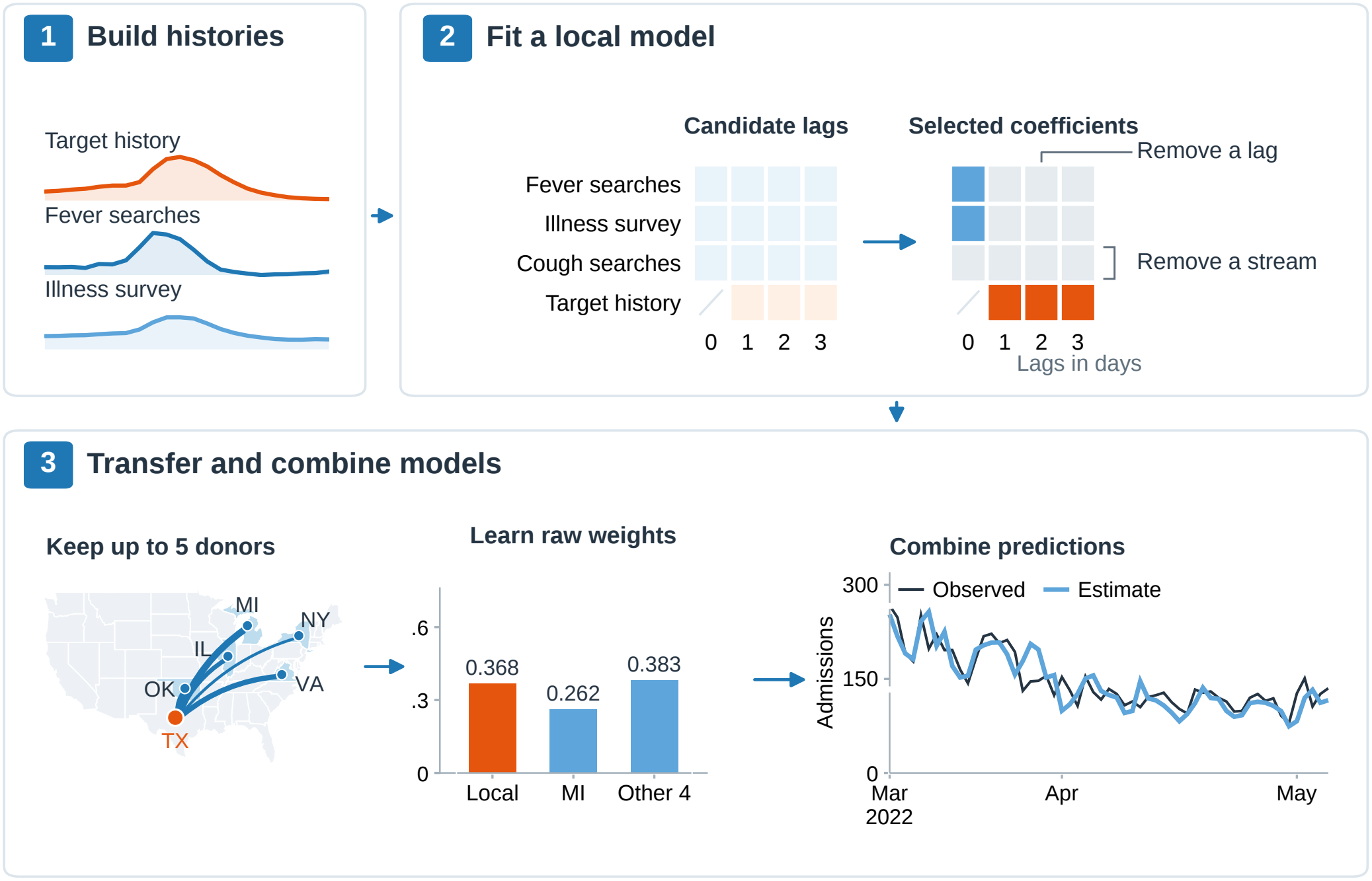
Overview of the GLCast nowcasting framework. (**1**) Target history and auxiliary signals are organized into a library of lagged predictors. The illustrative curves are seven-day means of standardized national COVID-19 series from November 1, 2021, through March 31, 2022. (**2**) Adaptive group penalties select streams, while additional coordinate penalties can remove individual lags. The candidate grid denotes feature availability; the selected grid shows a subset of the actual Texas fit on April 1, 2022. Blue and orange cells retain auxiliary and target-history coefficients, respectively; gray cells have zero coefficients. The daily example uses auxiliary lags 0-3 and target lags 1-3 days. (**3**) Donor models are evaluated on the target region’s features. After screening, up to five are retained by recent local calibration error, and their predictions are combined with the local model using learned nonnegative weights. The map and bars show actual Texas weights at the same origin; line widths increase with raw donor weight, and “Other 4” sums the four smaller donor weights. The daily Texas trace spans March 10-May 5, 2022.

GLCast produces two further outputs beyond pre-dictions themself: local stream-selection and coefficient-importance profiles, which summarize how auxiliary information enters each local fit; and a dynamic network of regional model weights, which summarizes geographic borrowing. This design enables inherent interpretability, allowing epidemiologists to see which data from what states led to individual predictions. It can also be informative for future data collection practices to see what information is particularly useful, and how that information remained useful across time and space.

We apply GLCast in three complementary retrospective settings spanning daily and weekly surveillance, outpatient illness, hospital admissions, and emergency-department visits (Table 1; *SI Appendix*, Table S1). First, we analyze six years of weekly influenza-like illness (ILI) from ILINet in 45 U.S. states (2012–2017), using 31 NielsenIQ retail-sales streams that include cough and cold remedies, thermometers, tissues, analgesics, and hygiene products (51, 52) (*SI Appendix*, Table S2). Second, we analyze three years of daily COVID-19 hospital admissions across the 50 states and the District of Columbia (2021– 2024), using HHS admissions distributed through Delphi COVIDcast together with seven Google symptom-search sets and four Delphi/Facebook COVID-19 Trends and Impact Survey signals (4, 23, 53). These auxiliary streams capture searches for symptoms such as fever, cough, and shortness of breath, alongside survey measures of COVID-like illness, community illness, testing, and test positivity (*SI Appendix*, Table S3). Third, we evaluate seven modern weekly respiratory targets across 51-52 juris-dictions, with target-specific scoring windows during 2023-2026: NHSN hospital admissions and NSSP emergency-department visit percentages for influenza, COVID-19, and RSV, together with ILINet %ILI (51, 54, 55). Each modern target uses eight shared and likely-less powerful context streams: temperature, dew point, precipitation, wildfire-smoke and heavy-smoke days, winter-storm and heat warnings, and TSA passenger throughput, plus disease-specific Wikipedia page views and GDELT news volume (56–61) (*SI Appendix*, Table S4). Across these settings, we examine 56 distinct auxiliary streams and compare GLCast with ten target-only baselines spanning persistence, seasonal, climatology, exponential smoothing, ARIMA, exponentially weighted local autoregression, and pooled or spatial-lag autoregressions (39, 62) (*SI Appendix*, Appendix G). Across all analyses, we refit models weekly, using rolling training windows of 55 weeks for classic ILI and the modern targets and 360 days for classic COVID-19. We assess accuracy overall and during outbreak and non-outbreak periods defined by LRTrend (41), examine coinciding respiratory upswings in the modern panel, and use ablations, sensitivity analyses, and temporal diagnostics to characterize the contributions of auxiliary information and geographic borrowing. Together, these analyses test the framework across distinct diseases, surveillance endpoints, epidemic phases, and information environments.

**Table 1.** Main analysis configurations.

| Setting | Classic ILI | Classic COVID-19 | Modern, seven targets |
|---|---|---|---|
| Target | ILINet %ILI | HHS daily admissions | NHSN admissions; NSSP ED%;<br>ILINet %ILI |
| Time unit | Week | Day | Week |
| Scored jurisdictions | 45 states | 50 states + D.C. | 51 for NSSP; 52 otherwise |
| Scored period | Feb 2012–Dec 2017 | Apr 2021–Apr 2024 | Feb 2023–Aug 2026; RSV<br>admissions Jan 2024–Aug 2026 |
| Scored cells | 13,680 | 55,080 | 5,779–9,301 per target |
| Auxiliary streams | 31 retail categories | 7 Google + 4 CTIS | 8 context + 2 attention per disease |
| Fit/calibration/test | 55/4/4 weeks | 360/30/30 days | 55/4/4 weeks |
| Auxiliary lags | {0} weeks | {0, 1, 2, 3} days | {0} weeks |
| Target-history lags | {2} weeks | {1, 2, 3} days | {2} weeks |
| Primary refit cadence | Weekly (Sunday) | Every 7 days (Friday) | Weekly (Saturday) |
| Primary served offsets | 1 week | 1–7 days | 2 weeks |

### Nowcasting influenza-like illness with retail information

We apply GLCast to weekly state-level ILI across 45 U.S. states, in conjunction with 31 auxiliary signals from state-level retail unit sales (Materials and Methods; Table S2). We evaluate on 13,680 state-weeks from February 19, 2012, through December 10, 2017. We apply LRTrend to retrospectively annotate outbreak periods, which cover 46.5% of scored state-weeks due to influenza’s seasonality.

Fig. 2*a* reports the mean across states of per-state RMSE. GLCast reaches 0.544 percentage points of ILI against 0.726 for the strongest overall comparator, the pooled autoregression with coefficients shared across states, a reduction of 25.1% (paired difference 0.182 percentage points; interval [0.149, 0.225]). The reduction against this same comparator is 30.2% inside outbreaks (difference 0.243; [0.195, 0.308]; Fig. 2*b*) and 18.2% outside them (difference 0.117; [0.092, 0.155]). ETS, a general state-space time series forecasting framework based on exponential smoothing, has the lowest comparator RMSE within outbreaks, where GLCast reduces error by 29.4% (0.561 versus 0.796). The simple Persistence method, which simply predicts the last observation, is within 0.6–3.6% of the best baseline in every stratum, while GLCast is the only method with a large advantage over Persistence, reducing RMSE by 18.7–31.9%. At the individual-state level, GLCast improves on Pooled AR all shared in all 45 states, with a median reduction of 23.9% and reductions ranging from 3.1% in Maine to 44.6% in Indiana. Additional evaluation stratifications across states and non-outbreak periods are shown in *SI Appendix*, Figs. S4 and S5, respectively.

**Fig. 2.**
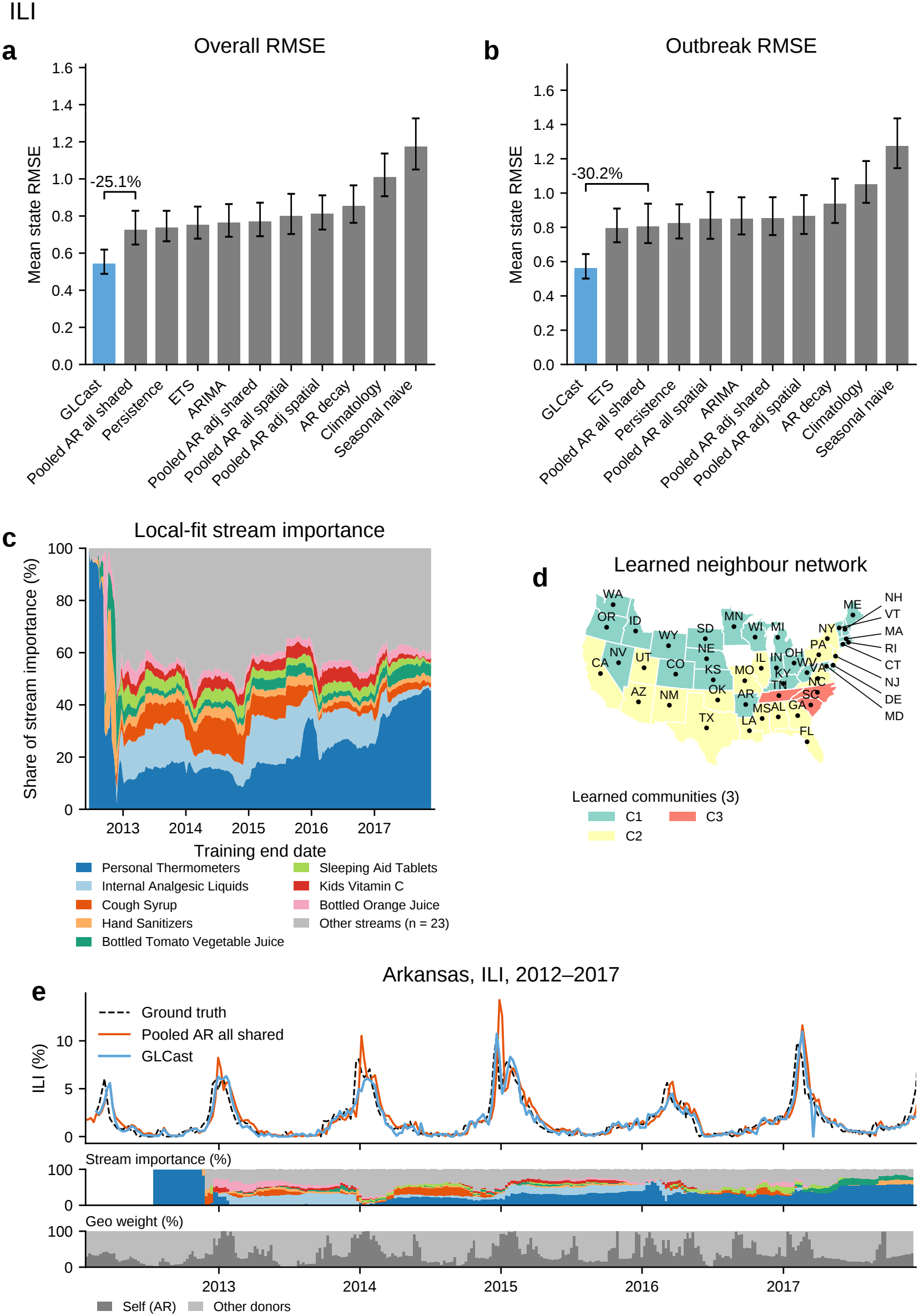
GLCast nowcasts of influenza-like illness with retail information. (**A**) Equal-state mean RMSE in ILI percentage points across 45 states for all scored weeks. Error bars are descriptive 95% BCa intervals from 10,000 state bootstrap resamples, conditional on the historical benchmark choice of specification. The bracket gives percentage change against the fixed Pooled AR all shared reference. (**B**) Outbreak weeks; the same reference is used although ETS has the lowest comparator RMSE in this stratum. National auxiliary-importance shares at retained retraining origins, with the top eight streams named and 23 pooled. Importance uses absolute local coefficients times rolling training-window standard deviations; zero-total origins are omitted from share means. (**D**) Leiden communities of the time-averaged, symmetrized display-share graph, excluding self-loops: three communities, *Q* = 0.192. (**E**) Arkansas observations and nowcasts, with state-specific auxiliary importance and normalized self-versus-donor shares. Predictions use raw blended weights and are floored at zero; normalization is for display. Further examples appear in *SI Appendix*, Fig. S1.

To understand how GLCast used retail information, we analyzed the importance of the NielsenIQ unit-sales streams (52), the stability of selected features, and geographic model borrowing. Within each local fit, we compute auxiliary-stream importance by summing absolute raw coefficients times the corresponding lagged predictors’ rolling training-window population standard deviations, then normalizing across auxiliary streams. First, retail importance was distributed across a broad set of product categories (Fig. 2*c*). The largest national mean auxiliary-importance shares were assigned to Personal Thermometers (25.8%), Internal Analgesic Liquids (10.4%), Cough Syrup (6.0%), and Hand Sanitizers (3.9%), while the 23 streams outside the displayed top eight together accounted for 40.0%. The complete profile for all 31 streams is shown in *SI Appendix*, Fig. S6. Several symptom-related retail streams also preceded ILI in the national correlation analysis: Adult Cough Cold Category (*r* = 0.52), Cough Syrup (*r* = 0.50), and Pediatric Cough Cold Category (*r* = 0.49) had the strongest stream– target correlations, each peaking when the stream led the target by three weeks (*SI Appendix*, Fig. S3). Second, local fits used retail inputs regularly and retained similar feature sets between successive refits. Only 10.1% of analyzed fits selected no auxiliary stream; the median fit retained 13 auxiliary streams and 14 total features, and successive nonempty selected-feature sets had a mean Jaccard similarity of 0.880. Contemporaneous retail inputs also accounted for most of the total coefficient mass: after scaling absolute coefficients by each predictor’s training-window standard deviation, their share was 74.6%, compared with 25.4% for target history (*SI Appendix*, Fig. S2). Third, geographic borrowing showed regional structure alongside changing donor membership. We summarized borrowing in a network whose connections represent normalized weights assigned to other states’ models, averaged over time and across the two directions of borrowing. The Leiden community-detection algorithm (63) identified three communities, grouping states with relatively strong borrowing connections within each group (modularity *Q* = 0.192; Fig. 2*d*). These comprised a compact North Carolina–South Carolina–Tennessee group and broader northern and southern groups. Individual donor candidates changed over time: after averaging the weekly fraction of the five donor candidates replaced within each state, the median across all 45 states was 30.3% (*SI Appendix*, Fig. S7).

We plot a representative example, Arkansas, to illustrate the dynamic stream and neighbor importance (Fig. 2*e*). Here we see how GLCast effectively uses auxiliary information to avoid over-predicting outbreak peaks against the strongest baseline. We see that outbreak peaks appear to be change points for feature importance values, for instance, Internal Analgesic Liquids hover around 30% importance in 2013, but after the peak at the start of 2014 reduces to almost 0%. Additionally, the share of neighboring importance also varies with outbreak peaks, with Arkansas relying heavily on its own information after an outbreak has begun and less so during down periods. We also see this pattern systematically across states; Thermometers account for 28.2% of auxiliary importance during outbreaks and 23.4% outside them in *SI Appendix*, Fig. S8. We show five additional states with similar patterns in *SI Appendix*, Fig. S1.

We perform 4 secondary analyses to support our conclusions. First, we vary the metric used to assess performance to mean absolute error (MAE) and *R*^2^. GLCast has the lowest MAE and highest mean *R*^2^ in all three evaluation strata (*SI Appendix*, Fig. S9). Second, we vary the size of the lag set chosen to four and eight most recent observations. Expanding both auxiliary and target histories increases overall RMSE by 3.2% and 4.0%, respectively (*SI Appendix*, Fig. S10). Third, we ablate components of GLCast. Removing geographic aggregation has the largest effect and increases RMSE by 8.9% (*SI Appendix*, Fig. S11). Varying the number of donor states from three to ten mildly reduces RMSE from 0.5442 to 0.5435 (*SI Appendix*, Fig. S12). Fourth, we vary community assignment methods between Leiden, Louvain, and Spectral. While assignments may vary between methods, all methods recapitulate exactly 3 clusters and a north-south split (*SI Appendix*, Fig. S13).

In summary, we conclude GLCast is a powerful method for nowcasting ILI using auxiliary retail and neighbor graph information.

### Nowcasting COVID-19 with search trends and surveys

We apply GLCast to daily COVID-19 hospital admissions across the 50 U.S. states and the District of Columbia, in conjunction with 11 auxiliary streams from Google symptom searches and the Facebook COVID-19 Trends and Impact Survey (CTIS) (4, 23, 53). These comprise seven search sets and four survey measures of symptoms, community illness, and testing (Materials and Methods; Table S3). We evaluate on 55,080 jurisdiction-days from April 26, 2021, through April 9, 2024. We apply LRTrend to retrospectively annotate outbreak periods, which cover 13.3% of scored jurisdiction-days.

Fig. 3*a* reports the mean across jurisdictions of perjurisdiction RMSE. GLCast reaches RMSE of 13.91 daily admissions against 15.98 for the strongest overall comparator, Pooled AR all shared, a reduction of 12.9% (paired difference 2.07; 95% BCa interval [1.52, 2.89]). The reduction against this same comparator is 13.8% inside outbreaks (difference 3.32; [2.37, 4.75]; Fig. 3*b*) and 12.6% outside them (difference 1.81; [1.32, 2.55]). At the individual-jurisdiction level, GLCast improves on Pooled AR all shared in 49 of 51 jurisdictions, with a median reduction of 10.6% and a maximum of 18.9% in California. Error is only slightly higher in the District of Columbia (+0.6%) and North Dakota (+0.3%). Additional comparisons during non-outbreak periods and across jurisdictions are shown in *SI Appendix*, Figs. S14 and S15, respectively.

**Fig. 3.**
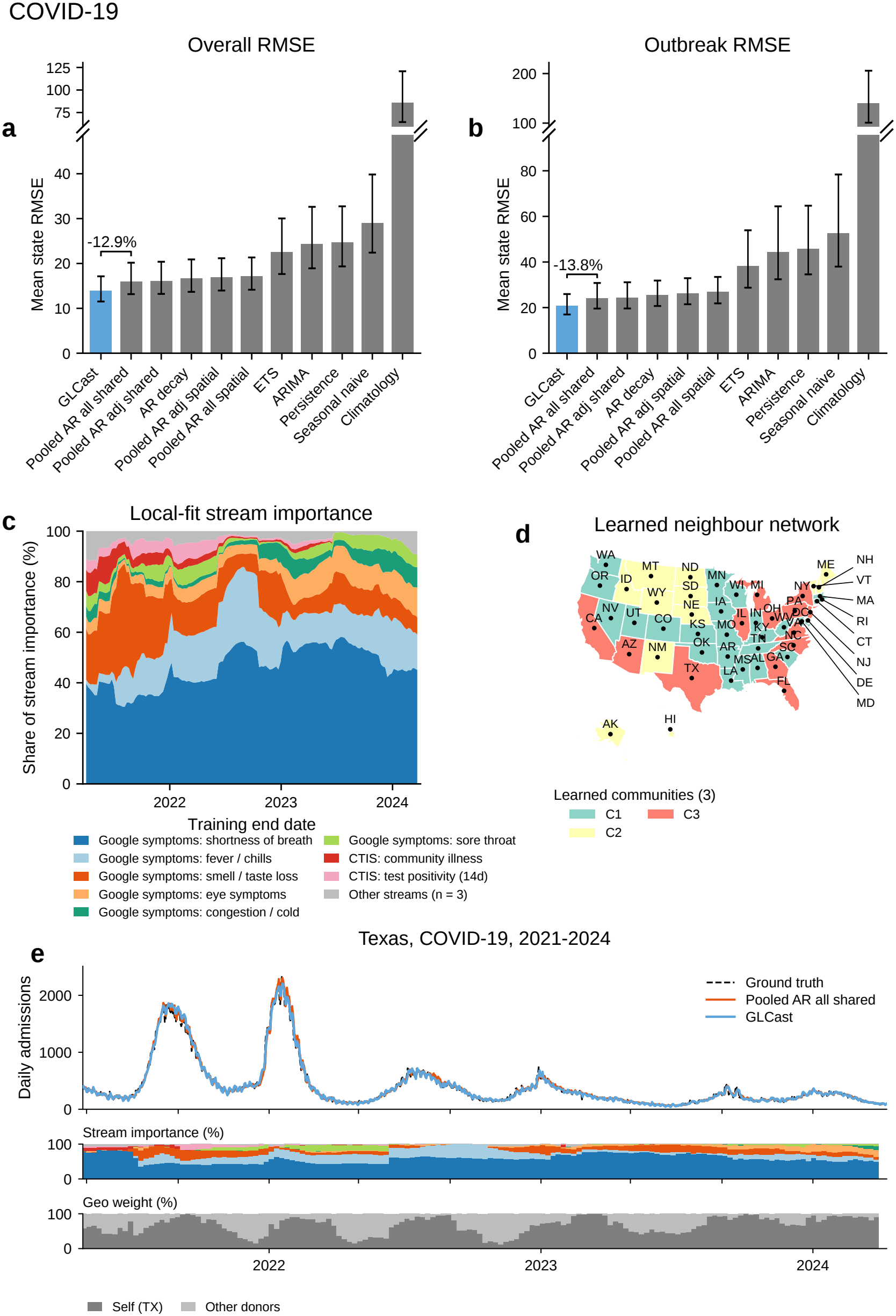
GLCast nowcasts of COVID-19 hospital admissions with search and survey information. (**A**) Equal-jurisdiction mean RMSE across 51 jurisdictions for all scored days, in daily admissions. (**B**) RMSE during outbreak days. The axes are broken to include Climatology. Error bars are descriptive 95% BCa intervals from 10,000 jurisdiction bootstrap resamples; brackets compare with Pooled AR all shared. (**C**) Time-varying auxiliary-importance shares for the eight leading search and survey streams, with three others pooled, based on absolute local coefficients scaled by training-window predictor variation. (**D**) Three Leiden communities in the time-averaged model-weight network (*Q* = 0.329). (**E**) Texas observations and nowcasts over all 1,080 scored days, compared with its best overall baseline, Pooled AR all shared. GLCast reduces RMSE by 18.1% (43.89 versus 53.58 admissions per day). The lower strips show Texas auxiliary importance and normalized local-versus-donor model weights.

We applied the same auxiliary information analyses to the Google symptom-search and CTIS survey streams, which measure relative search activity and survey-reported symptoms, community illness, and testing, respectively (23, 53). First, auxiliary importance was concentrated in symptom searches (Fig. 3*c*). The largest national mean shares were assigned to shortness of breath (46.2%), fever/chills (16.1%), smell/taste loss (15.1%), and eye symptoms (6.8%); the four CTIS measures together accounted for 7.3% (*SI Appendix*, Fig. S16). The national input–target correlation analysis identified strong associations in both data sources: smell/taste loss had the largest correlation (*r* = 0.82, with the stream leading the target by nine days), followed by CTIS test positivity (*r* = 0.72, contemporaneous) and CTIS community illness (*r* = 0.67, with a five-day lead). The CTIS inputs stopped updating in mid-2022 (*SI Appendix*, Fig. S18), but this was effectively handled by GLCast’s dynamic selection strategy. Second, auxiliary streams consistently supplemented target history, which carried the largest share of total scaled coefficient mass. Every analyzed local fit selected at least one auxiliary stream, the median fit contained seven auxiliary streams and 19 total features, and successive selected-feature sets had a mean Jaccard similarity of 0.893. Target history accounted for 62.8% of total scaled coefficient mass, contemporaneous auxiliary values for 20.3%, and older auxiliary values for 16.9% (*SI Appendix*, Fig. S17). Third, the borrowing network grouped geographically distant jurisdictions, while donor membership changed over time. Applying Leiden to the time-averaged regional-model network identified three communities (*Q* = 0.329; Fig. 3*d*). California, Texas, and Florida shared a community with New York, New Jersey, and Pennsylvania, while Washington and Oregon lay in another. The California–Texas–Florida grouping resembles a cluster reported in our earlier COVID-19 neighbor graph analysis using different datasets (41). After averaging the weekly fraction of the five donor candidates replaced within each jurisdiction, the median across all 51 jurisdictions was 20.6% (*SI Appendix*, Fig. S7).

We plot Texas to illustrate the changing stream and geographic shares (Fig. 3*e*; additional examples in *SI Appendix*, Fig. S19). Here, GLCast reduces RMSE by 18.1% against the strongest baseline for Texas, Pooled AR all shared, from 53.58 to 43.89 daily admissions across all 1,080 scored days. The nowcasts closely follow the summer 2021 and winter 2021–2022 waves and subsequent smaller waves. Shortness-of-breath searches remain the largest source of auxiliary importance, while fever/chills searches become more prominent in 2022 and eye-symptom searches receive a larger share later in the series. The geographic shares also change, with the contribution from other states declining in 2023 and early 2024. We also summarize information use across jurisdictions and epidemic phases: smell/taste-loss searches account for 21.6% of auxiliary importance during outbreaks and 13.4% outside them, while shortness-of-breath searches account for 31.0% and 39.4%, respectively. Normalized same-state model weight averages 83.5% during outbreaks and 80.5% outside them (*SI Appendix*, Fig. S20). Six additional state examples are shown in *SI Appendix*, Fig. S19.

We perform 4 secondary analyses to support our conclusions. First, we vary the performance metric to mean absolute error (MAE) and *R*^2^. GLCast has the lowest MAE and highest mean *R*^2^ in all three evaluation strata (*SI Appendix*, Fig. S9). Second, we expand the lag histories to one and two weeks. Overall RMSE decreases by 0.7% and 2.7%, respectively (*SI Appendix*, Fig. S21). Third, we ablate components of GLCast. Removing geographic aggregation increases RMSE by 4.8%, splitting auxiliary streams into singleton groups increases it by 0.17%, and removing the within-group coordinate penalty increases it by 1.2% (*SI Appendix*, Fig. S11). Varying the donor cap from three to ten mildly reduces RMSE from 13.9422 to 13.8701 (*SI Appendix*, Fig. S12). Fourth, we vary community assignment between Leiden, Louvain, and spectral clustering. Leiden and Louvain each return three communities, and spectral clustering uses this same count, although the assignments differ (*SI Appendix*, Fig. S22). In summary, we conclude that GLCast improves COVID-19 admission nowcasts by combining search, survey, and regional-model information.

### Nowcasting three diseases in the modern data ecosystem

We apply GLCast to seven weekly respiratory-surveillance targets across 51–52 U.S. jurisdictions: NHSN hospital admissions and NSSP emergency-department (ED) visit percentages for influenza, COVID-19, and RSV, together with outpatient ILINet %ILI (51, 54, 55). We combine each target with ten auxiliary streams: eight shared indicators of environmental conditions and travel, plus disease-specific Wikipedia page views and GDELT news volume (Materials and Methods; Table S4). We evaluate on 60,878 target-specific jurisdiction-weeks from February 11, 2023, through August 8, 2026, with RSV admissions beginning on January 13, 2024 (Table S1). We apply LRTrend to retrospectively annotate outbreak periods, which cover 17.6–34.9% of evaluated jurisdiction-weeks across the seven targets (Table S5).

Fig. 4*a* reports the reduction in mean jurisdiction-level RMSE against each target’s best overall baseline. GLCast has the lowest overall error for all seven targets. Against Persistence, error decreases by 20.5% for influenza admissions, 25.1% for influenza ED visits, and 22.5% for ILINet. Against the pooled autoregression with coefficients shared across states, reductions are 7.1% for COVID-19 admissions, 5.1% for COVID-19 ED visits, 0.1% for RSV admissions, and 6.0% for RSV ED visits. RSV admissions are nearly tied, with RMSEs of 39.98 and 40.02 weekly admissions; intervals for this target, RSV ED visits, and COVID-19 ED visits include zero reduction. Influenza and ILI gains are larger during outbreaks, while both RSV outcomes have higher outbreak error than their baseline. Full method, jurisdiction, and phase comparisons appear in *SI Appendix*, Figs. S23 and S24, and Table S6. We compared auxiliary-stream importance and geo-graphic borrowing during outbreak and non-outbreak periods. First, online attention accounted for most auxiliary importance, with a larger combined share during outbreaks for every target (Fig. 4*b*). National-level Wikipedia and news together accounted for 60.7– 91.3% of auxiliary importance during outbreaks and 50.1–86.2% outside them. The largest increases in their combined share occurred for RSV admissions (82.4% during outbreaks versus 68.7% outside them) and RSV ED visits (91.3% versus 79.1%); temperature had a smaller auxiliary-importance share during outbreaks for both targets. Second, models fitted in other states retained the majority of normalized aggregation weight in both phases for every target (Fig. 4*c*). The balance between same-state and other-state models also varied with the target and epidemic phase. For COVID-19 ED visits, the same-state share increased from 32.1% outside outbreaks to 43.6% during outbreaks.

**Fig. 4.**
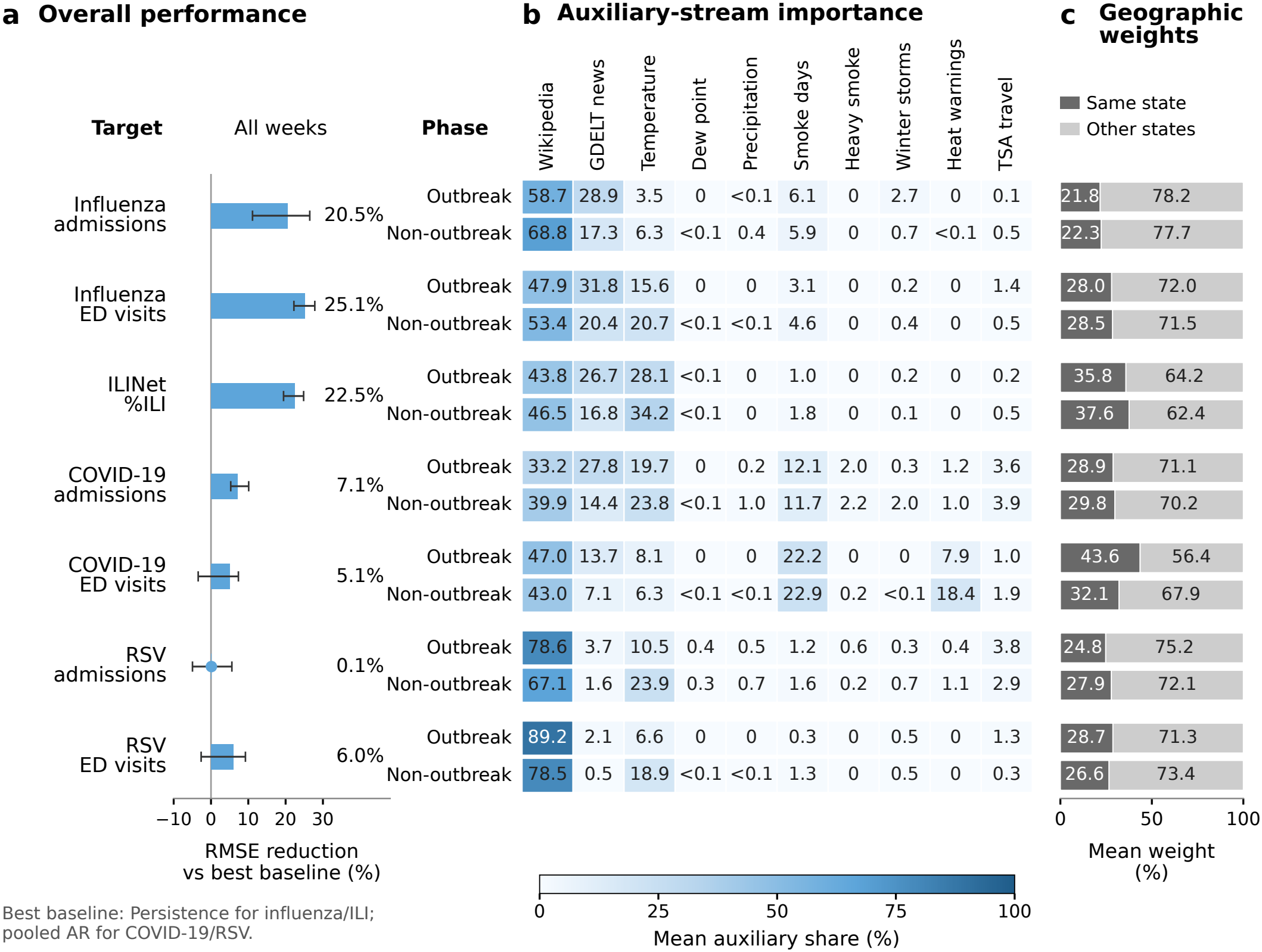
Modern respiratory nowcasting and target-specific information use. (**A**) Reduction in equal-jurisdiction mean RMSE against each target’s best overall baseline: Persistence for influenza/ILI and Pooled AR all shared for COVID-19/RSV. Positive values favor GLCast; whiskers are descriptive 95% paired-jurisdiction BCa intervals. (**B**) Mean auxiliary-importance shares for all ten streams, shown separately for each of the seven targets during outbreak and non-outbreak periods. Coefficient masses are summed across scored jurisdictions and normalized within each target, date, and phase, then averaged over defined weeks. (**C**) Mean normalized same-state and other-state model weights for the corresponding target and phase. Weights are normalized within each serving model, averaged across scored jurisdictions within each date and phase, then across weeks.

We perform 5 secondary analyses to support our conclusions. First, we compare outbreak and non-outbreak performance. Influenza and ILI reductions range from 23.5% to 28.8% during outbreaks and from 15.8% to 23.5% outside them; RSV outbreak error increases by 18.4% for admissions and 6.9% for ED visits, despite non-outbreak reductions of 7.3% and 12.8% (Table S6; additional admission comparisons in *SI Appendix*, Fig. S26). Second, we examine jurisdiction-level differences. Influenza admissions improve in 47 of 52 jurisdictions, influenza ED visits in all 51, and ILINet in 51 of 52, whereas RSV admissions improve in only 23 of 52 (*SI Appendix*, Fig. S24). Third, we require at least eight weeks in each phase of the shared-upswing comparison; the same four intervals include one and the influenza and RSV ED intervals remain above one (Table S7). Fourth, we examine all-stream importance and geographic networks separately by target. Wikipedia and news account for 53.0– 87.4% of mean auxiliary importance across targets, and the averaged admissions networks form three influenza, three COVID-19, and two RSV communities (*SI Appendix*, Figs. S27 and S28). Fifth, we examine how GLCast performs when different respiratory viruses rise at the same time (*SI Appendix*, Fig. S25). Comparing GLCast with itself, the shared-to-own-only ratios of amplitude-relative error are 0.97 and 1.04 for COVID-19 admissions and ED visits. The intervals include one for both COVID-19 targets and for influenza and RSV admissions, while influenza and RSV ED visits have higher relative error during shared upswings, with ratios of 1.31 ([1.21, 1.43]) and 1.24 ([1.14, 1.40]).

In summary, we conclude GLCast can improve nowcasting across respiratory diseases using context, attention, and regional-model information.

## Discussion

We introduced GLCast, a statistical framework for now-casting delayed epidemic surveillance measurements by combining target histories, auxiliary data streams, and models fitted in other geographic regions. Through extensive retrospective analyses, we demonstrated that GLCast improves nowcasting across different diseases, surveillance endpoints, and information environments. We applied GLCast to six years of weekly ILI with retail sales, three years of daily COVID-19 hospital admissions with search and survey information, and seven modern respiratory targets with context and online-attention streams (Figs. 2–4). GLCast reduced mean RMSE by 25.1% for historical ILI and 12.9% for historical COVID-19 admissions against the strongest overall target-only comparators, and achieved the lowest overall point error across the seven modern targets. Analyses of the fitted models further revealed that selected streams, donor identities, and geographic weights change across time and space, with differences in information use between outbreak and non-outbreak periods. These results provide a framework for improving epidemic monitoring while characterizing how different sources of information contribute to nowcasting. A key advantage of GLCast is its ability to repeatedly select useful streams from a supplied data library and learn which regional models contribute to each local estimate, which is particularly valuable when the relationships between auxiliary measurements and disease activity change over time (42, 50). In addition, geographic aggregation allows local nowcasts to use predictive relationships learned in other regions, including regions that are not immediate geographic neighbors. This approach builds on previous work combining autoregression, digital indicators, and geographic information (30, 33), bringing stream selection and regional-model borrowing into a common framework with explicit summaries of how both change over time.

We have demonstrated the value of GLCast in improving epidemic nowcasts, characterizing changes in the use of auxiliary information, and identifying regional relationships that contribute to local predictions. These analyses can help epidemiologists investigate why particular streams become more or less informative and prioritize candidate data sources for continued collection and prospective evaluation. For example, the larger importance of thermometers during ILI outbreaks and online attention during modern respiratory outbreaks suggests that different types of measurements may be useful at different stages of epidemic activity (Fig. 4*b*; *SI Appendix*, Fig. S8). Disease-specific nowcasts on a common calendar may also support situational awareness when several respiratory viruses circulate simultaneously. As additional surveillance and auxiliary datasets become available, these analyses could enable more detailed characterization of the information needed to monitor different diseases, locations, and epidemic phases.

We note several limitations and directions for future work. First, stream importance and geographic weights describe predictive associations and do not directly establish causal relationships, transmission pathways, or outbreak change points. Correlated streams can share or exchange importance, and local coefficient profiles do not fully decompose the final geographically aggregated prediction. Nonetheless, these summaries provide interpretable hypotheses about information use that can be examined through independent data and targeted analyses. Second, GLCast currently produces point estimates. The intervals reported here summarize performance comparisons and do not quantify uncertainty around individual nowcasts; extending the framework to calibrated probabilistic predictions would improve its usefulness for decisions under uncertainty. Third, we focused on respiratory surveillance at the U.S. state and jurisdiction level. Applications to other pathogens, geographic scales, and surveillance systems remain to be evaluated, as does the value of additional sources such as medical claims, wastewater, and wearable measurements (24, 25, 39).

## Materials and Methods

In the following sections, we describe the data and statistical components of GLCast. The goal is to estimate a delayed surveillance measurement from its recent history and auxiliary measurements indexed to the same target date. The framework first selects useful streams through adaptive grouped regression, then identifies models from other locations with similar predictive relationships, and combines their predictions according to their performance on recent target-location data. The same regularization and aggregation procedure is used for all nine targets; lag sets, fitting windows, and prediction schedules reflect the temporal resolution and reporting structure of each setting (Table 1).

### Data sources

We evaluate GLCast in three complementary surveillance settings. The classic influenza analysis combines weekly outpatient illness with retail purchases; the classic COVID-19 analysis combines daily hospital admissions with survey and search activity; and the modern analysis combines seven weekly respiratory targets with environmental and online-attention streams. Together, these settings span different diseases, temporal resolutions, surveillance outcomes, and auxiliary data modalities. Table S1 summarizes their definitions, evaluation periods, and geographic coverage.

### Influenza-like illness

The classic influenza target is weekly state-level influenza-like illness (ILI) from the CDC’s ILINet outpatient surveillance network, distributed through Delphi Epidata (4, 51). ILI measures the percentage of outpatient visits for illness defined by fever together with cough or sore throat, rather than laboratory-confirmed influenza. We evaluate both outbreak and non-outbreak weeks throughout the year.

### Retail product sales

The classic ILI auxiliary data comprise weekly state-level retail unit sales for 31 NielsenIQ product categories (52). These include cough and cold remedies, allergy and sinus products, thermometers, tissues, disinfectants, hand sanitizers, analgesics, juices, and related products (Table S2). The library was assembled using prior correlation screening and domain knowledge; GLCast selects among its streams at each refit. Such purchases can capture changes in symptoms and health-related behavior, complementing previous work using supermarket purchases, digital prescriptions, and over-the-counter drug sales for respiratory surveillance (64–66). Each retail category defines its own regression group; the broader product families in the table are display categories.

### COVID-19 admissions, surveys, and searches

The classic COVID-19 target is the daily state-level HHS `confirmed admissions covid 1d` signal distributed through Delphi COVIDcast, which records incident confirmed COVID-19 hospital admissions (4). Four auxiliary streams from the Delphi/Facebook COVID-19 Trends and Impact Survey (CTIS) measure COVID-like illness, illness in the respondent’s community, recent testing, and recent test positivity (23). These are survey-weighted percentages. Testing covers the preceding 14 days, and positivity is calculated among respondents tested during that period. Seven further streams summarize Google searches for predefined symptom sets (53). Search indices are normalized within location and expressed in arbitrary units, so their levels are not directly comparable across states. Table S3 gives the stream identifiers and definitions. The analysis retains the constant-filled survey histories following the end of CTIS collection.

### Modern surveillance targets

The seven modern outcomes comprise weekly NHSN hospital admissions and NSSP emergency-department visit percentages for influenza, COVID-19, and RSV, together with outpatient ILINet %ILI (51, 54, 55). Admissions are counts; NSSP and ILINet outcomes are percentages. Their RMSEs are therefore expressed in admissions or percentage points, respectively. Target histories begin in January 2022 for COVID-19 and influenza admissions and ILINet, October 2022 for NSSP, and October 2023 for RSV admissions. NSSP covers 51 jurisdictions; the remaining targets also include Puerto Rico. The disease input tables contain 242 Saturday-ending weeks from January 8, 2022, through August 22, 2026, with target-specific availability and fitting requirements determining the scored observations. NHSN measures reported admissions, and its reporting requirements changed during the study (67).

### Modern context and attention streams

Eight context streams describe environmental conditions and travel: NOAA temperature, dew point, and precipitation; wildfire-smoke and heavy-smoke days from the NOAA Hazard Mapping System; winter-storm and heat warnings from the National Weather Service; and TSA airport-security checkpoint passenger throughput (56–59). Two disease-specific streams measure online attention through Wikipedia page views and GDELT news volume (60, 61). Weather, smoke, and warning measurements vary geographically. TSA, Wikipedia, and news measurements are national series repeated across jurisdictions, allowing each local model to estimate its own relationship between these shared temporal signals and the target. Each disease therefore has ten auxiliary streams, with fourteen distinct streams across the three diseases (Table S4). No additional disease-surveillance stream is used as an auxiliary predictor; each target retains its own lagged values in the autoregressive group. The analysis uses weekly observations from these sources without reconstructing historical release vintages.

### Alignment and preprocessing

Within each location, auxiliary streams are aligned to the target calendar before constructing lagged predictors. Streams absent from that location’s input table are omitted, and rows with missing required predictors or responses are excluded after lagging. Observed values, including negative temperatures, are retained on their original scales. Predictors and responses are then standardized within each fitting window, with the same fitted transformation used to recover raw coefficients and intercepts (Appendix B).

### Outbreak periods

We evaluate accuracy separately during sustained increases in reported disease activity and during other periods. Outbreak windows are identified retrospectively in each location’s target series using LRTrend (41). These labels define evaluation strata and are not supplied to the models as predictors. Outbreak periods comprise 46.5% of evaluated ILI state-weeks, 13.3% of COVID-19 state-days, and 17.6–34.9% of modern state-weeks, depending on the target (Table S5).

### Grouped local regression

GLCast first fits a regression within each location to estimate a surveillance measurement from its own history and auxiliary data. We group the lags of each auxiliary stream so that the model can remove an entire stream, while an additional penalty can remove individual lags. This allows the fitted relationship to use a subset of the available information (68–70).

Let *Y*_*ℓ,t*_ be the surveillance value at location *ℓ* and target date *t*, and let 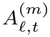 denote auxiliary stream *m*. The predictor vector *x*_*ℓ,t*_ contains auxiliary measurements 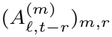 and target-history measurements (*Y*_*ℓ,t−k*_)_*k*_, with lag sets given in Table 1. The response is *Y*_*ℓ,t*_, so an auxiliary lag of zero refers to the date being estimated. Auxiliary streams form separate, nonoverlapping groups, and target-history columns form one additional group. The main weekly designs have one column per group; the daily COVID-19 design includes several lags per stream.

At each refit, we center and scale the predictors and response using their local fitting-window means and standard deviations. Write *Z*_*ℓ*_ and 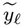 for the standardized data, *n*_*ℓ*_ for the number of complete fitting rows, and *b*_*g*_ for the coefficients in group *g*. The local fit uses the adaptive sparse-group criterion

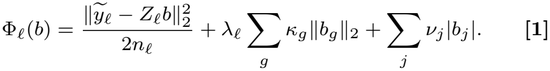

The first term measures prediction error, the second shrinks entire groups, and the third shrinks individual coefficients. We use *λ*_*ℓ*_ = *α* and *ν*_*j*_ = 0.7*αd*_*j*_, where *d*_*j*_ = *r*_*j*_ + 1 for an auxiliary column at lag *r*_*j*_ and *d*_*j*_ = 1 for a target-history column. Thus, older auxiliary lags receive larger coordinate penalties, while target-history coefficients are also penalized. The group weights *κ*_*g*_ are normalized inverse pilot-selection frequencies, so streams selected more often in the pilot receive smaller group penalties.

All nine analyses use this construction, with target-specific pilot weights and automatically selected *α* values (Appendix B). We compute coefficients with a screened iterative approximation that applies group shrinkage followed by coordinate shrinkage and checks excluded groups for re-entry. Appendix C gives the numerical updates and stopping rules.

### Transfer across locations

We next identify models from other locations whose predictive relationships may help the target location. For target location 0, we first compare each source’s fitting-period standardized feature–response cross-moments with those of the target. Up to 20 sources with the smallest discrepancies form a candidate library. We then rank the eligible source models by prediction error on the target’s calibration window and retain up to five, together with the local model (71). Screening and ranking are repeated at each refit, so the donor set can change over time. Appendix D gives the discrepancy score and eligibility rules.

To evaluate a source model on the target’s measurements, we first recover its raw-scale coefficient vector 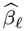 and intercept 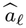. Let *P*_*ℓ→* 0_ align source coefficients to the target’s feature names, assigning zero to unmatched target coordinates. The transported prediction is

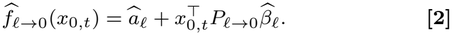

Thus, each source model applies its own fitted relationship to the target region’s raw predictors, retaining the source intercept and raw coefficients. These connections describe the use of other locations’ predictive models, rather than inferred transmission routes.

### Nonnegative stacking and Q-inspired aggregation

Finally, we combine the local and retained source predictions using two complementary weighting rules. Nonnegative stacking fits regression weights to the target’s calibration outcomes, with shrinkage on the source weights. A Q-inspired exponential-weight procedure uses candidate errors and disagreement with the current aggregate to construct weights that sum to one. Their mixture depends on the number of calibration observations.

Write the retained candidate set as *C*_0_ = {*c*_1_, …, *c*_*J*_ }, with *c*_1_ = 0 denoting the local model. Let *F*∈ ℝ^*m×J*^ contain their unfloored predictions on the *m* target calibration rows, and let *y*^v^ contain the corresponding outcomes. Both are in raw target units. The stacking weights are

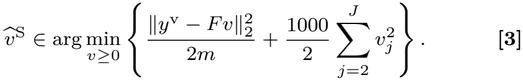

The ridge term shrinks the source weights while leaving the local weight unpenalized. There is no additional intercept, and the stacking weights need not sum to one. The second branch uses a finite, damped exponential-weight iteration after scaling both the outcomes and candidate predictions by the calibration-response standard deviation. It returns 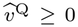 with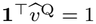 . This Q-inspired procedure follows the prediction-aggregation approach in (71, 72); Appendix E specifies its initialization, updates, and fixed settings.

The final weights and prediction are

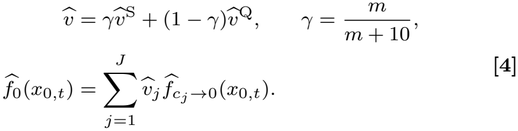

As the calibration sample grows, the blend gives more weight to stacking. The final weights are nonnegative but need not sum to one. We use these raw weights for prediction and normalize them only for network and percentage displays. The zero floor is applied after combination for scoring and plotting.

For a fixed response vector, the raw blend also has a simple squared-loss interpretation. With 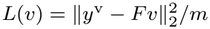, convexity gives

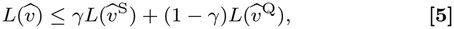

so its loss is bounded by the corresponding weighted average of the two branch losses. This deterministic inequality does not guarantee improvement over the best individual candidate or performance on new data (Appendix F).

### Stream importance

Let *A* denote the set of auxiliary streams, excluding autoregressive target-history terms, and let *ℒ*_*s*_ denote the lag set for stream *s*. For jurisdiction *i* and fit origin *t*, we define the stream’s coefficient mass and local importance share as

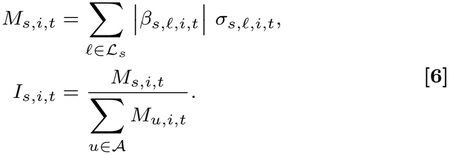

Here, *β*_*s,ℓ,i,t*_ is the raw-scale coefficient from the local model before geographic aggregation, and *σ*_*s,ℓ,i,t*_ is the population standard deviation of the corresponding lagged predictor over the rolling training window, with the window shifted to match lag *ℓ*.

For all-period national summaries, we first pool stream masses across jurisdictions at each fit origin, normalize across auxiliary streams, and then average the resulting shares equally across origins. Phase-specific summaries use the coefficients from the fit serving each scored date. For the classic ILI and COVID-19 analyses, we normalize locally, average the defined local shares equally across jurisdictions within each date and outbreak or non-outbreak phase, and then average equally across dates. For the modern targets, we pool stream masses across jurisdictions within each date and phase, normalize across auxiliary streams, and then average equally across dates.

### Retrospective evaluation

We refit GLCast on rolling windows and evaluate its predictions on the retained dates for each target. Fitting, calibration, and test-window lengths are 360, 30, and 30 days for classic COVID-19 and 55, 4, and 4 weeks for classic ILI and the modern targets. Calibration uses the trailing observations within the fitting window. Local regressions, standardization, and source screening use the fitting rows; donor ranking and aggregation use the calibration rows. Table S8 gives the refit schedules, and Appendix H defines the scoring populations and secondary analyses.

We compare GLCast with ten target-only baselines spanning local autoregression, classical univariate methods, and geographically pooled or spatial-lag autoregressions (Appendix G). After flooring predictions at zero, we calculate RMSE within each jurisdiction and average equally across jurisdictions. Descriptive 95% bias-corrected and accelerated (BCa) intervals use 10,000 whole-jurisdiction bootstrap resamples, paired for method contrasts (73). These intervals condition on the fitted models and historical specification. Classic analyses use Pooled AR all shared as the fixed reference; modern analyses use each target’s lowest-error overall baseline, held fixed across epidemic phases. The Texas example uses its locally best baseline over the full scoring period. Additional analyses assess alternative metrics, lag histories, model components, and donor counts, and summarize how stream selection and geographic weights change over time and between epidemic phases. For the modern disease-specific targets, we also compare GLCast’s amplitude-relative error during own-only and shared upswings (Fig. S25; Table S7).

### Data, Materials, and Software Availability

The GLCast repository is available at github.com/Alistair-Turcan/GLCast. Scripts for reproducing all analyses in this paper are located at github.com/Alistair-Turcan/GLCast-paper. The historical admissions, CTIS, and Google Symptoms streams are distributed through Delphi Epidata; ILINet, NHSN, and NSSP outcomes are provided by the CDC; and the modern context and attention sources are identified in Table S4. NielsenIQ retail sales are proprietary and were accessed under license.

## Data Availability

All data are available online at: NielsenIQ retail-sales data (https://nielseniq.com/global/en/products/retail-measurement-services-rms/); CDC ILINet influenza-like illness data distributed through Delphi FluView (https://cmu-delphi.github.io/delphi-epidata/api/fluview.html); HHS/NHSN hospital-admission and NSSP emergency-department surveillance data (https://cmu-delphi.github.io/delphi-epidata/api/covidcast_signals.html); Google symptom-search trends (https://cmu-delphi.github.io/delphi-epidata/api/covidcast-signals/google-symptoms.html); and aggregate COVID-19 Trends and Impact Survey measurements (https://cmu-delphi.github.io/delphi-epidata/api/covidcast-signals/fb-survey.html). Environmental inputs comprised NOAA GSOD weather observations (https://www.ncei.noaa.gov/access/metadata/landing-page/bin/iso?id=gov.noaa.ncdc%3AC00516), Iowa Environmental Mesonet ASOS observations (https://iem.weather.im/request/download.phtml), NOAA HMS smoke polygons (https://ospo.noaa.gov/products/land/hms.html), and National Weather Service warning records archived by the Iowa Environmental Mesonet (https://mesonet.agron.iastate.edu/request/gis/watchwarn.phtml). Additional inputs were TSA passenger counts (https://www.tsa.gov/travel/passenger-volumes), Wikimedia disease-article pageviews (https://doc.wikimedia.org/generated-data-platform/aqs/analytics-api/examples/page-metrics.html), and GDELT disease-related news counts (https://blog.gdeltproject.org/gdelt-2-0-api-now-supports-raw-result-counts/).

https://github.com/Alistair-Turcan/GLCast-paper/tree/main

https://github.com/Alistair-Turcan/GLCast/tree/main

## ACKNOWLEDGMENTS

This material is based upon work supported by the United States of America Department of Health and Human Services, Centers for Disease Control and Prevention, under award numbers U01IP001121 and NU38FT000005; and contract number 75D30123C1590. Any opinions, findings, and conclusions or recommendations expressed in this material are those of the author(s) and do not necessarily reflect the views of the United States of America Department of Health and Human Services, Centers for Disease Control and Prevention. We thank Roni Rosenfeld for useful advice throughout the project. We acknowledge the use of Biomni Lab (74) for performing and reproducing analyses and GPT-in helping polish the manuscript.

## SI Appendix

**Table S1.** Evaluation targets and data coverage.

| Setting | Target / source | Unit | Scored dates | Jurisd. | Cells |
|---|---|---|---|---|---|
| Classic | ILI / ILINet | Percentage | 19 Feb 2012–10 Dec 2017 | 45 | 13,680 |
| Classic | COVID-19 / HHS | Daily admissions | 26 Apr 2021–9 Apr 2024 | 51 | 55,080 |
| Modern | Influenza admissions | Weekly admissions | 11 Feb 2023–8 Aug 2026 | 52 | 9,301 |
| Modern | Influenza ED visits | Percentage | 11 Feb 2023–8 Aug 2026 | 51 | 9,167 |
| Modern | ILINet %ILI | Percentage | 11 Feb 2023–8 Aug 2026 | 52 | 9,052 |
| Modern | COVID-19 admissions | Weekly admissions | 11 Feb 2023–8 Aug 2026 | 52 | 9,301 |
| Modern | COVID-19 ED visits | Percentage | 11 Feb 2023–8 Aug 2026 | 51 | 9,180 |
| Modern | RSV admissions | Weekly admissions | 13 Jan 2024–8 Aug 2026 | 52 | 5,779 |
| Modern | RSV ED visits | Percentage | 11 Feb 2023–8 Aug 2026 | 51 | 9,098 |

**Table S2.** Retail categories used as auxiliary streams for classic ILI. These 31 weekly unit-sales categories form the prespecified retail library. Product families are display headings; each category is a separate regression group. The main design contains one contemporaneous value per category, so its auxiliary groups are singletons. Expanded-history sensitivities group multiple lags within each category.

| Product family | Categories |
|---|---|
| Vitamins and supplements | Vitamin C Type; Vitamin C Immunity Category; Kids Vitamin C |
| Cough and cold | Adult Cough Cold Category; Pediatric Cough Cold Category |
| Allergy and sinus | Adult Sinus Allergy Category; Pediatric Allergy Sinus Category |
| Upper respiratory relief | Total Upper Respiratory; Cough Drops; Cough Syrup; Sore Throat Remedy Liquids |
| Cold/allergy/sinus remedies | Cold Allergy Sinus Liquid; Cold Allergy Sinus Tablets |
| Nasal care | Nasal Aspirators; Nasal Spray / Drops / Inhalers; Nasal Strips |
| Antacids | Antacid Liquid / Powder |
| Analgesics | Internal Analgesic Liquids; Internal Analgesic Tablets |
| Sleep aids | Sleeping Aid Liquids; Sleeping Aid Tablets |
| Diagnostics | Personal Thermometers |
| Juices | RFG Orange Juice; Bottled Apple Juice; Bottled Grape Juice; Bottled Orange Juice; Bottled Tomato Vegetable Juice |
| Sanitizers and cleaners | Hand Sanitizers; All Purpose Cleaner / Disinfectant |
| Personal care | Lip Treatment; Facial Tissue |

**Table S3.**
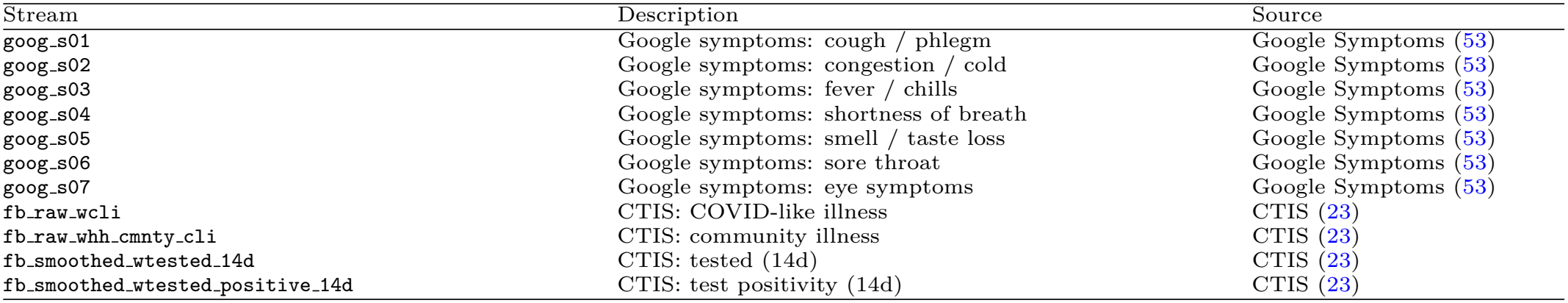
Auxiliary streams for COVID-19 nowcasting. Seven Google Symptoms sets and four COVID-19 Trends and Impact Survey (CTIS) signals, distributed through Delphi COVIDcast. Each stream is a separate feature group.

| Stream | Description | Source |
|---|---|---|
| goog_s01 | Google symptoms: cough / phlegm | Google Symptoms (53) |
| goog_s02 | Google symptoms: congestion / cold | Google Symptoms (53) |
| goog_s03 | Google symptoms: fever / chills | Google Symptoms (53) |
| goog_s04 | Google symptoms: shortness of breath | Google Symptoms (53) |
| goog_s05 | Google symptoms: smell / taste loss | Google Symptoms (53) |
| goog_s06 | Google symptoms: sore throat | Google Symptoms (53) |
| goog_s07 | Google symptoms: eye symptoms | Google Symptoms (53) |
| fb_raw_wcli | CTIS: COVID-like illness | CTIS (23) |
| fb_raw_whh_cmnty_cli | CTIS: community illness | CTIS (23) |
| fb_smoothed_wtested_14d | CTIS: tested (14d) | CTIS (23) |
| fb_smoothed_wtested_positive_14d | CTIS: test positivity (14d) | CTIS (23) |

**Table S4.**
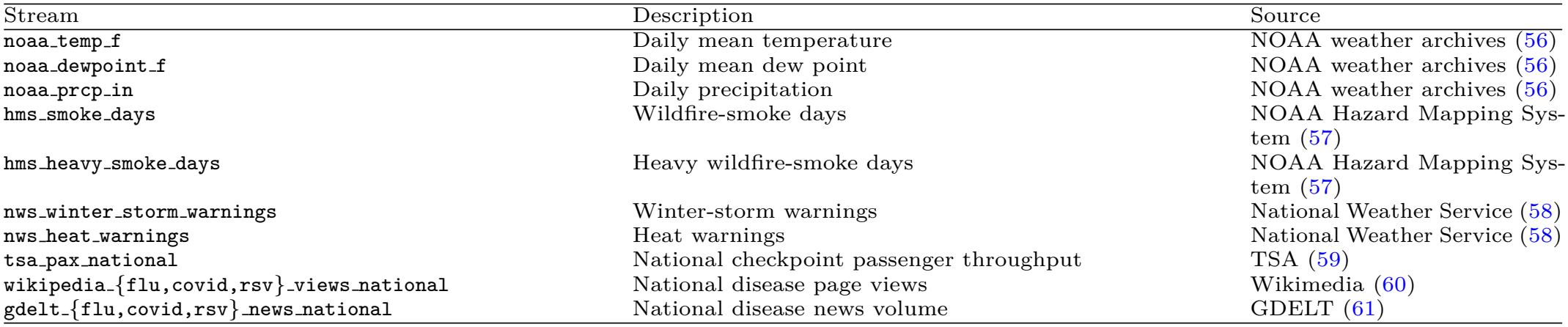
Auxiliary streams in the modern analyses. Eight context streams are shared across diseases, and Wikipedia and news streams are disease-specific. The ten inputs per disease contain no additional disease-surveillance measurement. National TSA, Wikipedia, and GDELT values are repeated across jurisdictions.

| Stream | Description | Source |
|---|---|---|
| noaa_temp_f | Daily mean temperature | NOAA weather archives (56) |
| noaa_dewpoint_f | Daily mean dew point | NOAA weather archives (56) |
| noaa_prcp_in | Daily precipitation | NOAA weather archives (56) |
| hms_smoke_days | Wildfire-smoke days | NOAA Hazard Mapping Sys-tem (57) |
| hms_heavy_smoke_days | Heavy wildfire-smoke days | NOAA Hazard Mapping Sys-tem (57) |
| nws_winter_storm_warnings | Winter-storm warnings | National Weather Service (58) |
| nws_heat_warnings | Heat warnings | National Weather Service (58) |
| tsa_pax_national | National checkpoint passenger throughput | TSA (59) |
| wikipedia_{flu,covid,rsv}_views_national | National disease page views | Wikimedia (60) |
| gdelt_{flu,covid,rsv}_news_national | National disease news volume | GDELT (61) |

**Table S5.** Modern outbreak labels and evaluated coverage. Stored intervals are all jurisdiction-specific windows in the supplied retrospective label archive; contributing intervals contain at least one retained scored cell. Scored cells are jurisdiction-weeks, and shares use each target-specific retained evaluation mask.

| Target | Stored | Contributing | Outbreak cells | All scored cells | Share (%) |
|---|---|---|---|---|---|
| COVID-19 admissions | 469 | 349 | 2,535 | 9,301 | 27.3 |
| COVID-19 ED visits | 189 | 174 | 1,616 | 9,180 | 17.6 |
| Influenza admissions | 330 | 223 | 2,433 | 9,301 | 26.2 |
| Influenza ED visits | 221 | 172 | 2,629 | 9,167 | 28.7 |
| ILINet %ILI | 290 | 222 | 3,025 | 9,052 | 33.4 |
| RSV admissions | 166 | 147 | 1,662 | 5,779 | 28.8 |
| RSV ED visits | 179 | 159 | 3,171 | 9,098 | 34.9 |

**Table S6.** Modern paired comparisons with the baseline selected by overall error. The reference is the target-only method with the lowest observed all-period equal-jurisdiction mean RMSE among the ten retained baselines: persistence for influenza admissions, influenza ED visits, and ILINet; Pooled AR all shared for the four COVID-19 and RSV targets. Each choice is held fixed across all three strata. RMSE is averaged equally across jurisdictions, in weekly admission counts or percentage points. Reduction is 100(1− *e*_GLCast_*/e*_reference_); positive values favor GLCast. Intervals are descriptive paired-jurisdiction BCa 95% intervals for the percentage reduction (10,000 resamples, seed 7 separately for each target–stratum contrast). Both mean errors are recomputed in each resample. All listed comparisons use identical retained cells for both methods.

| Target | Stratum | GLCast | Reference | Reduction % [95% interval] |
|---|---|---|---|---|
| Influenza admissions | All weeks | 135.28 | 170.26 | 20.5 [11.1, 26.5] |
|  | Outbreak | 161.24 | 226.44 | 28.8 [12.0, 34.0] |
|  | Non-outbreak | 122.74 | 145.86 | 15.8 [1.9, 22.6] |
| Influenza ED visits | All weeks | 1.120 | 1.496 | 25.1 [22.2, 27.8] |
|  | Outbreak | 1.420 | 1.960 | 27.5 [25.6, 29.4] |
|  | Non-outbreak | 0.970 | 1.267 | 23.5 [19.0, 27.7] |
| ILINet %ILI | All weeks | 0.936 | 1.208 | 22.5 [19.4, 24.8] |
|  | Outbreak | 1.022 | 1.337 | 23.5 [21.3, 25.6] |
|  | Non-outbreak | 0.881 | 1.132 | 22.2 [17.6, 25.5] |
| COVID-19 admissions | All weeks | 66.66 | 71.75 | 7.1 [5.3, 10.1] |
|  | Outbreak | 74.85 | 79.42 | 5.7 [3.0, 8.2] |
|  | Non-outbreak | 62.65 | 68.44 | 8.5 [5.0, 14.6] |
| COVID-19 ED visits | All weeks | 0.383 | 0.403 | 5.1 [−3.4, 7.3] |
|  | Outbreak | 0.365 | 0.389 | 6.2 [4.8, 7.5] |
|  | Non-outbreak | 0.387 | 0.406 | 4.5 [−6.5, 7.4] |
| RSV admissions | All weeks | 39.98 | 40.02 | 0.1 [−5.0, 5.6] |
|  | Outbreak | 44.25 | 37.38 | −18.4 [−29.6, −4.8] |
|  | Non-outbreak | 36.69 | 39.58 | 7.3 [2.0, 12.3] |
| RSV ED visits | All weeks | 0.157 | 0.167 | 6.0 [−2.6, 9.2] |
|  | Outbreak | 0.170 | 0.159 | −6.9 [−12.4, −3.1] |
|  | Non-outbreak | 0.150 | 0.171 | 12.8 [3.1, 16.5] |

**Table S7.** GLCast accuracy during own-only and shared respiratory upswings. Each target uses the same jurisdictions in both phases, with at least four original scored weeks per phase and observed outcomes for all three viruses in its endpoint family. Admission and ED outcomes use their own three-virus observations and labels. Shared weeks fall inside the target’s archived upswing and at least one other virus’s upswing; own-only weeks fall inside the target’s own upswing and outside both other viruses’ upswings. A, Within each jurisdiction and phase, amplitude-relative error is 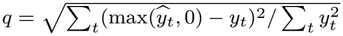 on the same cells. Phase means average *q* equally across jurisdictions. The contrast is the ratio of the shared and own-only means; values above one indicate larger relative error during shared upswings. B, Unnormalized RMSE and observed RMS amplitude are likewise calculated within jurisdiction and then averaged equally. Raw units are weekly admissions or ED percentage points. Brackets contain descriptive 95% paired-jurisdiction BCa intervals from 10,000 resamples, conditional on saved fits, recipe selection, retrospective labels, and eligibility.

| **A. Amplitude-relative GLCast error** |  |  |  |  |  |
|---|---|---|---|---|---|
| Target | Juris. | Cells, own/shared | Own-only *q* | Shared *q* | Shared/own [95% interval] |
|---|---|---|---|---|---|
| Influenza admissions | 40 | 346/1,092 | 0.437 | 0.521 | 1.194 [0.969, 1.409] |
| Influenza ED | 40 | 399/1,721 | 0.399 | 0.522 | 1.307 [1.212, 1.430] |
| COVID-19 admissions | 50 | 707/931 | 0.326 | 0.318 | 0.974 [0.868, 1.087] |
| COVID-19 ED | 44 | 778/681 | 0.247 | 0.256 | 1.036 [0.966, 1.123] |
| RSV admissions | 47 | 502/1,039 | 0.475 | 0.485 | 1.021 [0.701, 1.274] |
| RSV ED | 50 | 768/2,339 | 0.274 | 0.339 | 1.237 [1.135, 1.401] |
| **B. Raw GLCast error and observed amplitude** |  |  |  |  |  |
| Target | Own RMSE | Shared RMSE | Ratio [95% interval] | Observed RMS, own/shared |  |
| Influenza admissions | 206.08 | 136.95 | 0.665 [0.374, 0.926] | 504.38/263.37 |  |
| Influenza ED | 2.026 | 1.174 | 0.579 [0.483, 0.731] | 4.938/2.257 |  |
| COVID-19 admissions | 38.49 | 93.04 | 2.417 [2.011, 2.980] | 146.09/313.26 |  |
| COVID-19 ED | 0.218 | 0.464 | 2.126 [1.857, 2.457] | 0.883/1.838 |  |
| RSV admissions | 26.38 | 47.55 | 1.803 [1.202, 2.898] | 79.84/105.55 |  |
| RSV ED | 0.138 | 0.172 | 1.249 [1.056, 1.505] | 0.476/0.511 |  |

### A. Amplitude-relative GLCast error

**Inputs, lagged designs, and analysis configurations**

We first describe how surveillance histories and auxiliary streams form the predictors of a local model. Let *Y*_*ℓ,t*_ be the recorded surveillance value for location *ℓ* and target date *t*, and let 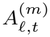 denote auxiliary stream *m*. For auxiliary lags K_*X*_ and target-history lags K_AR_, the predictor row is

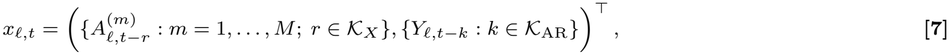

The response for this row is *Y*_*ℓ,t*_. An auxiliary lag of zero therefore refers to the date being estimated. All lags of one auxiliary stream form one group. Columns are ordered by stream and then lag, with all target-history columns forming the final group. Stream and lag identifiers allow the same features to be matched across locations even when their available streams differ. Table 1 gives the lag sets used in each analysis. For a given refit, let *X*_*ℓ*_ stack the complete predictor rows, *y*_*ℓ*_ contain their responses, *n*_*ℓ*_ be the number of rows, and *p*_*ℓ*_ be the number of columns. The disjoint column sets *ℐ*_*g*_ define the groups, with 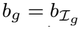.

We construct the design separately for each location. A stream absent from its input table is omitted, and a row with a missing response or required predictor is removed after lagging. We neither impute missing predictors with zero nor interpolate missing targets. Daily and weekly dates retain their original calendar anchoring; a weekly increment is seven elapsed days. Each daily auxiliary group contains several lags of one stream, whereas the main weekly groups each contain one column. Thus, splitting groups into single columns leaves the main ILI design unchanged.

The two classic targets and seven modern targets share one specification, denoted S3 in the analysis archive: the same numerical fitting routine, standardized penalty scale, adaptive group weighting, coordinate-penalty multiplier, candidate penalty grid, and geographic combiner. Each target has its own adaptive weights and penalty history. This common specification was chosen using the historical benchmarks and then held fixed. The lag sets describe the information included in each regression; they do not establish historical release-time availability. The retrospective timing and model-selection scope are defined in Appendix H.

## B. Standardization and penalty specification

### B.1. Fitting-window standardization and raw coefficients

Standardization puts the predictors and response on a common scale before regularization. For each location and refit, let 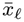 and 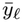 be their means over the complete fitting rows, and let 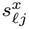 and 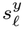 be their standard deviations. Standard deviations use divisor *n*_*ℓ*_, with a zero value replaced by one. Defining 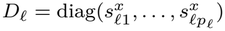, we fit the standardized variables

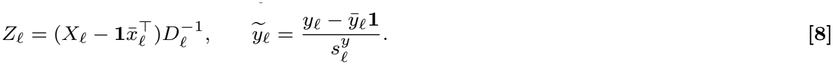

Centering absorbs the intercept. After fitting 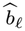, we return to the original measurement scale through

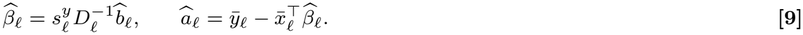

The local prediction is then 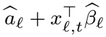. A constant fitting column is centered to zero and receives coefficient zero. These recovered raw coefficients and intercepts are also the quantities used when a source model is applied to another location (Appendix D).

### B.2 Common adaptive group and coordinate penalties

The local criterion in Eq. 1 combines a group penalty, which selects streams, with a coordinate penalty, which acts on their individual lags. All nine targets use

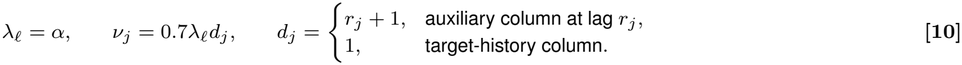

Here *α* directly penalizes standardized coefficients; neither penalty is divided by the response standard deviation. The coordinate penalty applies to target-history columns as well as auxiliary columns. Its lag factor gives longer auxiliary lags a larger penalty. At lag zero the penalty remains positive, even though there are no differences between auxiliary lags to penalize. Fitting rows have equal weight; the lag penalty is separate from the time-decay weights used by the autoregressive comparators.

We use a first pass to identify streams that are repeatedly selected. For each target, this pilot uses its main lag sets and origin schedule, uniform group weights, and no coordinate penalty. It selects its own penalties by the automatic rule defined below. Let 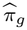 be the fraction of available origin–location pilot fits in which named group *g* has at least one nonzero coefficient. The adaptive weight is

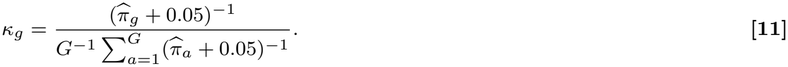

The average in the denominator includes all *G* requested groups, including autoregression, so the weights have mean one. Frequently selected groups receive less shrinkage in the final fit. A group absent from the available pilot fits has frequency zero; unavailable origin–location fits are not counted as observations. We align these weights to each location’s columns by stream name.

The pilot frequencies pool the full analysis period. The resulting weights remain fixed in the final primary and monthly fallback fits, while the final penalties are selected again using the adaptive, coordinate-penalized models. Pilot and final penalty histories can therefore differ. This use of selection frequency follows the motivation of adaptive regularization (70); its analysis-wide estimation is part of the retrospective design.

### B.3 Automatic penalty selection for every target

We choose the amount of regularization separately at each origin, using the performance of local models from earlier origins. Both the pilot and final fits use the same 19-value grid for all nine targets:

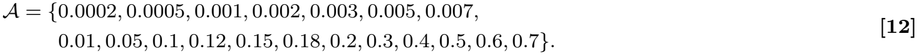

At origin *o*, one *α* is shared across locations for that target. Let *ℰ*_*b,α*_ contain the available test rows from earlier origin *b* at candidate *α*. We select

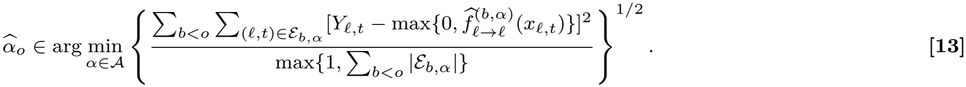

This criterion pools squared errors and row counts across locations and earlier origins, then takes their root mean square. Predictions are floored at zero before scoring. Each candidate uses its available rows, and ties are resolved in increasing *α* order. A zero accumulated count is replaced by one as shown in the denominator. No penalty is served before a usable local prediction is available; if the first usable fit has no earlier errors, the zero-history scores choose the smallest available grid value. The first scheduled origin is always treated as burn-in.

Primary and fallback schedules each use their own complete earlier-origin history. Here “earlier” refers to the fitting origin: its test window can extend beyond the origin at which the next penalty is selected. Thus, the criterion uses historical prediction errors but is not restricted to outcomes released by the current origin. This distinction, together with the analysis-wide pilot weights, determines the interpretation of the evaluation in Appendix H.

## C. Numerical fitting and screening

We reduce the fitting problem by starting with a subset of groups and adding omitted groups when they remain associated with the residual. The same routine is used for all nine targets. Below we describe the coefficient updates, the initial screen, and the residual check that expands this working set.

### Coefficient updates

For a working standardized design *Z* with *n* rows, the accelerated routine uses step size

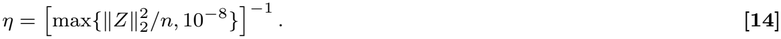

Starting from zero coefficients, let *z* be the current accelerated-gradient extrapolate and form 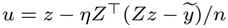. The update first shrinks each group and then each coordinate:

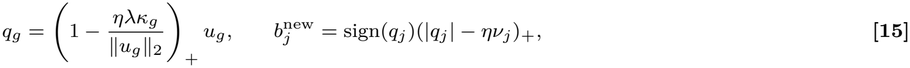

A zero group remains zero, and the coordinate operation is omitted when *ν*= 0. Acceleration starts at *a*_0_ = 1 and uses 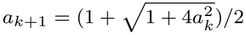 with extrapolation factor (*a*_*k*_ −1)*/a*_*k*+1_. Iteration stops when the coefficient change is at most 10^*−*6^ max {1, ∥*b*∥ _2_ }or after 3000 steps.

Every main fit has a positive coordinate penalty and uses this accelerated routine. Group-only pilot fits and zero-coordinate-penalty ablations instead use `celer.GroupLasso`, when available, with tolerance 10^*−*4^ and no additional intercept; the accelerated routine is its fallback. For multicolumn groups, the group-then-coordinate update in Eq. 15 is not asserted to equal the exact proximal map of the combined penalty. The numerical procedure above, rather than an exact sparse-group optimization claim, defines the local fits used in this study.

### Initial group screen

The routine represents groups with a common width *d*. A shorter trailing autoregressive group is padded with zero columns, whose coefficients are removed before prediction. To rank the groups initially, we summarize their row-wise magnitudes as

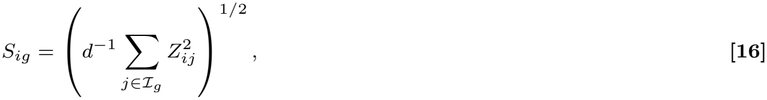

and compute the absolute correlation of each *S*_*·g*_ with 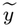. The initial working set contains the highest-ranked

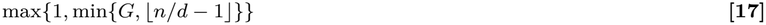

groups. Because the summary uses squared predictor values, it measures group magnitude without retaining individual predictor signs. It is a heuristic starting point for fitting (75, 76).

### Residual check and expansion

After a restricted fit, let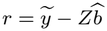. We add each omitted group satisfying

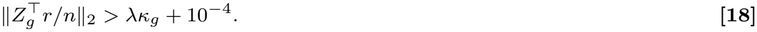

and refit the expanded set. This process ends when no omitted group satisfies the rule or after 50 rounds. For comparison, the exact zero-group Karush–Kuhn–Tucker (KKT) condition for the ideal sparse-group criterion would be

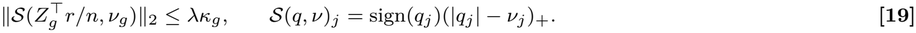

Soft-thresholding cannot increase the Euclidean norm, so the plain-norm check can add extra groups beyond those violating the thresholded condition. It provides a conservative expansion rule for omitted groups. It does not check active-group stationarity or alter the order of the numerical updates; termination therefore supplies no exact-optimizer certificate.

## D. Source screening and raw-coefficient transport

Geographic borrowing begins by finding locations with similar predictor–response associations. For each fitted location, we calculate the standardized cross-moments

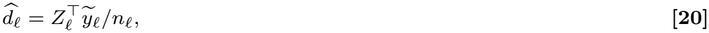

and index them by feature name. For target location 0 and source *ℓ*, let *J*_*ℓ*0_ be their common feature identifiers and 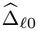 the source–target difference on these coordinates. Constant features remain in this alignment with their standardized zero values. We score each source by

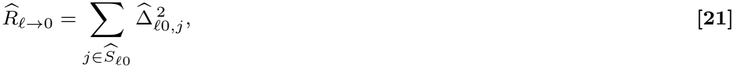

where 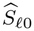 contains the min{50, |*J*_*ℓ*0_|} largest absolute discrepancies. A source with no shared coordinates receives infinite score. Up to 20 lowest-scoring sources form the candidate library, with source order resolving ties. This first screen uses the fitting-window associations to reduce the number of models considered for borrowing.

We then evaluate each candidate on the target location. The source’s raw coefficients from Eq. 9 are matched to the target feature names, an unmatched target coefficient is set to zero, and the source intercept is retained. Equation 2 applies these coefficients to the target’s raw predictor values. This operation uses the source’s fitted scale and intercept directly; target-specific centering or rescaling is not applied to the transported prediction.

The local candidate must provide finite predictions on all calibration and prediction rows. A source is likewise excluded if either prediction column has a nonfinite value. Among the remaining library sources, we keep up to five with the lowest target-calibration RMSE, resolving ties in screened order. The local model is always the first column, and all candidates use the selected common *α*. This second step ranks sources by their predictive performance on the target. Availability is checked from predictions, although the historical complete-case design itself also uses observed outcomes to determine which rows are retained.

## E. Stacking and Q-inspired aggregation

We combine the target’s own model with the retained sources using two complementary weight calculations. Stacking fits the target’s calibration outcomes while shrinking donor weights; a Q-inspired calculation assigns normalized weights according to the candidate predictions. The final model blends these two weight vectors.

### E.0.0.1 Nonnegative stacking

Let *F* ∈ ℝ^*m×J*^ contain the candidates’ raw calibration predictions, with the local model in column 1, and let *y*^v^ contain the *m* calibration outcomes. The stacking objective in Eq. 3 is solved by nonnegative least squares on

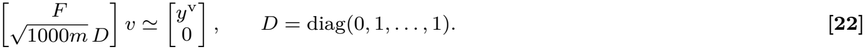

The diagonal matrix leaves the local-model weight unpenalized and applies ridge strength 1000 to the source weights. There is no extra intercept or sum-to-one constraint. Stacking can therefore recalibrate the local model as well as borrow from other locations.

### E.0.0.2. Q-inspired weights

To calculate the second weight vector, divide the calibration outcomes and candidate predictions by the outcome standard deviation *s*_v_, using divisor *m* and replacing a zero standard deviation by one. Write *F* 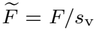 and 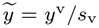 ; these quantities are scaled without centering. With temperature *τ* = 0.25, initialize

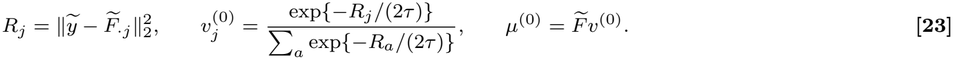

The uniform-prior calculation then makes at most ten updates:

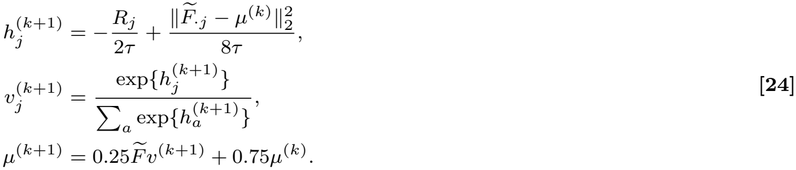

Before exponentiation, we subtract the largest log weight for numerical stability. Iteration can end earlier when ∥ *v*^(*k*+1)^ −*v*^(*k*)^ ∥ _1_ *<* 10^*−*3^. The returned 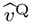 is the last weight vector; *μ*^(*k*)^ is only the damped running prediction used in the update. With a single candidate, its weight is one. The approach is motivated by Q-aggregation (71, 72, 77), but this finite iteration is the procedure evaluated here. We do not substitute the exact minimizer of a loss-plus-entropy criterion or invoke its oracle guarantees.

### E.0.0.3. Blending the two predictions

Equation 4 combines the stacking and Q-inspired vectors with *γ* = *m/*(*m* + 10). Thus, stacking receives more weight when more calibration rows are available. With complete calibration windows, *m* = 30 gives *γ* = 0.75 for classic COVID-19, while *m* = 4 gives *γ* = 2*/*7 for the weekly analyses. Missing rows can reduce *m*. The library size 20, donor cap five, ridge strength 1000, blending constant 10, and Q temperature 0.25 are fixed across analyses.

For candidate identities *c*_*j*_, the aggregate can equivalently be written

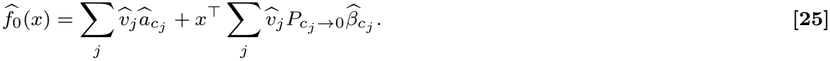

Both the coefficients and intercept are weighted sums of their local and source counterparts. Predictions use the raw nonnegative blended weights, whose sum need not be one. For visualization we divide each weight by the total, obtaining the share *v*_*j*_*/* _*a*_ *v*_*a*_; these display shares do not replace the prediction weights. Candidate predictions may be negative, so the zero floor is applied after combination for scoring and plotting.

## F. Interpretation and deterministic properties

The following properties explain what the source screen and aggregation steps measure. They are algebraic consequences of the stated calculations. Group penalties motivate joint selection of related columns (68, 69); the numerical routine and finite Q-inspired updates remain those in Appendices C and E.

### F.0.0.1. 1. Source screening compares predictive associations

For any coefficient vector *b*_*ℓ*_, write the standardized response as 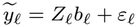, with *ε*_*ℓ*_ denoting its residual. Substituting this representation into Eq. 20 gives

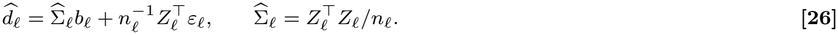

This identity separates coefficient effects, predictor covariance, and residual variation. Any of these can change the cross-moment score between locations. The score therefore measures similarity of marginal predictive associations, not transmission between locations or target-specific excess risk. It does not by itself protect against negative transfer (71).

### F.0.0.2. 2. Squared loss of the blend is bounded by the average parent loss

For fixed *F* and *y*, any weight vectors *v*^S^ and *v*^Q^, and 0 ≤ *γ* ≤ 1,

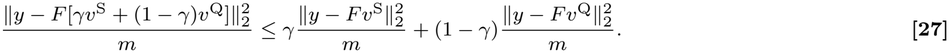

*Proof*. The prediction residual of the blend is the same convex combination of the two parent residuals. Applying convexity of the squared Euclidean norm gives the inequality. The bound holds on the fixed design regardless of how the parents were computed. It compares the blend with their weighted average loss, rather than with the better parent or an unseen-data risk.

### F.0.0.3. 3. The stacking ridge limits donor weight

Let 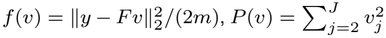, and let *v*_*λ*_ solve min_*v≥*0_{*f* (*v*) + *λP* (*v*)*/*2} exactly. Increasing *λ* cannot increase *P* (*v*_*λ*_) or decrease the corresponding residual loss *f* (*v*_*λ*_). Moreover,

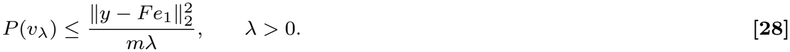

*Proof*. For *λ*^*′*^ *< λ*^*′′*^, add the two optimality inequalities obtained by comparing each solution with the other. This gives the ordering of *P* ; substituting it into either inequality gives the ordering of *f* . Comparing *v*_*λ*_ with the feasible self-only vector *e*_1_ = (1, 0, …, 0)^*⊤*^, whose penalty is zero, gives the displayed bound. Thus donor weights approach zero as the ridge increases without bound, while the unpenalized self weight remains free to recalibrate the local model. These are properties of the stacking optimization problem, not accuracy guarantees for the final blend.

### F.0.0.4. 4. Prediction weights and display shares have different roles

Each Q update is a probability vector, and stacking produces nonnegative weights. Their blend is therefore nonnegative, with total weight 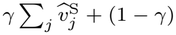. This total generally differs from one. The resulting prediction is nonnegative when all candidate predictions are nonnegative; otherwise it still requires the downstream floor. If the calibration-response variance is positive, multiplying all outcomes and candidate predictions by the same positive constant leaves the normalized Q update unchanged. The fixed raw-scale stacking ridge prevents this invariance from extending generally to the full blend. These observations explain why normalized display shares summarize borrowing without defining the prediction itself.

These properties do not establish a best-candidate risk bound for the surveillance analyses. Such a result would require assumptions about dependence, data availability, and selection that are not supplied by the retrospective evaluation. In particular, calibration overlaps fitting, and the pilot weights, penalty histories, and common specification use historical information as described in Appendix H.

## G. Target-only comparators

We use ten comparators to separate the value of auxiliary information and learned model borrowing from simpler uses of target history. They comprise one local autoregression, five classical univariate methods, and four geographically pooled autoregressions. Their prediction panels are fixed across the reported GLCast comparisons. All methods are scored after predictions are floored at zero.

### Local autoregression

This method regresses the target on its lagged values, giving recent training rows more weight. A row at elapsed distance Δ_*i*_ from the fitting-window end receives weight exp(−*η*Δ_*i*_), where elapsed time is measured in days or weeks for the corresponding analysis. The decay rate *η* ∈ {0, 0.01, 0.05, 0.1} is selected by calibration mean squared error. A 10^*−*8^ ridge provides numerical stabilization (39, 62, 78).

### Classical univariate methods

Persistence carries forward the latest available target value. Seasonal naive uses the available value at the same seasonal phase, defined by weekday for daily data and ISO week for weekly data. Climatology uses the expanding mean at that phase. ETS fits an additive-error damped-trend model and adds seasonality after two complete seasonal periods; it falls back to seasonal naive and then persistence when needed (79). ARIMA selects autoregressive and moving-average orders from {0, 1, 2} and differencing order from {0, 1} by the Akaike information criterion, using persistence if the fitting candidates fail (80).

### Geographic autoregressions

We cross two pools, land-adjacent locations and all other locations, with two borrowing strategies. Shared-coefficient models stack observations across the pool and fit common autoregressive coefficients with location-specific intercepts. Spatial-lag models retain a separate target-location regression and add the leave-one-out pool mean at the local target lags. These four models borrow surveillance histories without auxiliary streams or a learned model-aggregation network (81, 82). The name *Pooled AR all shared* identifies the shared-coefficient model using all locations. It is the classic reference and the labeled reference in selected modern supplements. The main modern comparisons instead use each target’s lowest-error overall comparator.

## H. Historical windows, scoring, and secondary analyses

### Window construction and availability scope

We refit models at successive origins and evaluate the predictions assigned to the retained target dates. For origin *O*, let *W, V*, and *H* be the nominal fitting, calibration, and test lengths. The date sets are

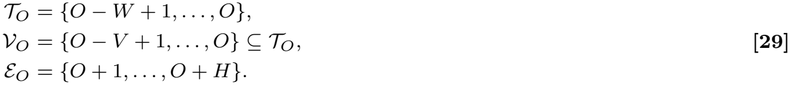

The arithmetic is in days or weeks. After lagging and complete-case filtering, an origin requires at least ten fitting rows, two calibration rows, and two test rows. The first scheduled origin is burn-in. Later fits supply the primary offsets in Table 1, with the classic fallback assignments described below. Table S8 lists the schedules.

**Table S8.** Primary refit schedules and retained scoring masks. The first scheduled origin is burn-in. Post-burn-in origins are potential refits and need not contribute to a retained scoring mask. Modern target-specific start dates and missingness are reported with their target coverage.

| Analysis | First scheduled origin | Last scheduled origin | Schedule | Later origins |
|---|---|---|---|---|
| Classic ILI | 15 Jan 2012 | 26 Nov 2017 | Weekly | 306 |
| Classic COVID-19 | 26 Mar 2021 | 22 Mar 2024 | Every 7 days | 156 |
| Modern, seven targets | 21 Jan 2023 | 25 Jul 2026 | Weekly | 183 |

### H.2 Metrics and interval estimates

We first measure prediction error within each jurisdiction, then give jurisdictions equal weight in the summary. For method *a* and jurisdiction *ℓ*, let *C*_*aℓ*_ be its available scored cells in the all-period, outbreak, or non-outbreak stratum. Its RMSE is

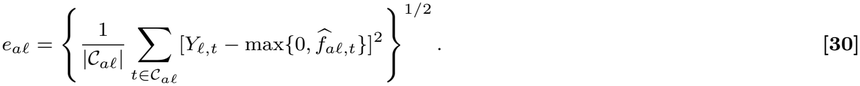

The aggregate is the unweighted mean *e* over jurisdictions with a defined error. A percentage reduction relative to reference *r* is 100(1 *e /e* ), computed from these mean errors. MAE and *R*^2^ are also calculated within jurisdictions and then averaged. For *R*^2^, the denominator is the within-jurisdiction, within-stratum sum of squared deviations from its own outcome mean; zero denominators are missing.

We summarize between-jurisdiction variation with descriptive 95% bias-corrected and accelerated (BCa) bootstrap intervals (73). Each of 10,000 draws resamples whole jurisdictions with replacement, leaving their saved scores and time series intact. For paired comparisons, the same draw supplies both methods. Acceleration is calculated by deleting one jurisdiction at a time. Calculations use seed 7; the additional modern ratio calculations use the separate generators specified below. A paired mean-difference interval is on the original RMSE scale unless explicitly labeled as a percentage interval.

Relative RMSE, *e*_*a*_*/e*_*r*_, permits comparison across modern outcomes with different measurement units; one denotes equal mean error. The supplementary full-roster heatmap uses Pooled AR all shared. The main modern figure and Table S6 use each target’s lowest-error overall comparator, held fixed across phases.

**Fig. S1.**
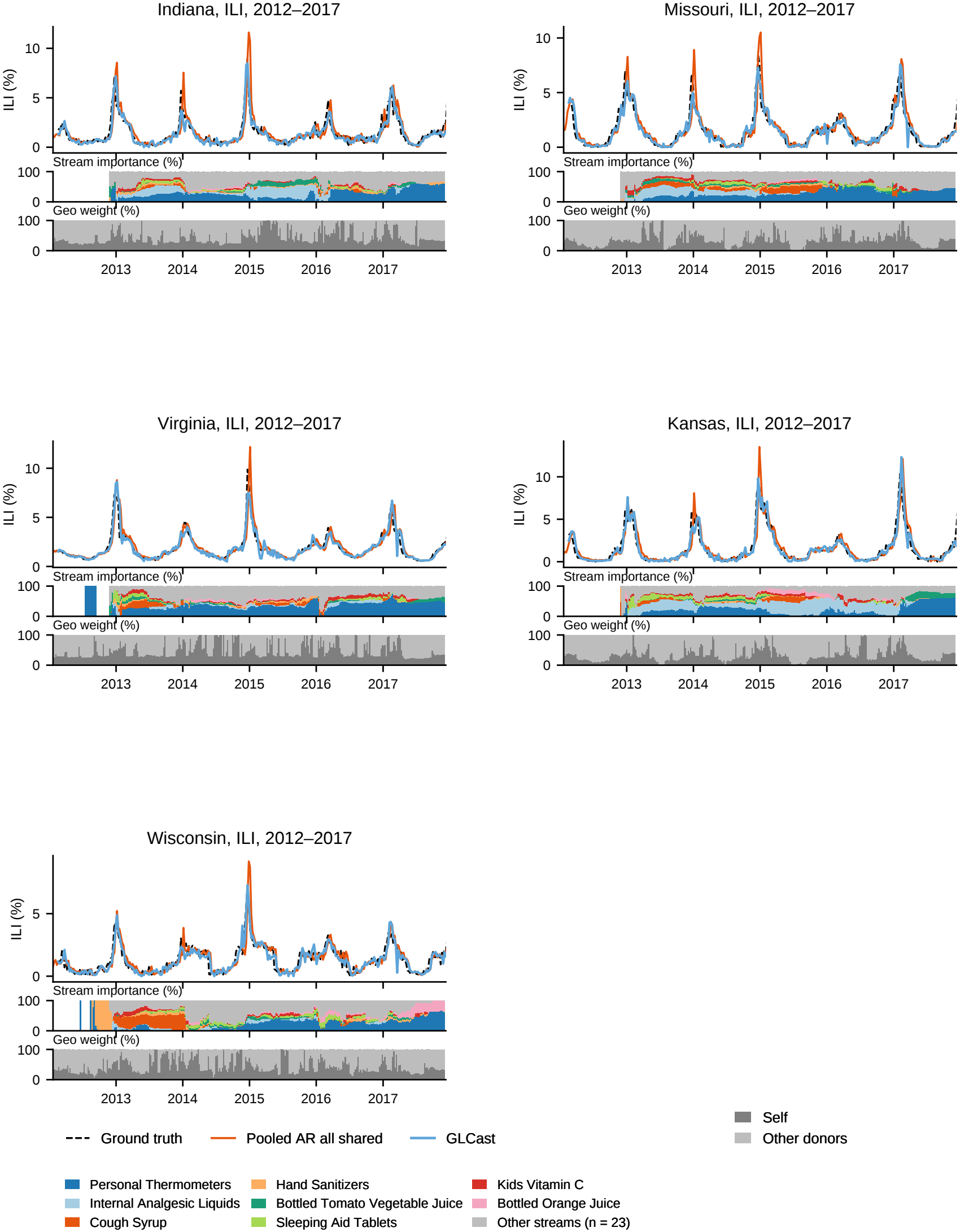
Five ILI states: nowcasts and fitted information shares. Indiana, Missouri, Virginia, Kansas, and Wisconsin are retained illustrative examples. In each cell, the upper trace shows observed weekly %ILI against GLCast and Pooled AR all shared nowcasts, with predictions floored at zero. The middle strip shows state-specific auxiliary-importance shares, using the same eight streams with the largest national mean importance in every cell and pooling the remaining 23. The lower strip shows normalized shares on the state’s own model and other donors; predictions use raw blended weights. Legends are shared across cells. These descriptive traces and fit diagnostics extend beyond the retained scoring mask.

**Fig. S2.**
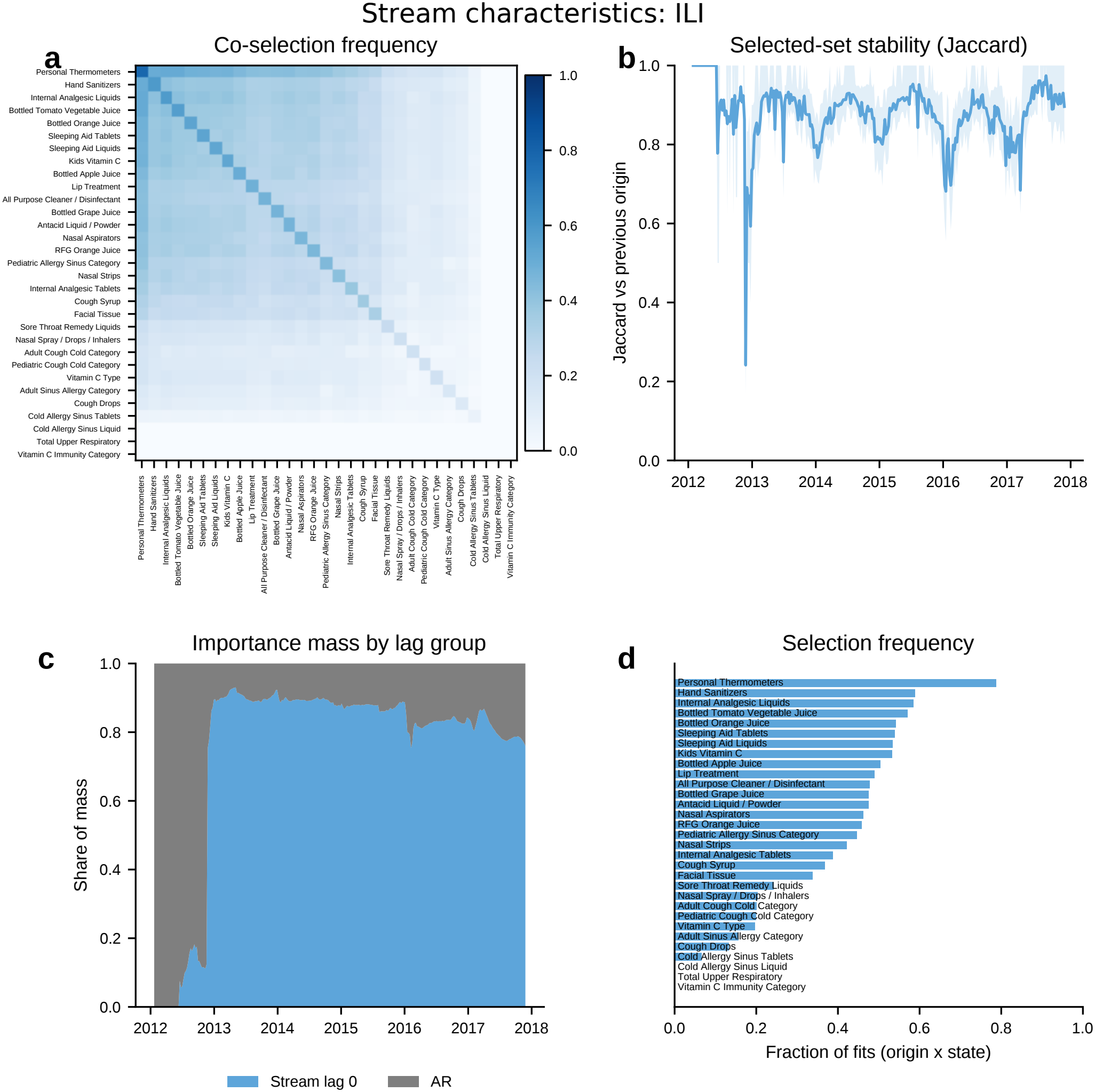
How GLCast uses the 31 retail streams over time (ILI). Diagnostics over 13,770 retained state-origin fits (306 origins *×* 45 states). (**A**) Stream co-selection frequency; the diagonal gives each stream’s selection frequency. (**B**) Jaccard similarity between successive nonempty selected-feature sets within a state; the line is the across-state mean and the band is the interquartile range. The mean over available state-origin pairs is 0.880; 16 entirely empty coefficient sets are skipped. (**C**) Per-origin shares of total coefficient mass, defined as absolute coefficient magnitude times rolling training-window standard deviation. Mean shares are 74.6% contemporaneous auxiliary and 25.4% autoregressive. **(D)**Stream selection frequencies. Overall, 10.1% of fits select no auxiliary stream; the median fit contains 13 auxiliary streams and 14 total features. Personal Thermometers, Hand Sanitizers, and Internal Analgesic Liquids are selected in 78.8%, 58.8%, and 58.5% of fits. These are local-fit diagnostics, including origins outside the scoring mask; the Jaccard band describes variation across states.

**Fig. S3.**
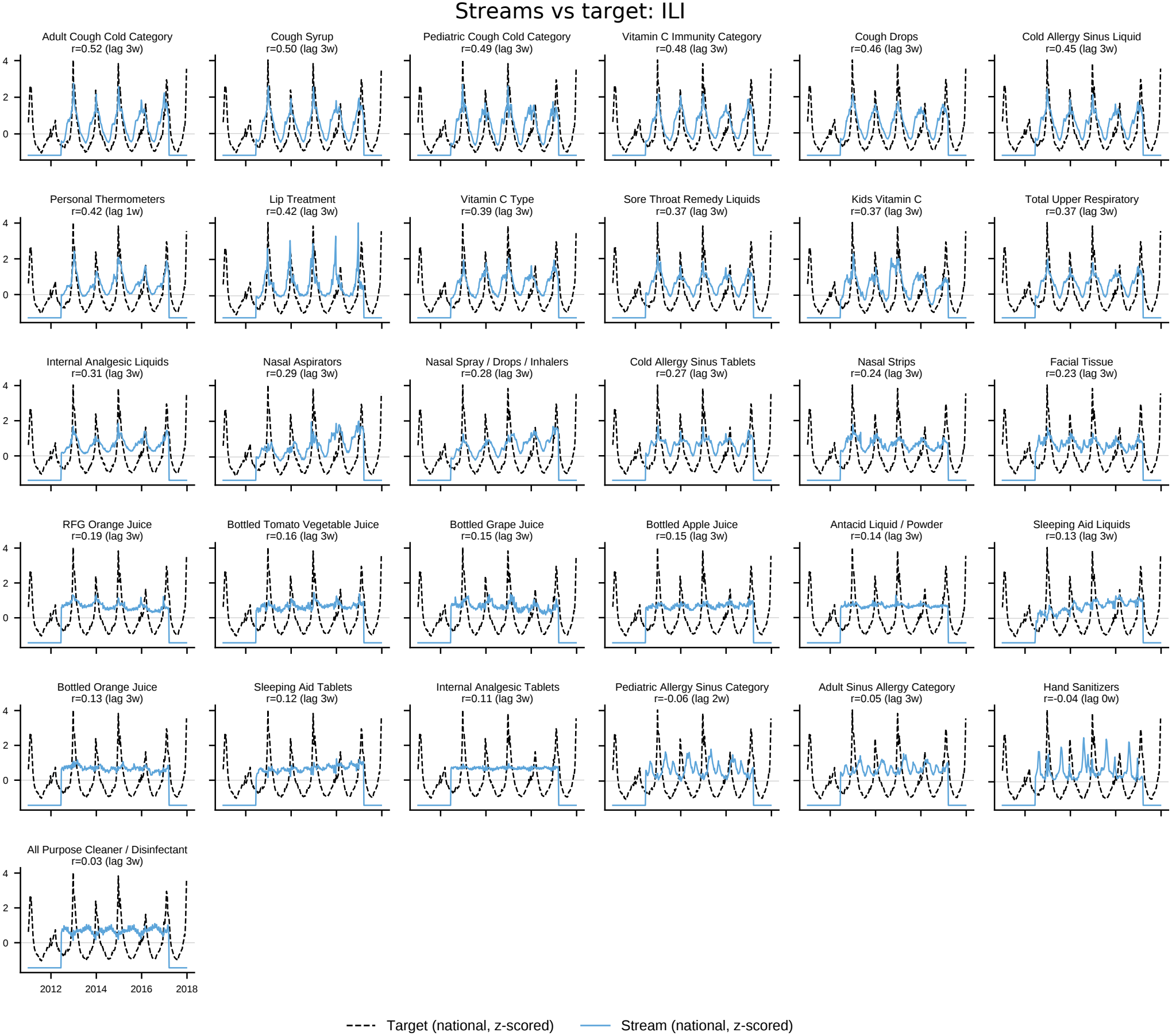
Retail streams against the influenza target. Each mini-panel overlays a nationally aggregated retail stream (blue) on the national %ILI target (black dashed), both *z*-scored over the full input period, January 2011–December 2017. The national target is the unweighted mean across the 45 evaluated states. Curves are plotted at their unshifted dates; panels are ordered by maximum absolute Pearson correlation over lags of 0–3 weeks, and each title reports the maximizing correlation and lag (positive lag: the stream leads the target). The strongest alignments are Adult Cough Cold Category (*r* = 0.52, three-week lead), Cough Syrup (0.50, three weeks), and Pediatric Cough Cold Category (0.49, three weeks). Flat edge plateaus are constant-fill periods in the supplied inputs. These are descriptive input summaries, not measures of held-out prediction accuracy.

**Fig. S4.**
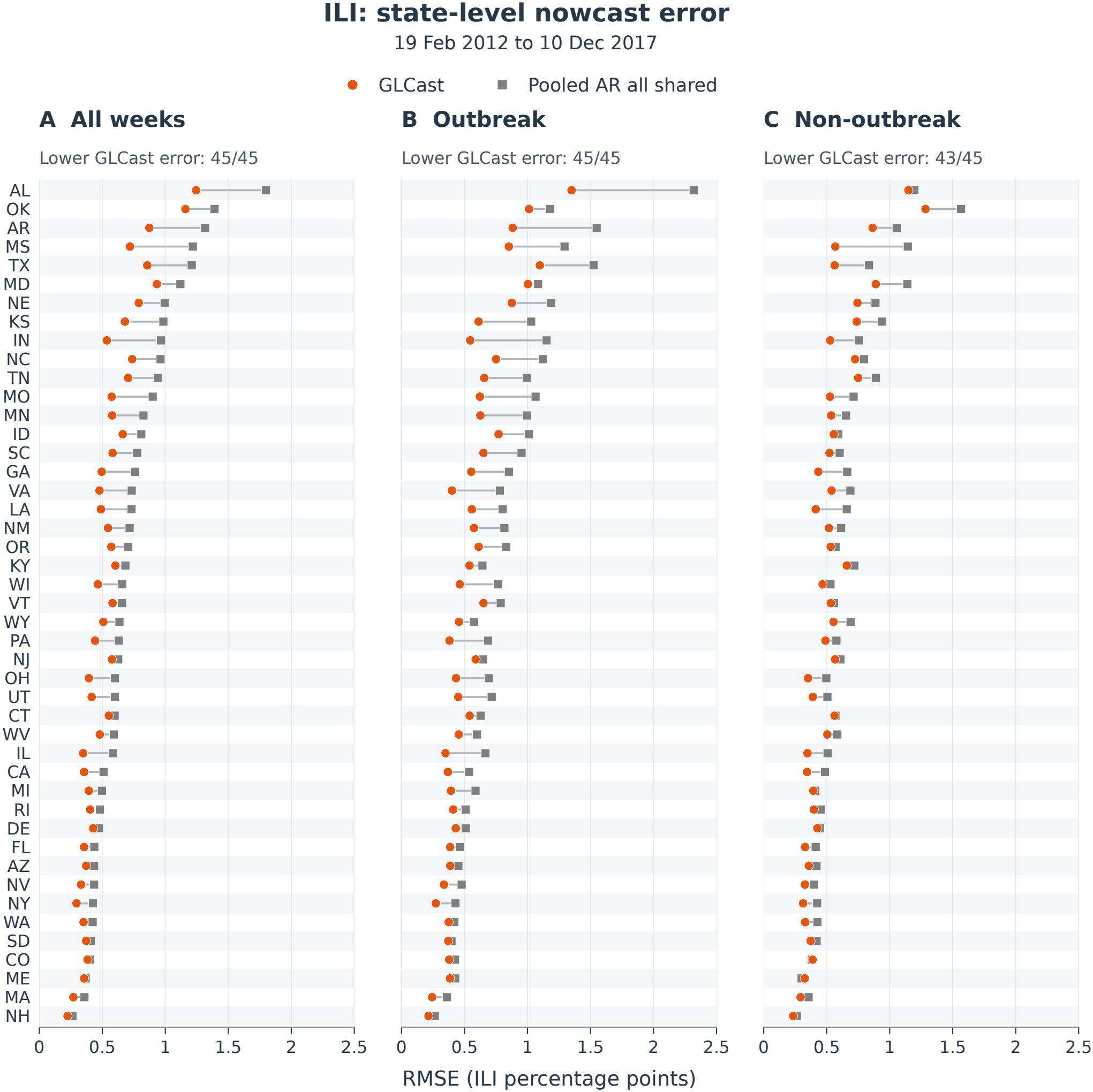
State-level ILI prediction error. Connected orange circles and gray squares show GLCast and the fixed Pooled AR all shared comparator for each of 45 states over (**A**) all scored weeks, (**B**) outbreak weeks, and (**C**) non-outbreak weeks. RMSE is measured in ILI percentage points, using the original 13,680 scored state-weeks from February 19, 2012, through December 10, 2017, and predictions floored at zero. States are ordered by decreasing comparator RMSE over all weeks, with that order retained across panels; labels are postal abbreviations. GLCast has lower RMSE in 45, 45, and 43 states, respectively. Both methods cover identical cells in every state and stratum. The points are individual-state error estimates.

**Fig. S5.**
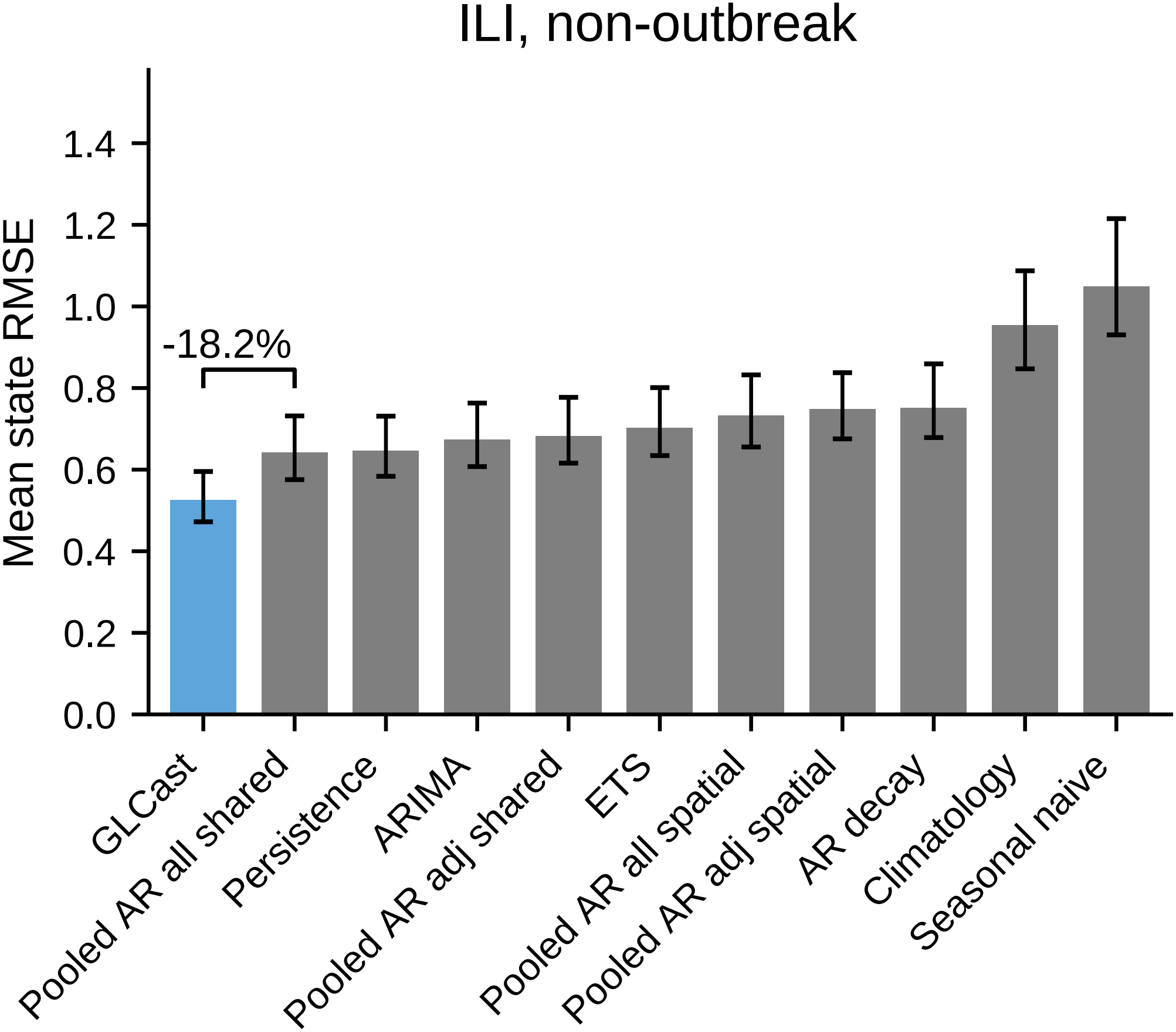
Non-outbreak ILI performance of the full comparator roster. Equal-state mean RMSE across 45 states for GLCast and ten comparators over non-outbreak weeks. Error bars are descriptive 95% BCa intervals from 10,000 state resamples, conditional on the benchmark-selected specification. The bracket compares GLCast with Pooled AR all shared: 0.525 versus 0.642 percentage points of ILI, an 18.2% reduction (paired absolute difference 0.117; interval [0.092, 0.155]). The annotation expresses signed percentage change, with a negative value favoring GLCast.

**Fig. S6.**
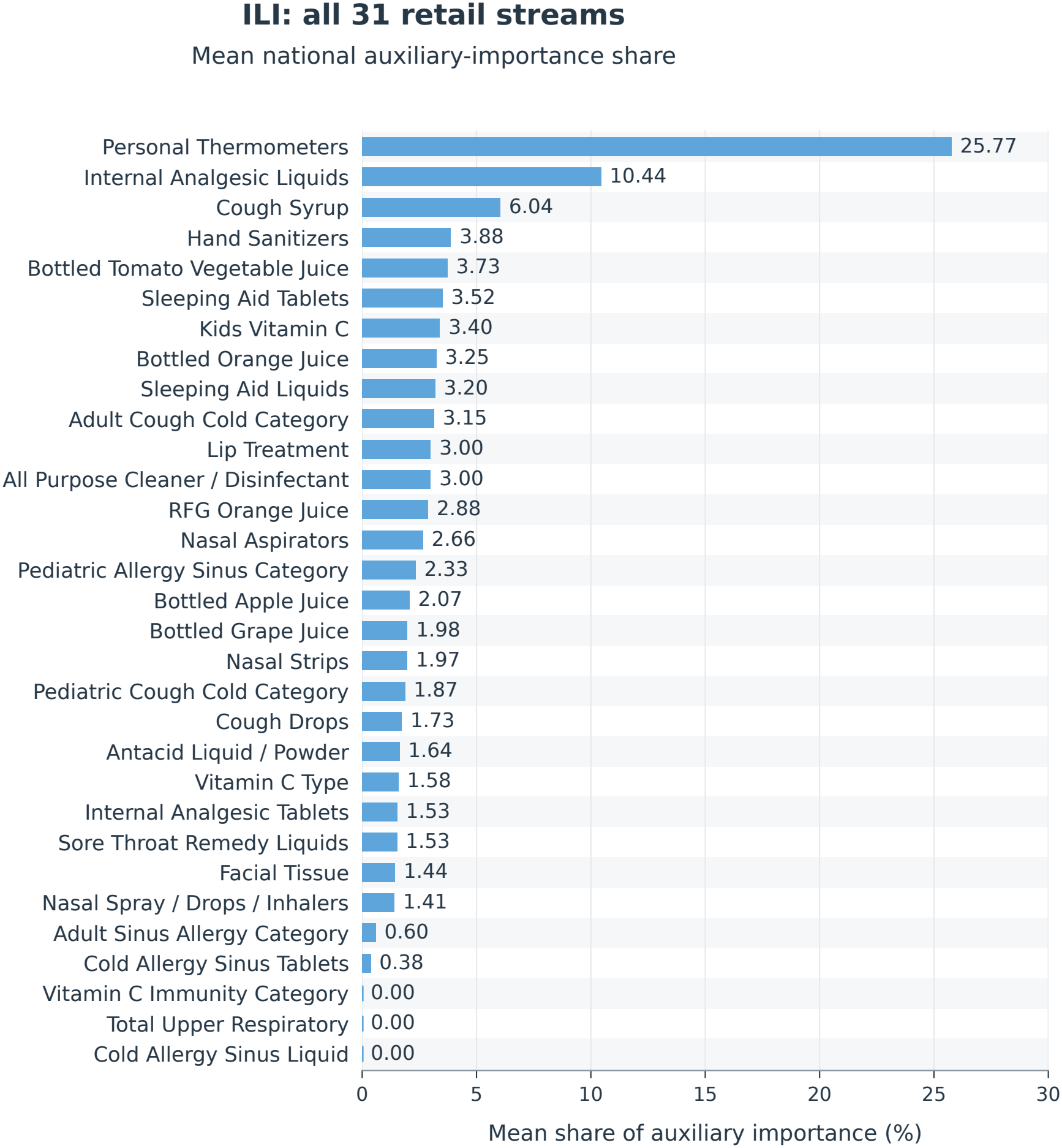
Mean auxiliary importance across all 31 retail streams. For each retained local-fitting origin, absolute raw coefficients multiplied by the corresponding rolling 55-week predictor standard deviations are summed across states and lags, then normalized across auxiliary streams. Bars average these national shares equally over the 285 origins with nonzero auxiliary mass; 21 zero-total origins are excluded from the 306 retained origins spanning January 22, 2012, through November 26, 2017. The target-history group is excluded from this denominator. All streams are retained, including the three with exactly zero recorded mass. These are local-fit coefficient summaries, distinct from total coefficient-mass shares and from attribution of the final geographic ensemble. Retained diagnostic origins need not serve scored cells. Values are rounded to two decimal places; no uncertainty intervals are estimated.

**Fig. S7.**
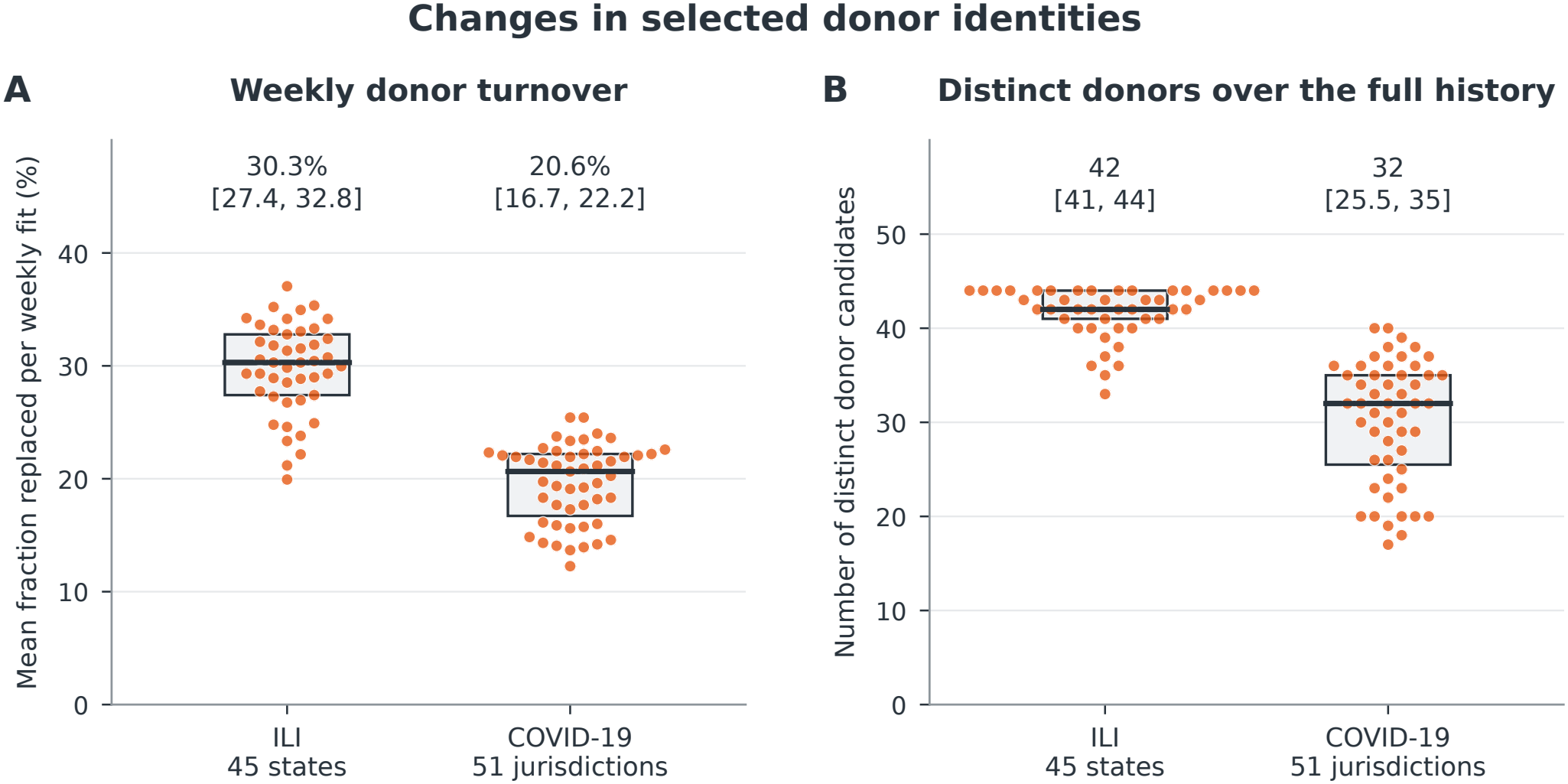
Changes in selected donor identities across all classic jurisdictions. (**A**) Each point is a jurisdiction’s mean percentage of the five retained donor candidates replaced between successive weekly fitting origins. Replacement is 100(1 *−* |*D*_*o*_ *∩ D*_*o−*7_|*/*5), where *D*_*o*_ excludes the local model. (**B**) Number of different donor candidates retained at least once over the full recorded history. All 45 ILI states and 51 COVID-19 jurisdictions are shown. Boxes span the empirical interquartile range, horizontal bars mark medians, and annotations give median [interquartile range]; horizontal displacement separates points without changing their values. ILI uses 306 weekly origins per state from January 22, 2012, to November 26, 2017; COVID-19 uses 156 from April 2, 2021, to March 22, 2024. There are 13,725 and 7,905 exact seven-day transitions, respectively, with no gaps. These are retained-fit diagnostics; monthly fallback fits are excluded. Candidate membership is recorded irrespective of final weight and does not measure material predictive influence. Distinct-donor totals depend on the different history lengths and donor pools (44 versus 50 possible donors). The displayed ranges describe jurisdictional variation, not confidence intervals.

**Fig. S8.**
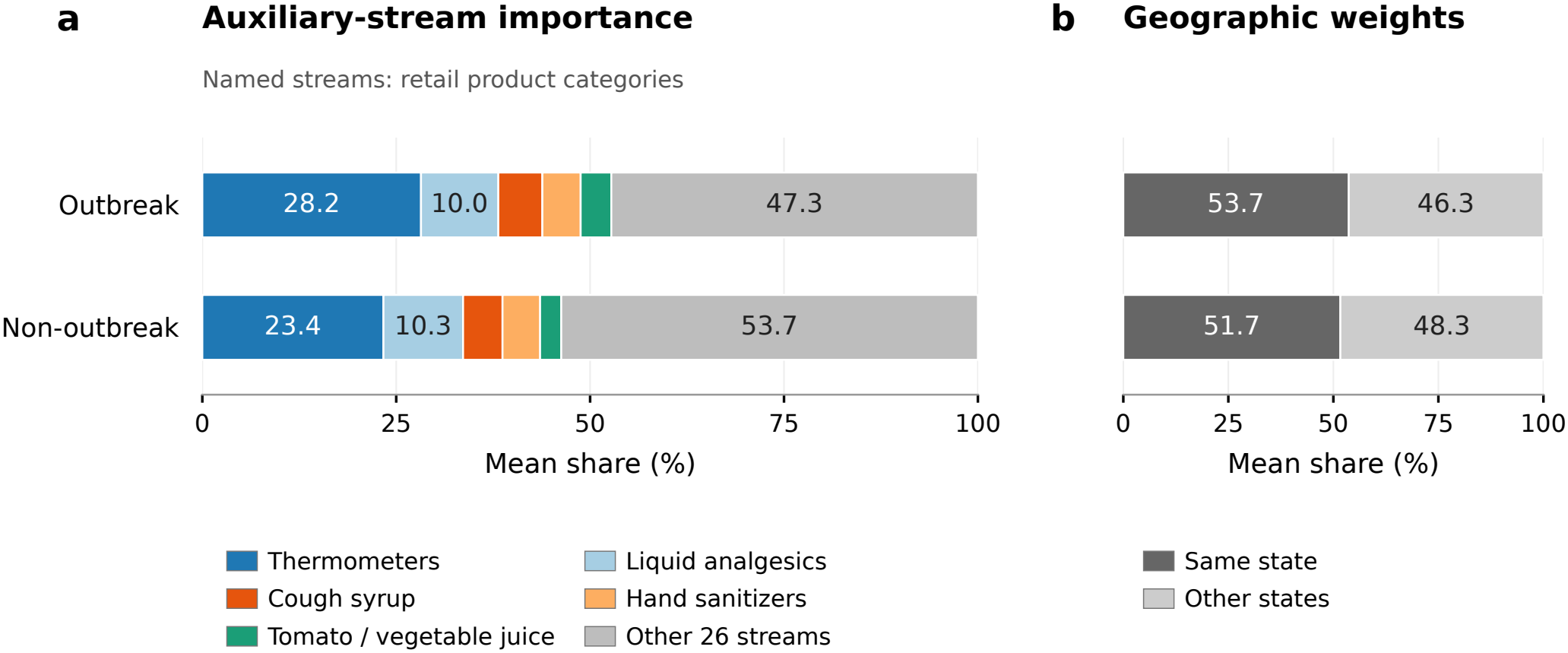
Stream and geographic importance during and outside classic ILI outbreaks. (**A**) Mean auxiliary-stream shares for the five leading streams, selected once from the all-period summary, and the remaining 26 streams. Local shares use absolute coefficient magnitude times the predictor’s rolling training-window standard deviation, summed over lags and normalized across auxiliary streams. Defined local shares are averaged equally across states within each scored week and phase, then equally across defined weeks. These are averages of local compositions, rather than pooled raw coefficient masses. (**B**) Same-state and other-state shares of the serving models’ nonnegative geographic weights, normalized within each model and averaged in the same order. The analysis uses all 13,680 scored state-weeks across 45 states. Panel A includes 6,020 outbreak and 6,404 non-outbreak state-weeks with positive auxiliary mass; 1,256 state-weeks with undefined auxiliary compositions are omitted from A but retain defined geographic weights in B. Archived outbreak labels are assigned to scored target dates. Bar lengths use unrounded descriptive means; no confidence intervals are estimated.

**Fig. S9.**
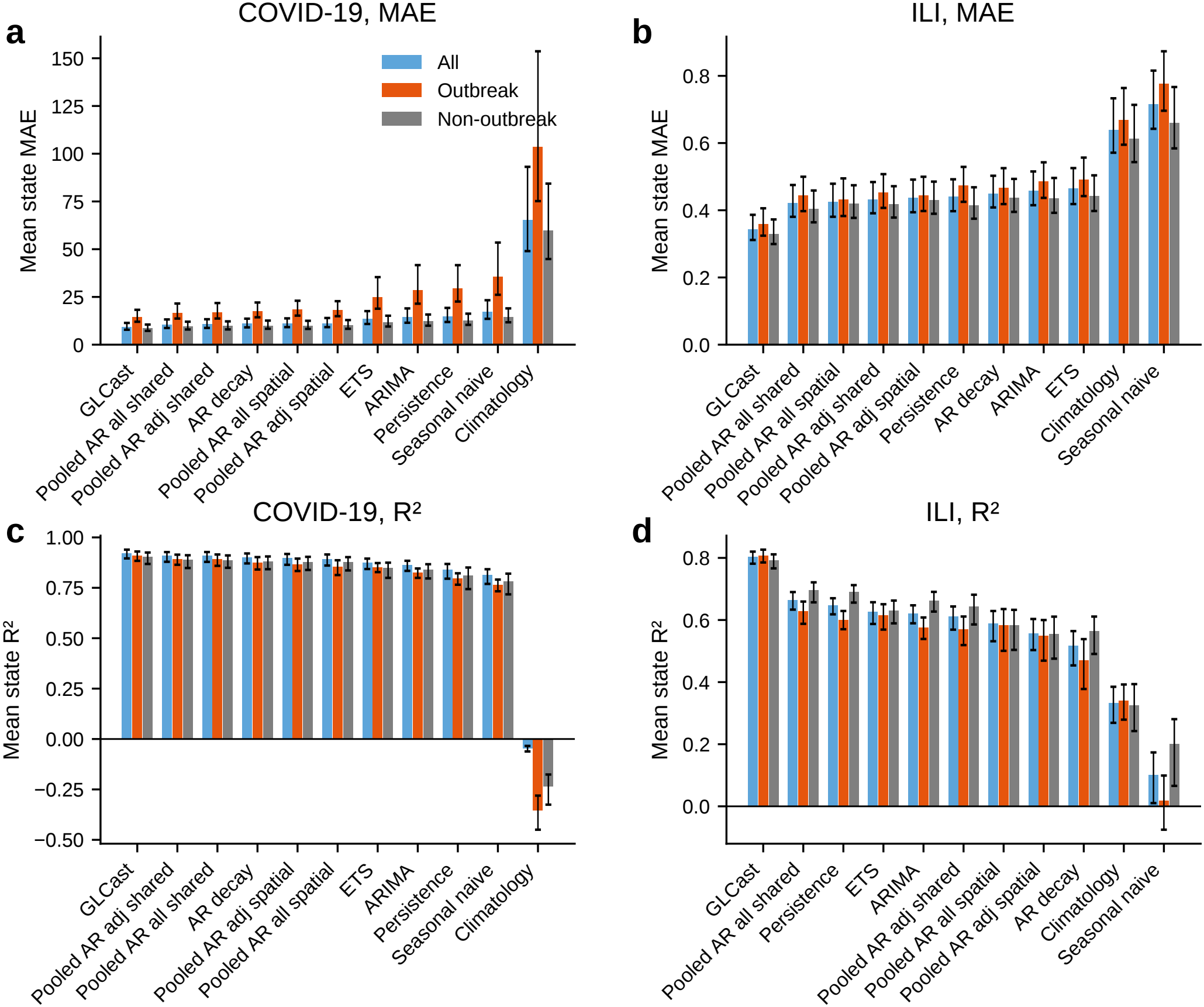
Alternate metrics of performance. Mean absolute error and *R*^2^ for all eleven methods, computed within each jurisdiction and averaged equally over jurisdictions, for COVID-19 and ILI over all scored times, inside outbreaks, and outside them. *R*^2^ uses each jurisdiction’s own stratum mean as its reference and may be negative. GLCast has the lowest mean absolute error and highest mean *R*^2^ in every disease–stratum combination. Error bars are descriptive 95% BCa intervals from 10,000 jurisdiction bootstrap resamples, conditional on the historical benchmark choice of specification.

**Fig. S10.**
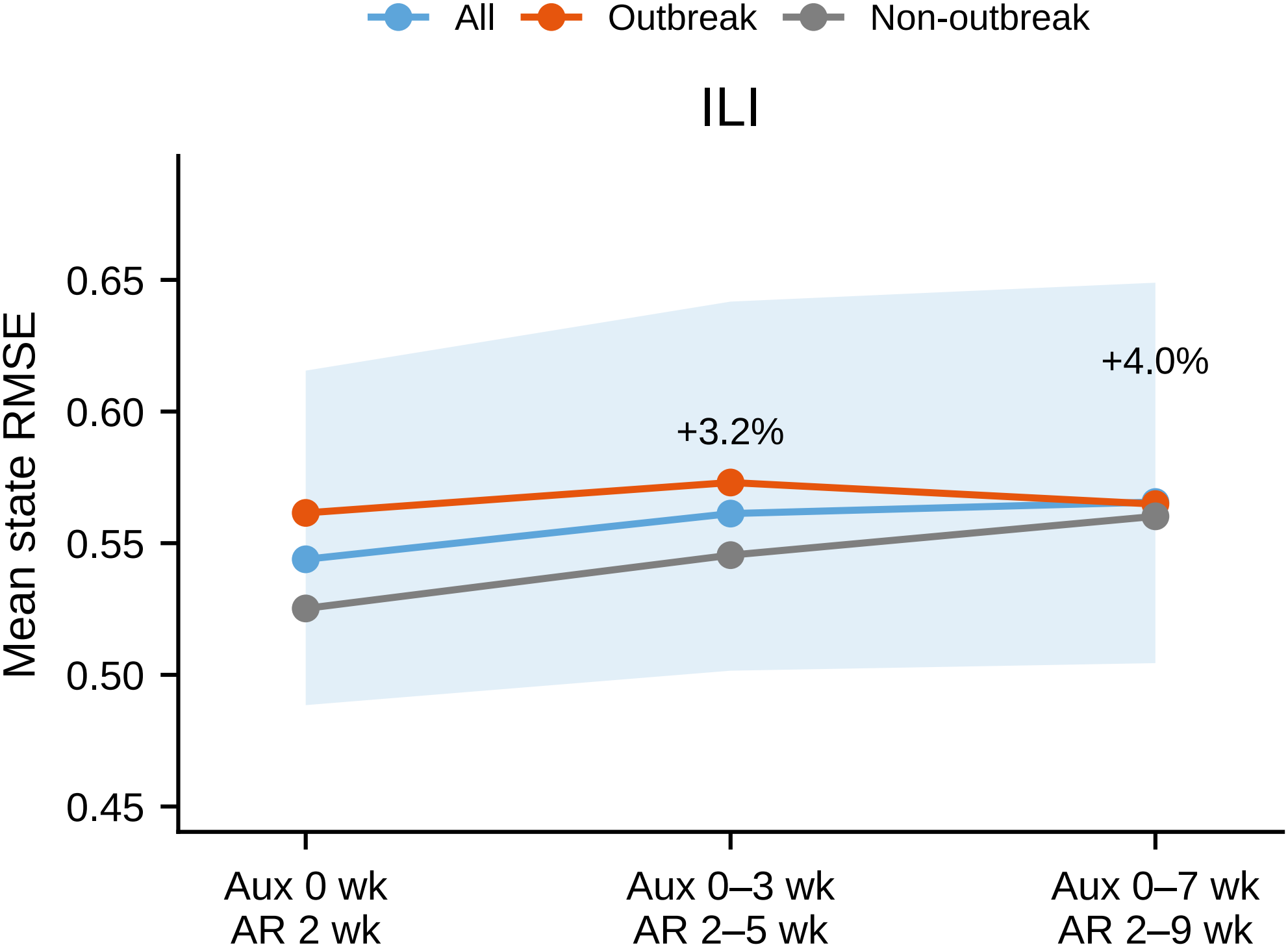
ILI accuracy under expanded lag windows. Mean state RMSE with the main design (auxiliary lag 0, target lag 2 weeks), a four-week history (auxiliary lags 0–3, target lags 2–5 weeks), and an eight-week history (auxiliary lags 0–7, target lags 2–9 weeks). Lines show all scored weeks, outbreak weeks, and non-outbreak weeks. The band is the descriptive 95% BCa interval for the all-period mean. Annotations give overall changes against the main design: +3.2% and +4.0%. For the longest history, changes are +6.7% outside outbreaks and +0.6% inside them. Both auxiliary and target lag sets change, and each design uses lag-matched pilot weights and alpha histories. These are sensitivities conditional on the historical benchmark; the main lag design is retained.

**Fig. S11.**
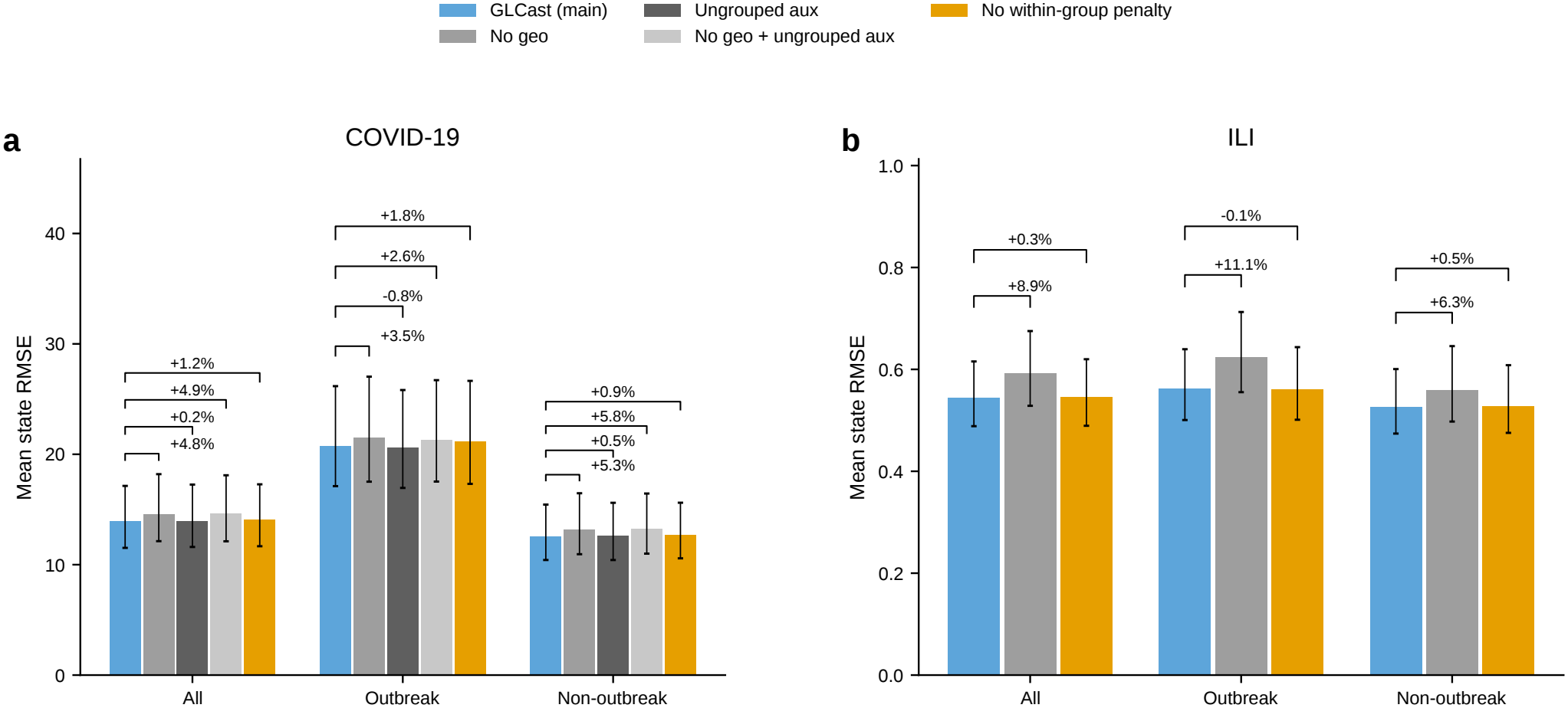
Geographic aggregation, auxiliary grouping, and coordinate penalties. Mean state RMSE over all scored times and within outbreak strata. Error bars are descriptive 95% BCa intervals; annotations give percentage change relative to the main model, with positive values indicating larger error. (**A**) COVID-19: removing geography changes overall error by +4.8%; splitting auxiliary coordinates into singleton groups while retaining the joint autoregressive group gives +0.17%; removing both gives +4.9%. Removing the within-group coordinate penalty gives +1.2%. (**B**) ILI: removing geography gives +8.9%, and removing the coordinate penalty gives +0.25%. ILI auxiliary groups are already singletons, so auxiliary ungrouping reproduces the corresponding grouped model. Penalty removal retains auxiliary and autoregressive lag sets and named adaptive group weights, while keeping the automatic alpha policy. It removes coordinate penalties from both feature types and follows the recorded group-only solver dispatch. All comparisons are descriptive, conditional on historical benchmark selection.

**Fig. S12.**
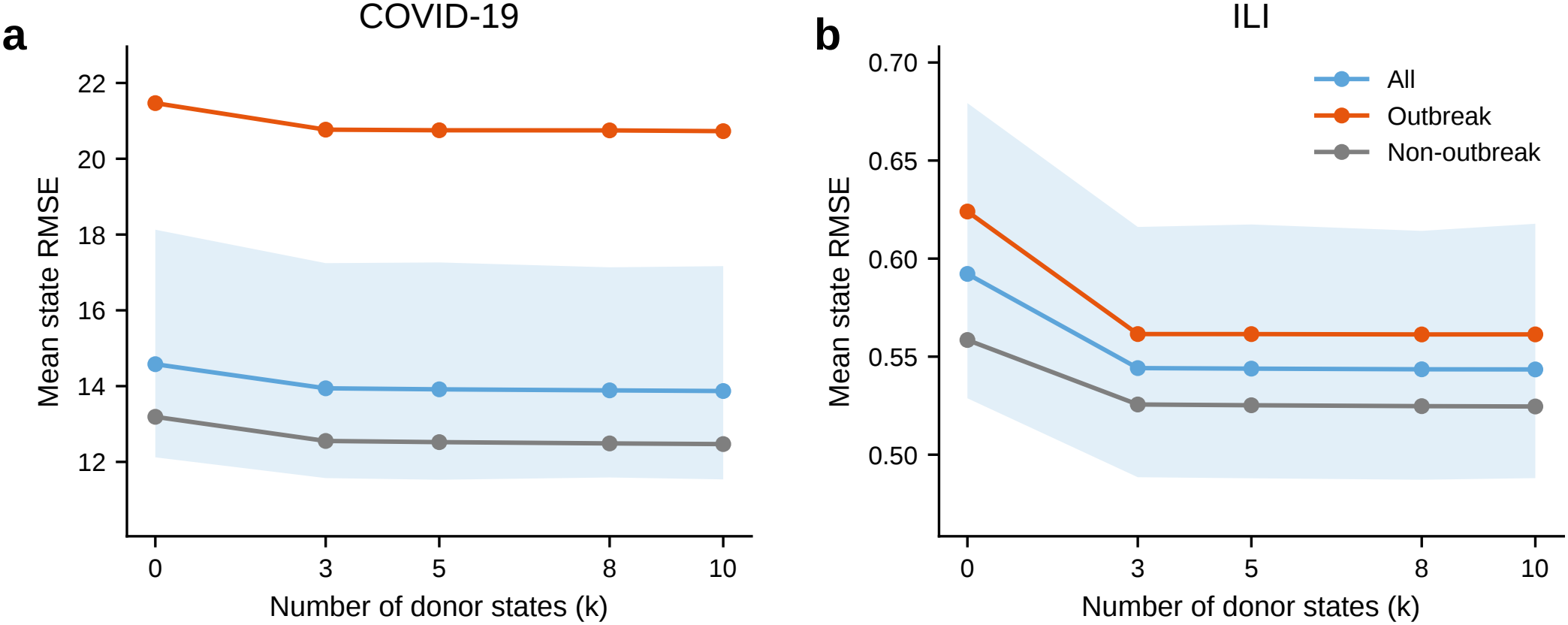
Sensitivity to the geographic donor cap. Mean state RMSE over all scored times, outbreak periods, and non-outbreak periods as the retained donor cap varies over *k* = 3, 5, 8, 10, with the similarity-screened library fixed at 20. The *k* = 0 reference is the raw unaggregated local prediction. The main configuration uses *k* = 5. Overall RMSE for COVID-19 is 14.5789 at *k* = 0, 13.9422 at *k* = 3, 13.9150 at *k* = 5, and 13.8701 at *k* = 10; corresponding ILI values are 0.5922, 0.5442, 0.5439, and 0.5435. All configurations use the complete original masks, including monthly fallback cells. Bands show descriptive 95% BCa intervals for all-period mean RMSE, conditional on the benchmark-selected specification. Larger donor caps give small additional overall gains beyond the main configuration.

**Fig. S13.**
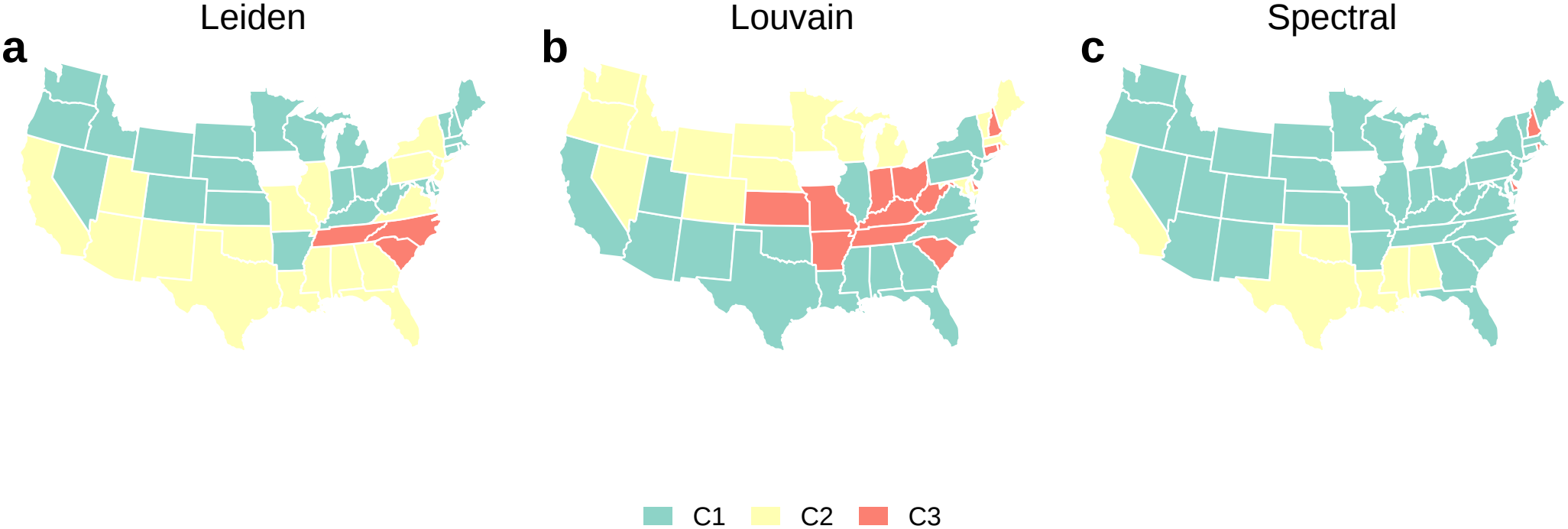
ILI network communities under alternative clustering algorithms. Communities of the time-averaged, symmetrized graph of normalized aggregation shares, with self-loops removed, detected by (**A**) Leiden (63), (**B**) Louvain (83), and (**C**) spectral clustering (84). Leiden and Louvain each return three communities; the spectral fit uses the Louvain community count. Modularity is *Q* = 0.192, 0.200, and 0.089, respectively. Assignments differ, and community colors are indexed separately within each partition. These maps summarize average display shares; a fixed partition is not imposed on individual nowcasts.

**Fig. S14.**
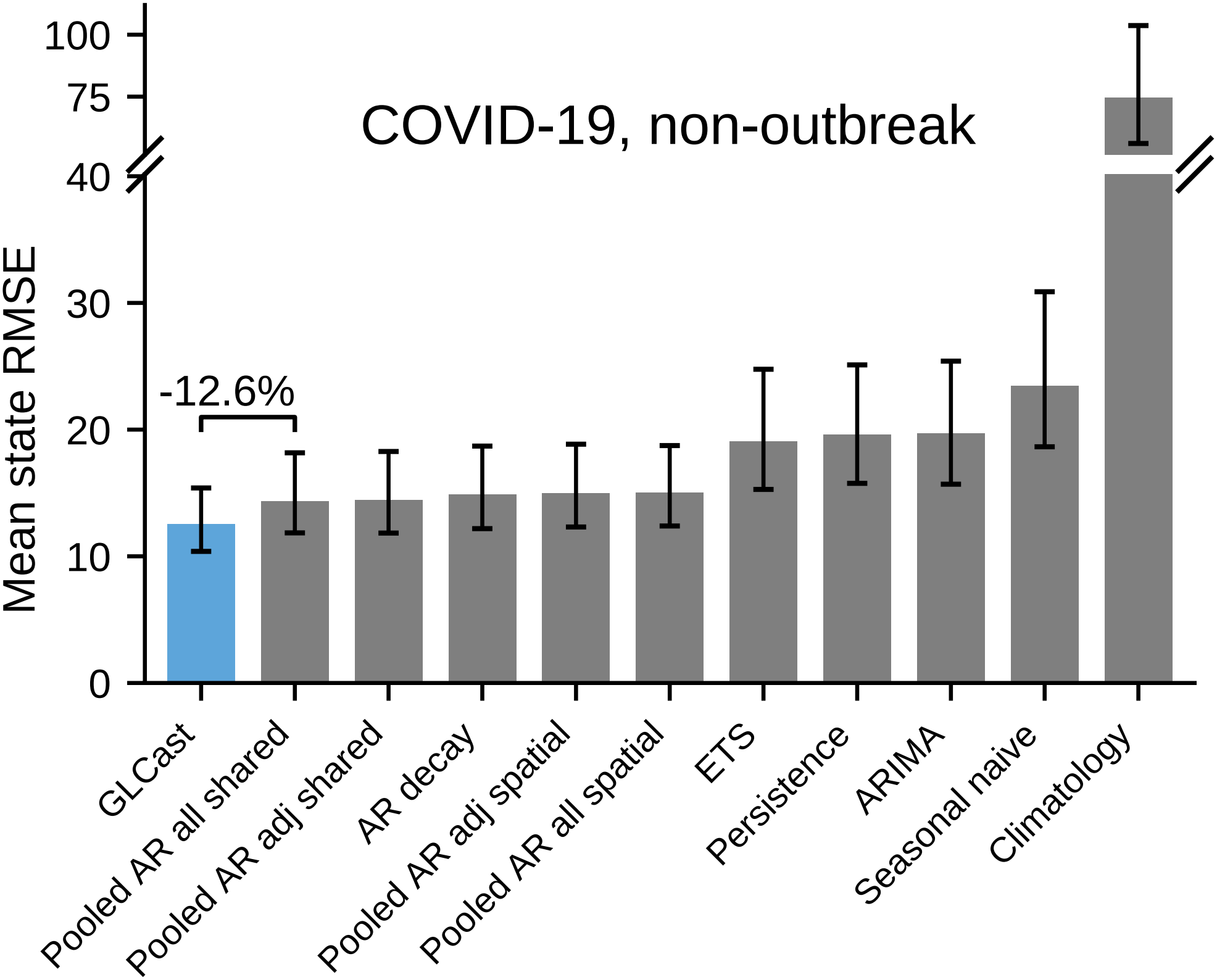
Non-outbreak COVID-19 performance of the full comparator roster. Equal-jurisdiction mean RMSE across 51 jurisdictions for GLCast and ten comparators over non-outbreak days; the axis is broken to accommodate Climatology. Error bars are descriptive 95% BCa intervals from 10,000 jurisdiction resamples, conditional on the benchmark-selected specification. The bracket compares GLCast with Pooled AR all shared: 12.52 versus 14.33 admissions per day, a 12.6% reduction (paired absolute difference 1.81; interval [1.32, 2.55]). The annotation expresses signed percentage change, with a negative value favoring GLCast.

**Fig. S15.**
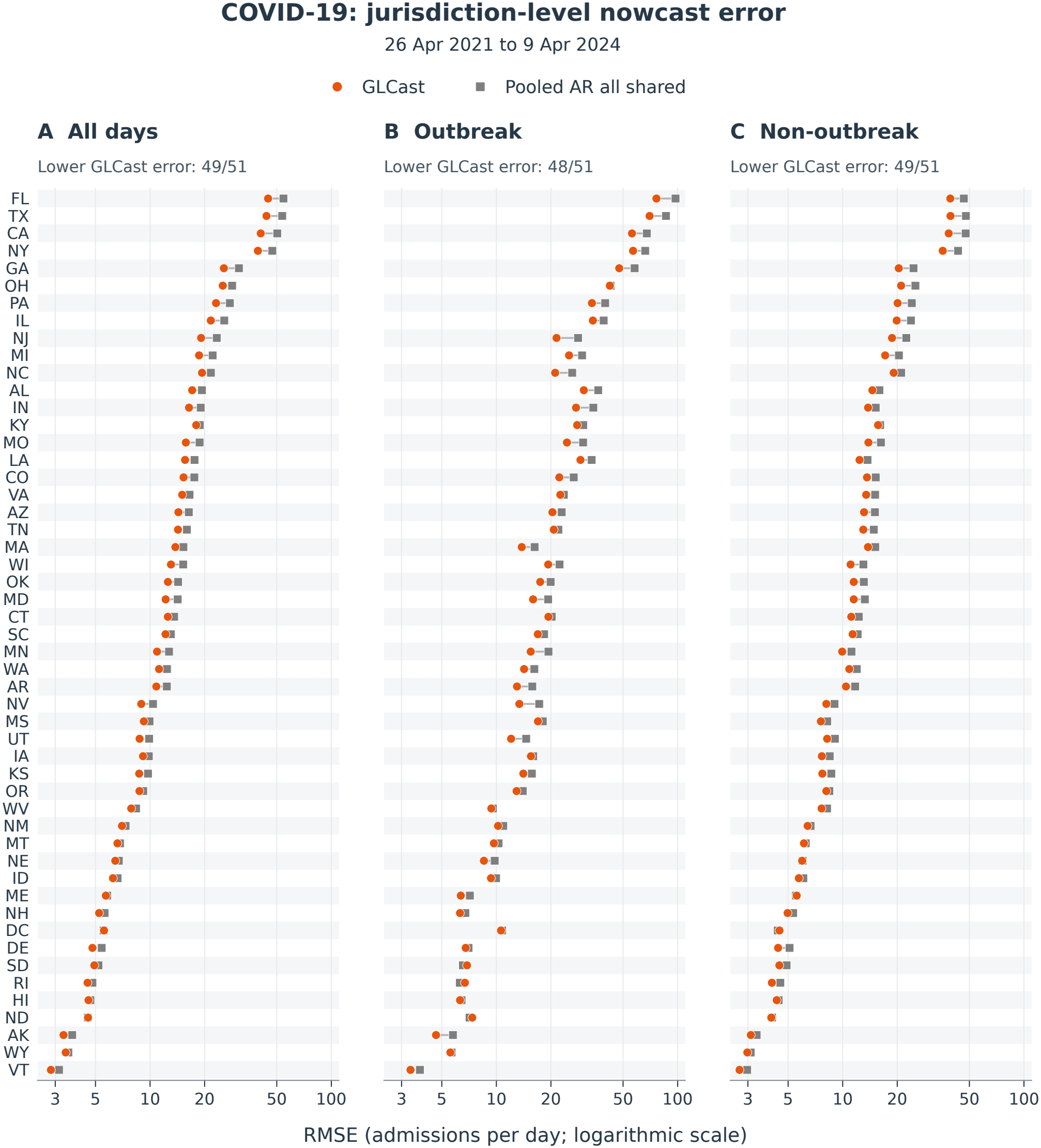
Jurisdiction-level COVID-19 admission prediction error. Connected orange circles and gray squares show GLCast and the fixed Pooled AR all shared comparator for all 50 states and the District of Columbia over (**A**) all scored days, (**B**) outbreak days, and (**C**) non-outbreak days. RMSE is measured in daily admission counts and shown on a logarithmic horizontal scale. Scoring retains the original 55,080 jurisdiction-days from April 26, 2021, through April 9, 2024, with predictions floored at zero. Jurisdictions are ordered by decreasing comparator RMSE over all days, with identical order across panels. GLCast has lower RMSE in 49, 48, and 49 jurisdictions, respectively. Both methods cover identical cells in every jurisdiction and stratum. The points are individual-jurisdiction error estimates.

**Fig. S16.**
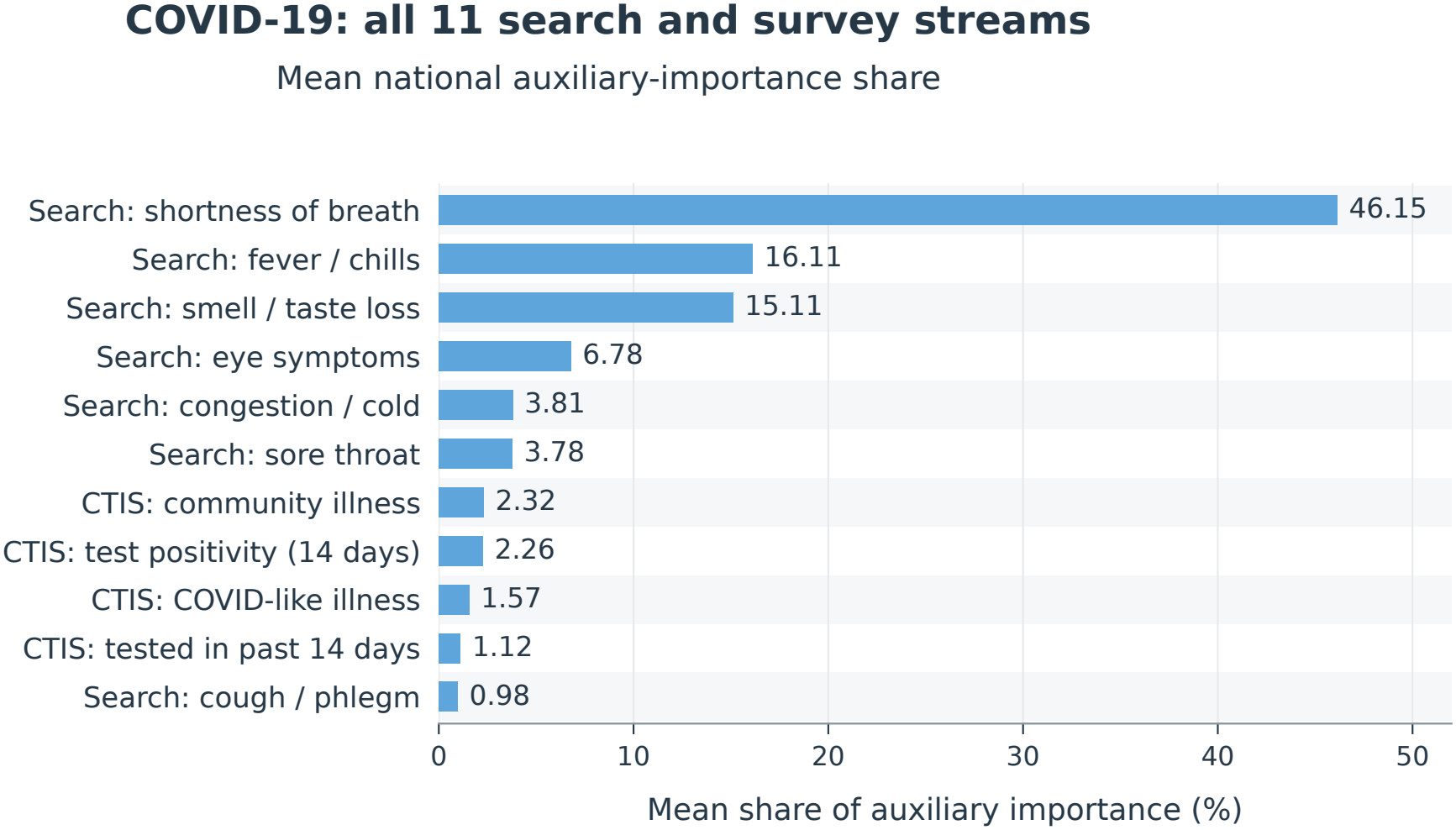
Mean auxiliary importance across all 11 search and survey streams. For each retained local-fitting origin, absolute raw coefficients multiplied by the corresponding rolling 360-day predictor standard deviations are summed across states and each stream’s lags, then normalized across auxiliary streams. Bars average the national shares equally across all 156 retained origins, April 2, 2021, through March 22, 2024; all have nonzero auxiliary mass. Search denotes Google symptom sets, and CTIS denotes survey measures. The target-history group is excluded, so these auxiliary-only shares differ from the reported fractions of total coefficient mass. The summaries describe local fits rather than attribution of the final geographic ensemble and retain the supplied histories, including constant filling after CTIS collection ceased. Retained diagnostic origins need not serve scored cells. Values are rounded to two decimal places; no uncertainty intervals are estimated.

**Fig. S17.**
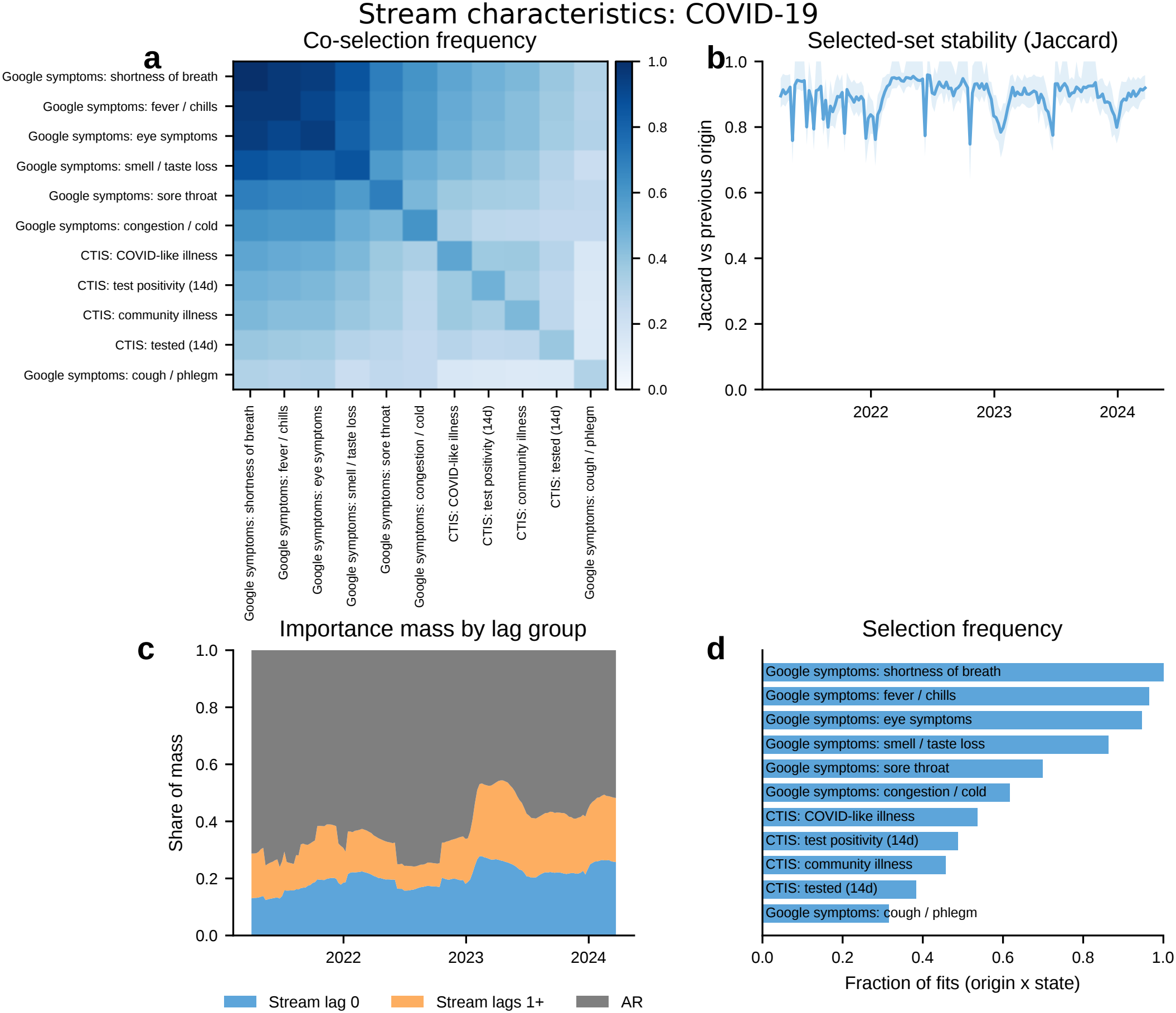
How GLCast uses the 11 search and survey streams over time (COVID-19). Diagnostics over 7,956 retained state-origin fits (156 origins *×* 51 jurisdictions), with panels defined in *SI Appendix*, Fig. S2. The mean successive-set Jaccard similarity is 0.893; its plotted band is the across-state interquartile range. Mean coefficient-mass shares are 62.8% autoregressive, 20.3% contemporaneous auxiliary, and 16.9% older auxiliary. Every retained fit selects an auxiliary stream; the median fit contains seven streams and 19 total features. Shortness of breath is selected in 100% of fits, fever/chills in 96.5%, and eye symptoms in 94.7%. Smell/taste loss is available in 45 jurisdictions; selection frequencies use all retained fits. These diagnostics describe local models at the automatically selected penalties and include origins outside the final scoring mask.

**Fig. S18.**
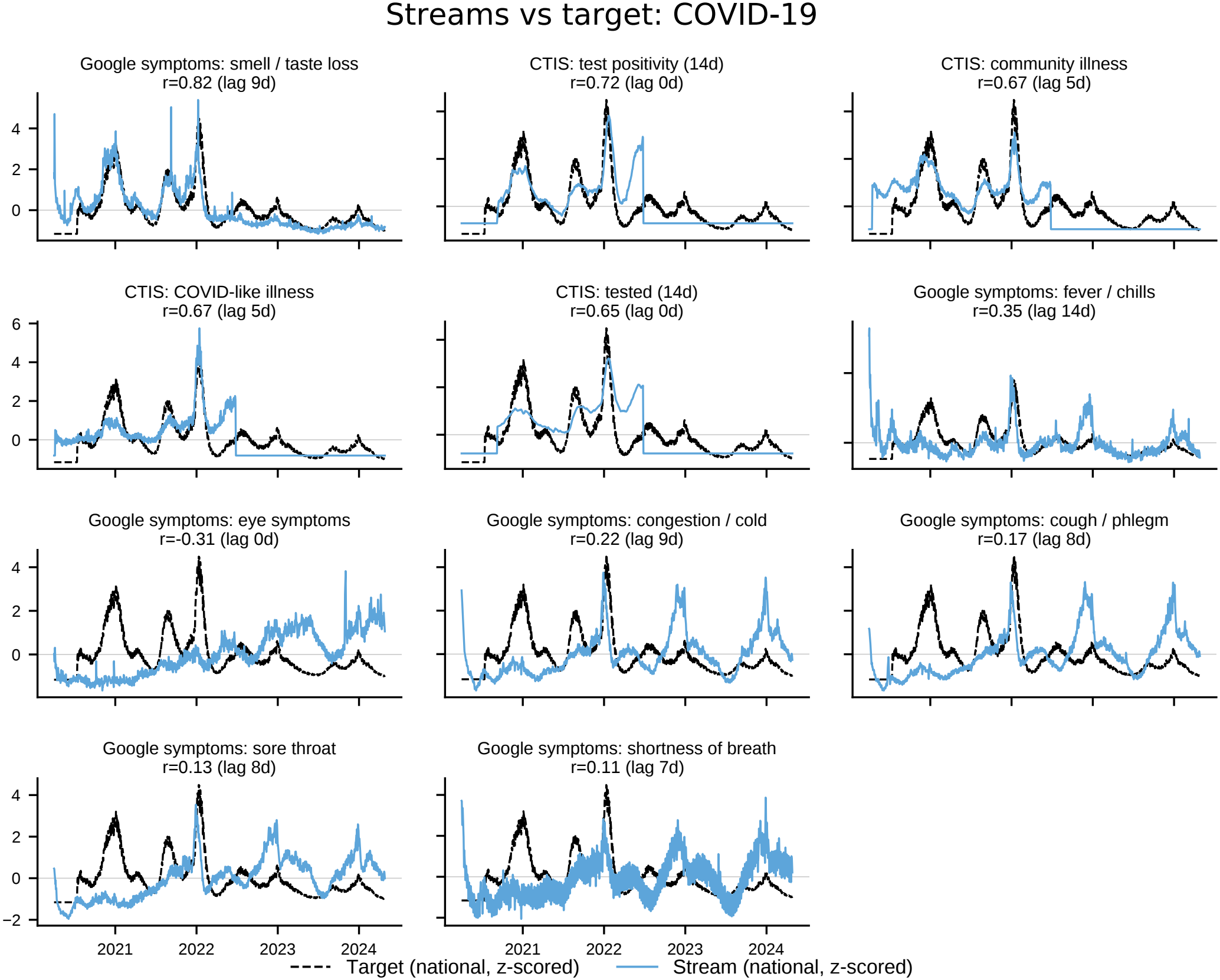
Search and survey streams against the COVID-19 target. Each mini-panel overlays a nationally aggregated auxiliary stream (blue) on national daily admissions (black dashed), both *z*-scored over the full input period, April 2020–April 2024. The national target is the unweighted mean across the 51 evaluated jurisdictions. Curves are unshifted; panels are ordered by maximum absolute Pearson correlation over lags of 0–14 days, and titles report the maximizing correlation and lag (positive lag: the stream leads the target). The strongest alignments are smell/taste loss (*r* = 0.82, nine-day lead), CTIS test positivity (0.72, contemporaneous), and CTIS community illness (0.67, five-day lead). Flat CTIS plateaus after mid-2022 are constant-filled inputs following survey cessation. These are descriptive input summaries, not measures of held-out prediction accuracy.

**Fig. S19.**
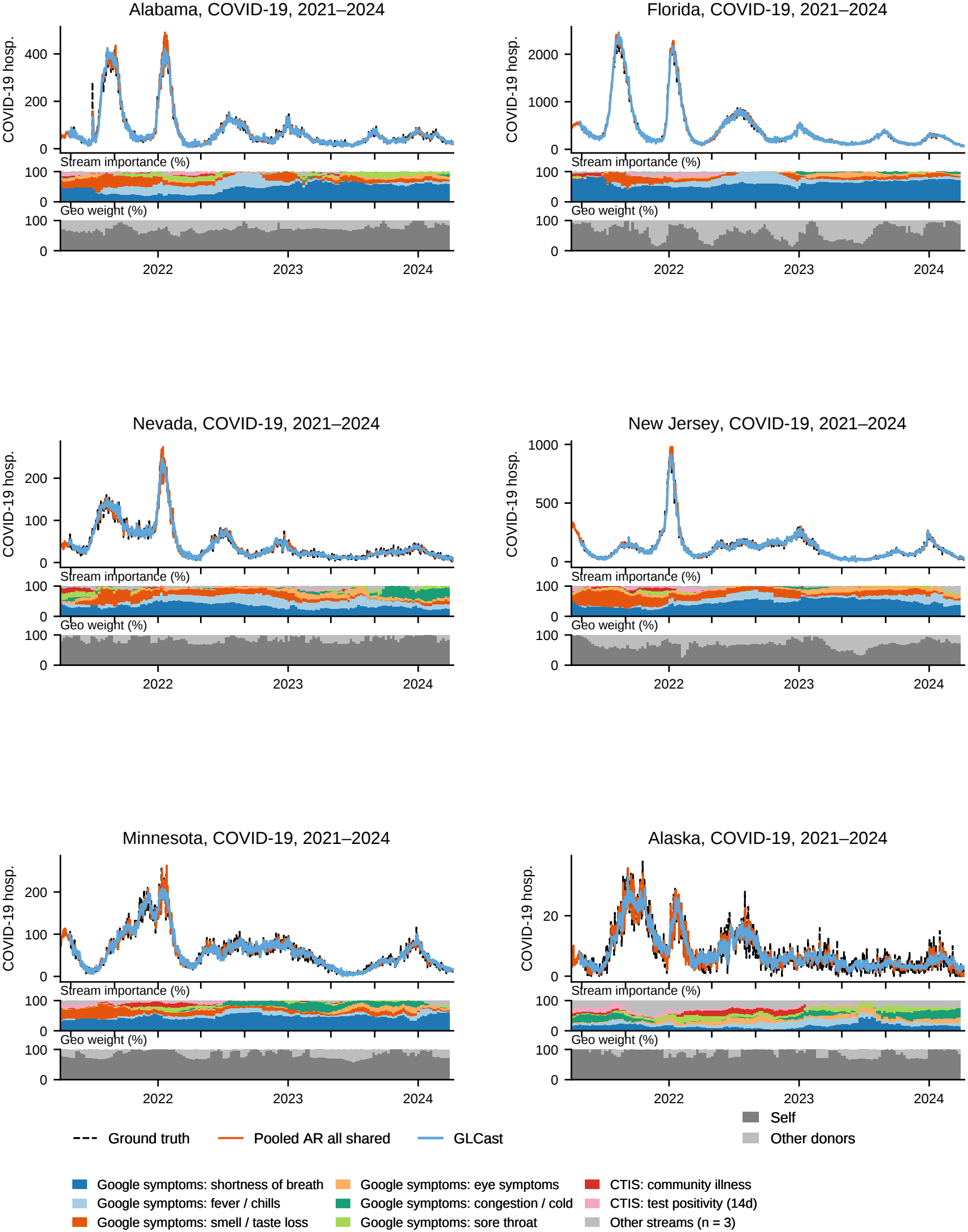
Six COVID-19 states: nowcasts and fitted information shares. Alabama, Florida, Nevada, New Jersey, Minnesota, and Alaska are retained illustrative examples, constructed as in *SI Appendix*, Fig. S1. The upper trace shows observed daily admissions against GLCast and the fixed Pooled AR all shared comparator, with predictions floored at zero. The middle strip shows state-specific auxiliary-importance shares for the same nationally ranked top eight streams in every cell, with three remaining streams pooled. The lower strip shows normalized self-versus-donor shares; nowcasts use raw blended weights. The traces and local-fit diagnostics extend beyond the final scoring mask.

**Fig. S20.**
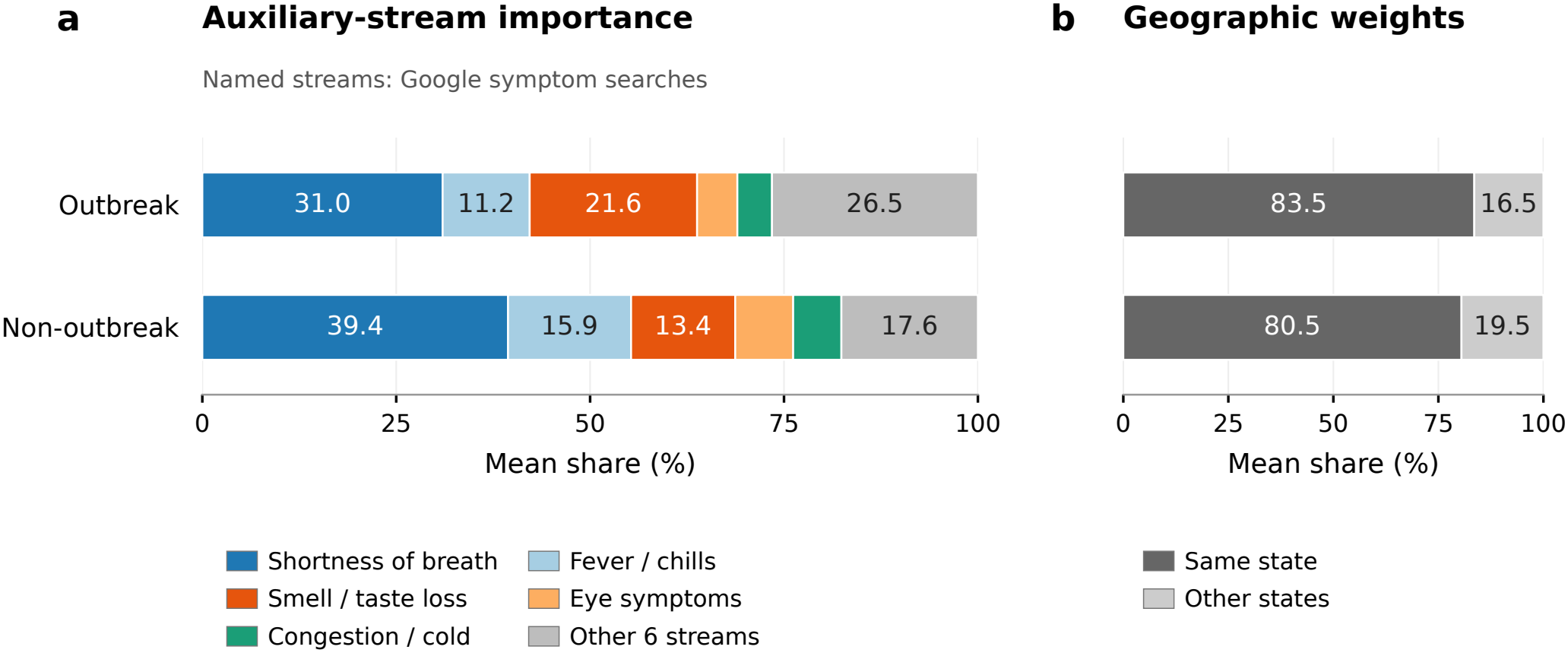
Stream and geographic importance during and outside classic COVID-19 outbreaks. (**A**) Mean auxiliary-stream shares for the five leading streams, selected once from the all-period summary, and the remaining six streams. Named symptom streams are Google searches. Local shares use absolute coefficient magnitude times the predictor’s rolling training-window standard deviation, summed over lags and normalized across auxiliary streams. Local shares are averaged equally across states within each scored day and phase, then equally across days. These are averages of local compositions, rather than pooled raw coefficient masses. (**B**) Same-state and other-state shares of the serving models’ nonnegative geographic weights, normalized within each model and averaged in the same order. Both panels use 54,519 scored state-days across 51 jurisdictions: 7,326 outbreak and 47,193 non-outbreak cells. The 561 fallback cells on March 30–April 9, 2024 are excluded from both panels because their serving fits lack archived local-importance records. All included auxiliary compositions are defined. Archived outbreak labels are assigned to scored target dates. Bar lengths use unrounded descriptive means; no confidence intervals are estimated.

**Fig. S21.**
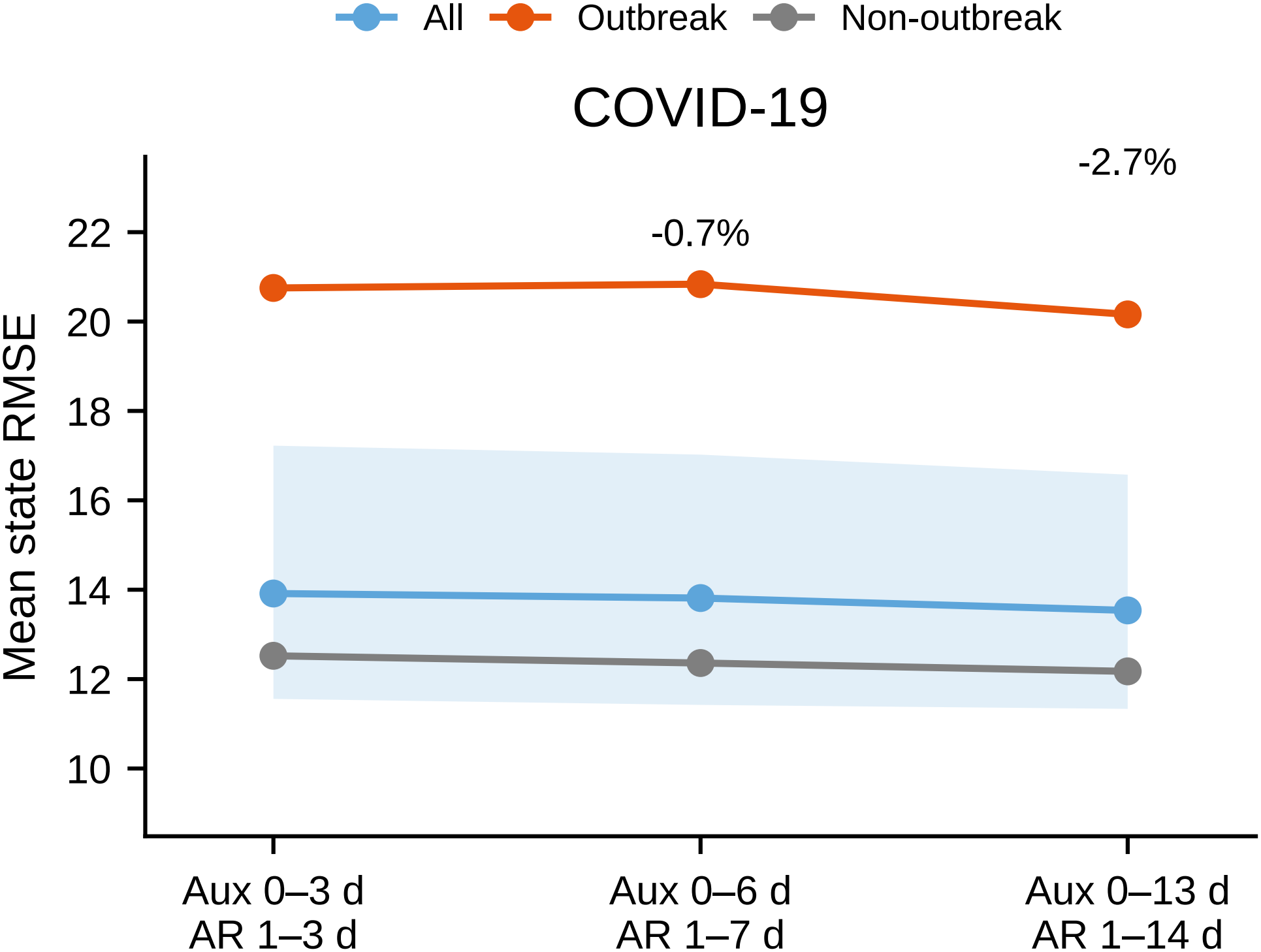
COVID-19 accuracy under expanded lag windows. Mean state RMSE with the main design (auxiliary lags 0–3, target lags 1–3 days), a one-week history (auxiliary lags 0–6, target lags 1–7 days), and a two-week history (auxiliary lags 0–13, target lags 1–14 days), over all days, outbreak days, and non-outbreak days. The band is the descriptive 95% BCa interval for the all-period mean. Overall changes against the main design are *−*0.7% and *−*2.7%. The longest history has 2.9% lower outbreak error and 2.8% lower non-outbreak error. Both lag sets change, with their own lag-matched pilot weights and alpha histories. These are sensitivities conditional on the historical benchmark; the main configuration remains fixed.

**Fig. S22.**
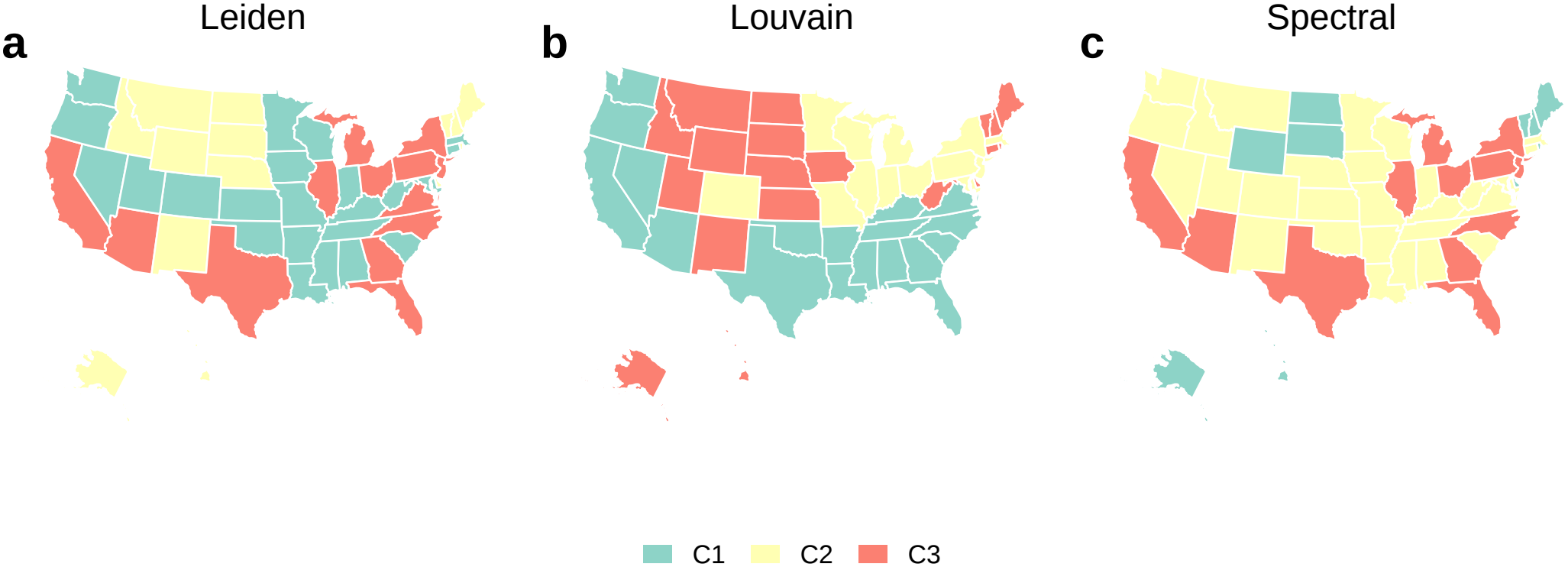
COVID-19 network communities under alternative clustering algorithms. Communities of the time-averaged, symmetrized graph of normalized aggregation shares, with self-loops removed, detected by (**A**) Leiden (63), (**B**) Louvain (83), and (**C**) spectral clustering (84). Leiden and Louvain each return three communities; the spectral fit uses the Louvain community count. Modularity is *Q* = 0.329, 0.316, and 0.293, respectively. Assignments differ, and community colors are indexed separately within each partition. These are descriptive summaries of the average display-share network.

**Fig. S23.**
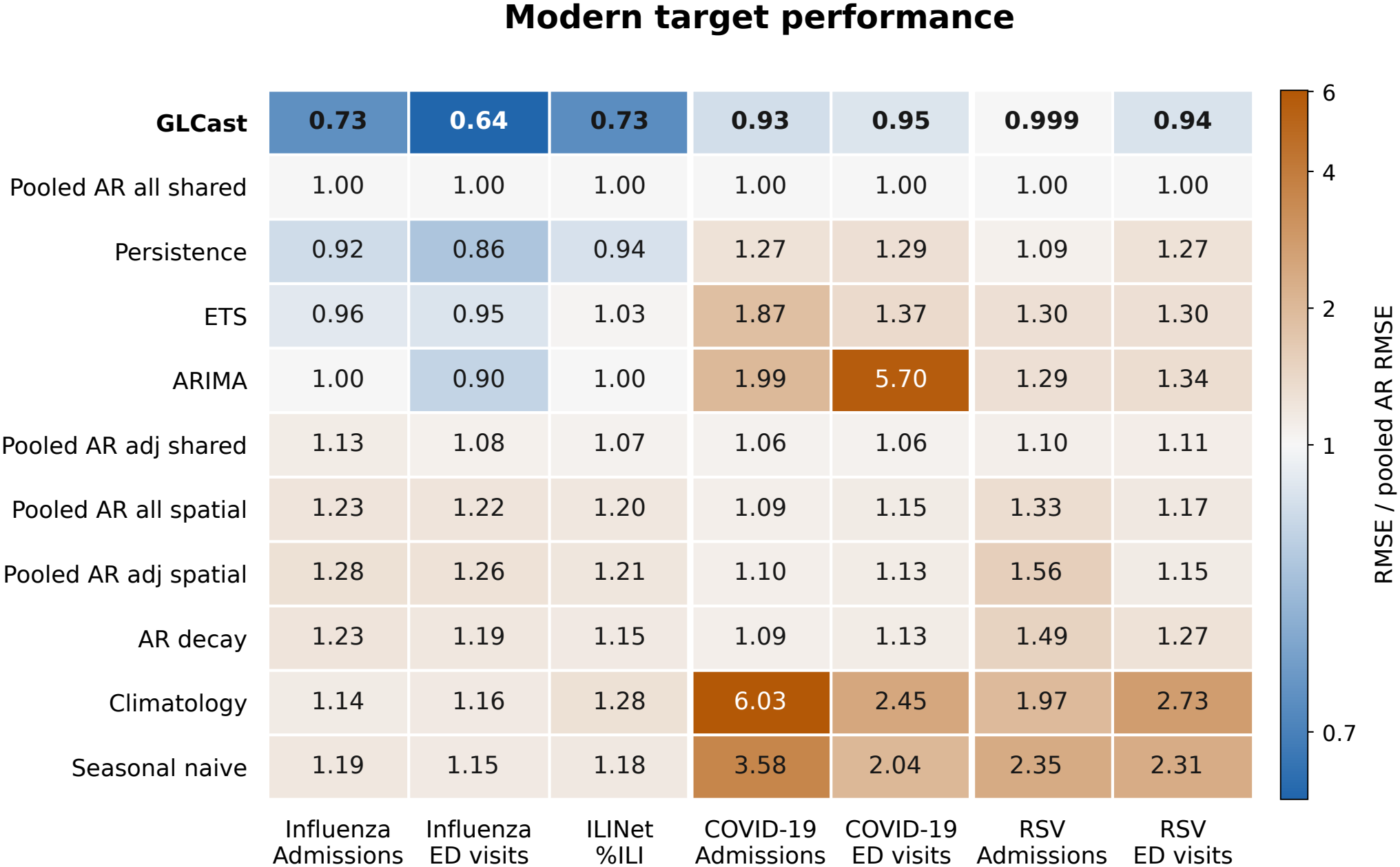
Full comparator roster across all seven modern targets. Each cell gives equal-jurisdiction mean RMSE divided by the mean RMSE of the fixed Pooled AR all shared reference for that target. One denotes equal error, and smaller values are better. Color is based on the logarithm of this ratio, centered at one; raw errors with different units are not pooled. GLCast has the lowest reported point error on all seven targets, with only a 0.1% reduction for RSV admissions; that ratio is shown to three decimal places to retain the small point difference. The ranks do not imply identical evaluation cells for every method. The overall contrasts in Fig. 4*a* use each target’s observed best overall baseline, with complete, identical cells for GLCast and that selected reference. Influenza admissions and ED visits are pathogen-specific outcomes; ILINet measures syndromic ILI. These are descriptive historical ranks conditional on the common specification selected using these benchmarks.

**Fig. S24.**
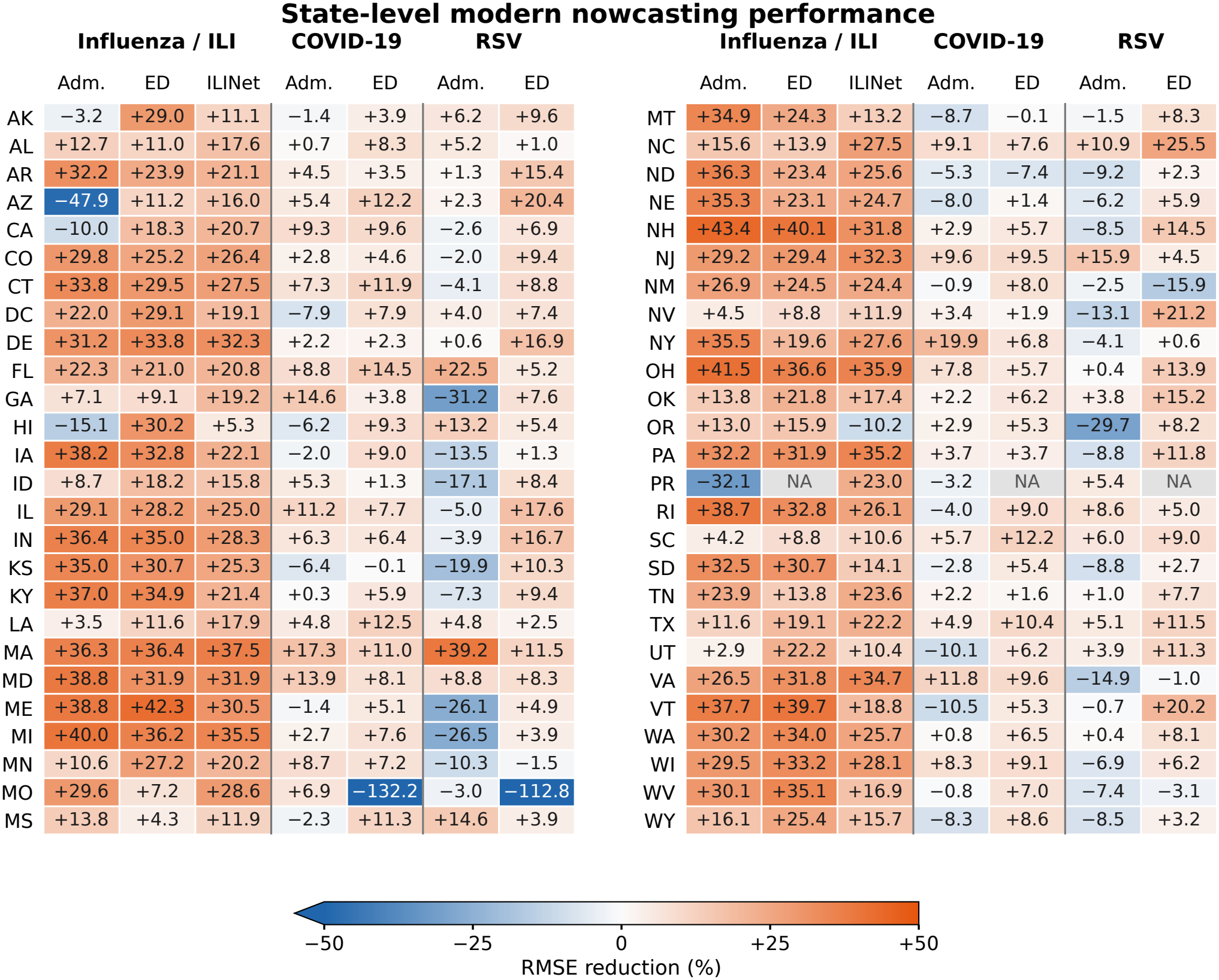
Modern RMSE comparisons in every evaluated jurisdiction. Each cell gives 100(1 *− e*_GLCast_*/e*_reference_), with RMSE calculated on the same retained weeks within the jurisdiction after flooring predictions at zero. The reference is the target-wide best overall baseline, held fixed across jurisdictions: Persistence for the three influenza/ILI targets and Pooled AR all shared for the four COVID-19/RSV targets. Positive orange values favor GLCast; negative blue values indicate higher error. All 361 evaluated target–jurisdiction combinations are shown. The three gray NA cells are Puerto Rico ED outcomes, which are not evaluated. Rows follow postal-code order in two blocks; Adm. denotes hospital admissions, DC the District of Columbia, and PR Puerto Rico. Cell values are rounded to one decimal. The common linear color scale saturates below *−*50%; the two more extreme Missouri values remain printed. Improvements occur in 47/52 influenza-admission jurisdictions, 51/51 influenza ED, 51/52 ILINet, 34/52 COVID-19 admissions, 47/51 COVID-19 ED, 23/52 RSV admissions, and 46/51 RSV ED. These are descriptive state-level comparisons conditional on the saved fits and the target-wide comparator choice; no state-specific intervals or tests are added.

**Fig. S25.**
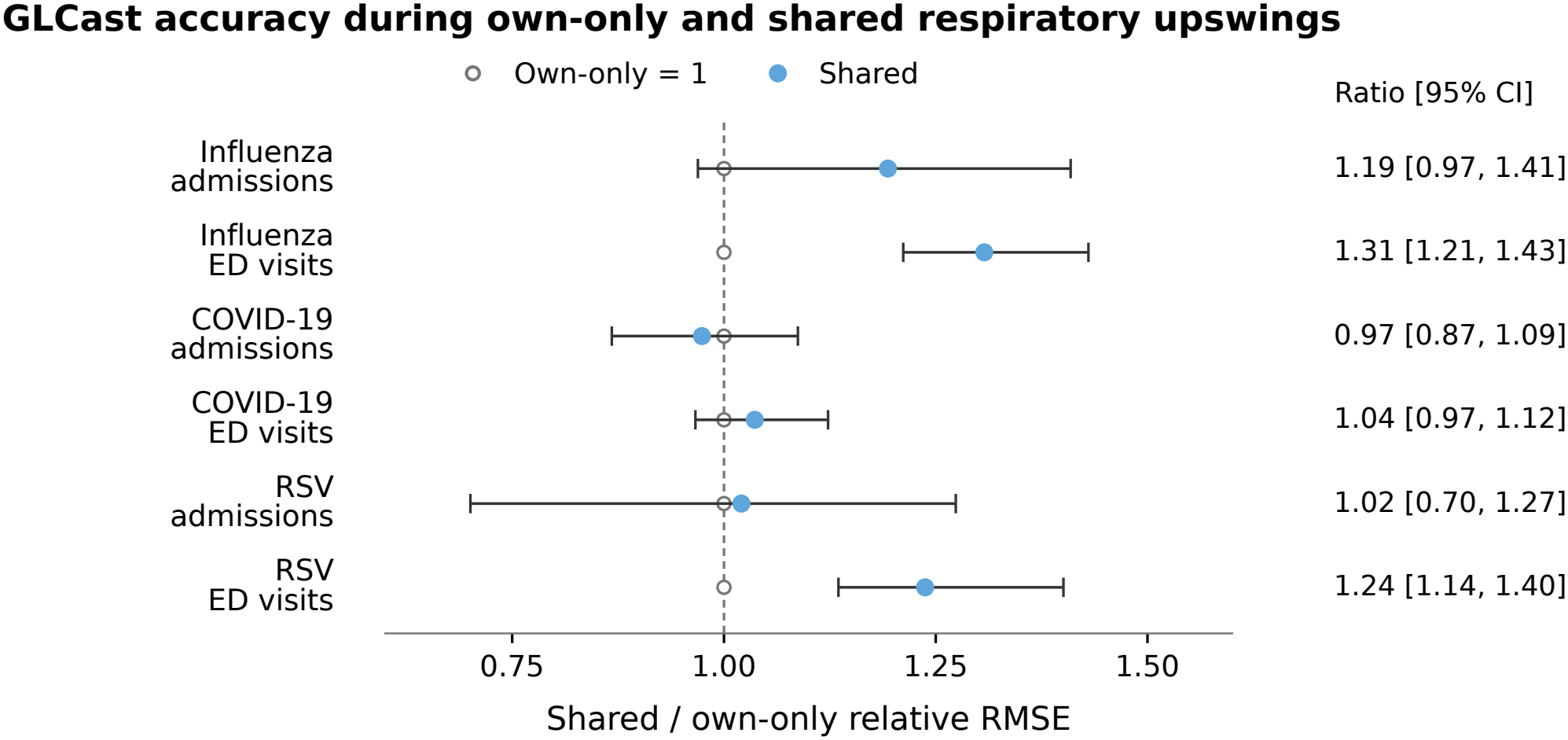
GLCast accuracy during shared and own-only respiratory upswings. Points show the ratio of mean amplitude-relative error during shared upswings to that during own-only upswings for the six disease-specific admission and ED targets. Within each jurisdiction and phase, RMSE is divided by the observed outcome RMS; both phase means then weight the same eligible jurisdictions equally. A ratio of one indicates equal mean relative error, and values above one indicate higher relative error during shared upswings. Whiskers are descriptive 95% paired-jurisdiction BCa intervals from 10,000 resamples. Each jurisdiction has at least four scored weeks in each phase and complete observations for all three viruses in the corresponding admission or ED family. This is a within-GLCast comparison and uses no baseline method. Four intervals include one; influenza and RSV ED intervals lie above one. An interval containing one does not establish equivalent accuracy.

**Fig. S26.**
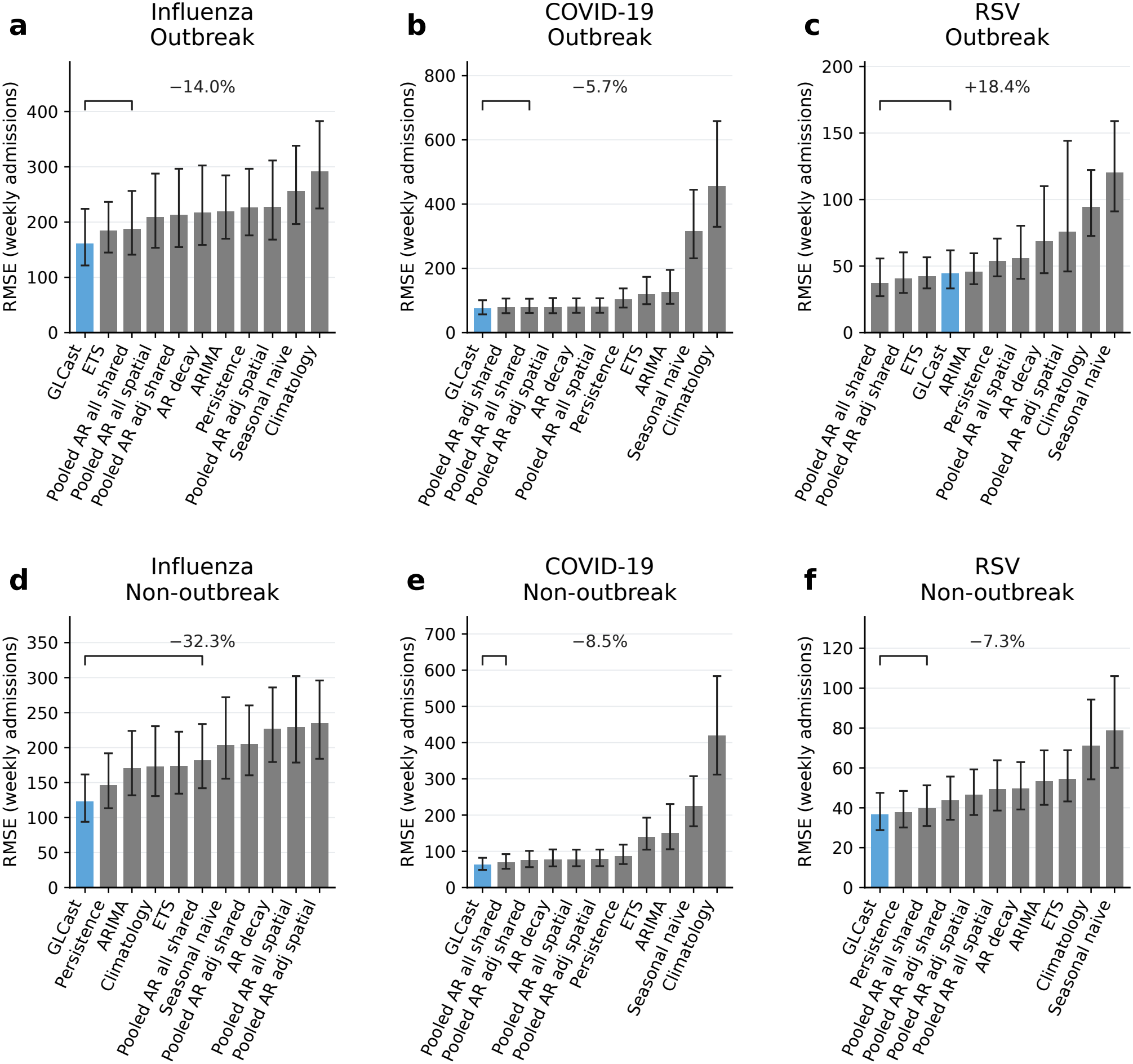
Modern NHSN admission nowcasting, stratified by outbreak status. Equal-jurisdiction mean RMSE for GLCast and ten comparators for influenza, COVID-19, and RSV weekly admission counts. (**A**–**C**) Retrospectively detected outbreak weeks. (**D**–**F**) Non-outbreak weeks. Error bars are descriptive 95% BCa intervals from 10,000 jurisdiction bootstrap resamples, conditional on the saved predictions and benchmark-selected recipe. Brackets compare GLCast with the fixed Pooled AR all shared comparator, which need not be the lowest-error baseline. Annotations show percentage change in RMSE; negative values favor GLCast. Outbreak changes are *−*14.0%, *−*5.7%, and +18.4%; non-outbreak changes are *−*32.3%, *−*8.5%, and *−*7.3%, respectively. Each panel includes 52 jurisdictions. GLCast and the fixed comparator cover all retained cells. These intervals do not account for historical recipe selection and are not confirmatory tests.

**Fig. S27.**
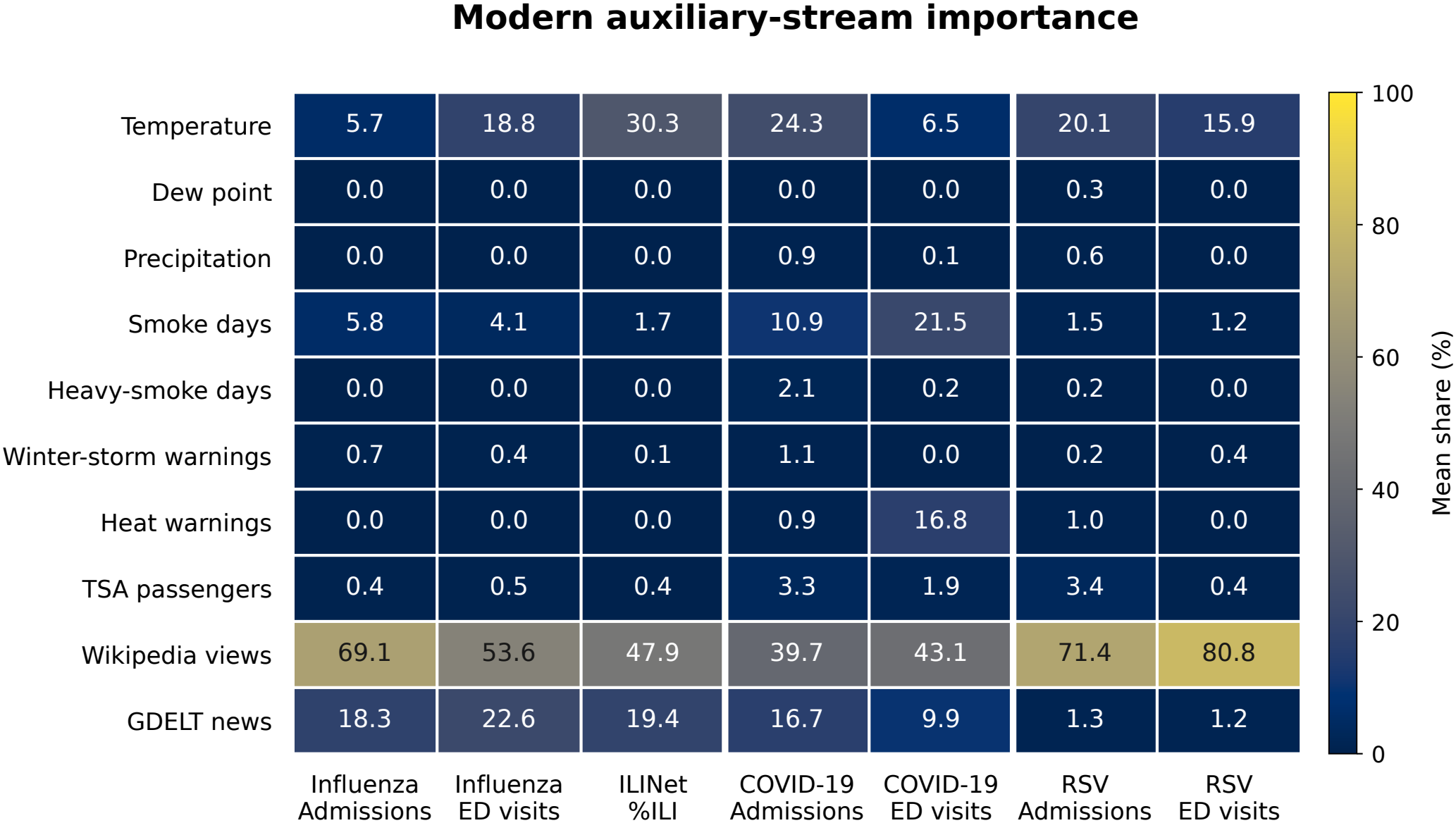
Local auxiliary-stream importance across seven modern targets. Cells show the mean percentage share of auxiliary coefficient importance at served origins. Importance is absolute raw local coefficient magnitude times rolling 55-week predictor standard deviation, summed across jurisdictions and normalized across auxiliary streams within each origin before averaging. The target-history group is excluded, and disease-specific Wikipedia and GDELT columns are mapped to common concepts for display. Columns sum to 100%. Twelve zero-total origins for RSV admissions are excluded, leaving 120 contributing origins; all other targets have 180 contributing origins. Wikipedia and news together account for 53.0–87.4% of mean auxiliary importance. Values are rounded for display, so a displayed zero need not imply an identically absent contribution. These are descriptive local-fit diagnostics, not causal effects or attribution of the final geographic ensemble.

**Fig. S28.**
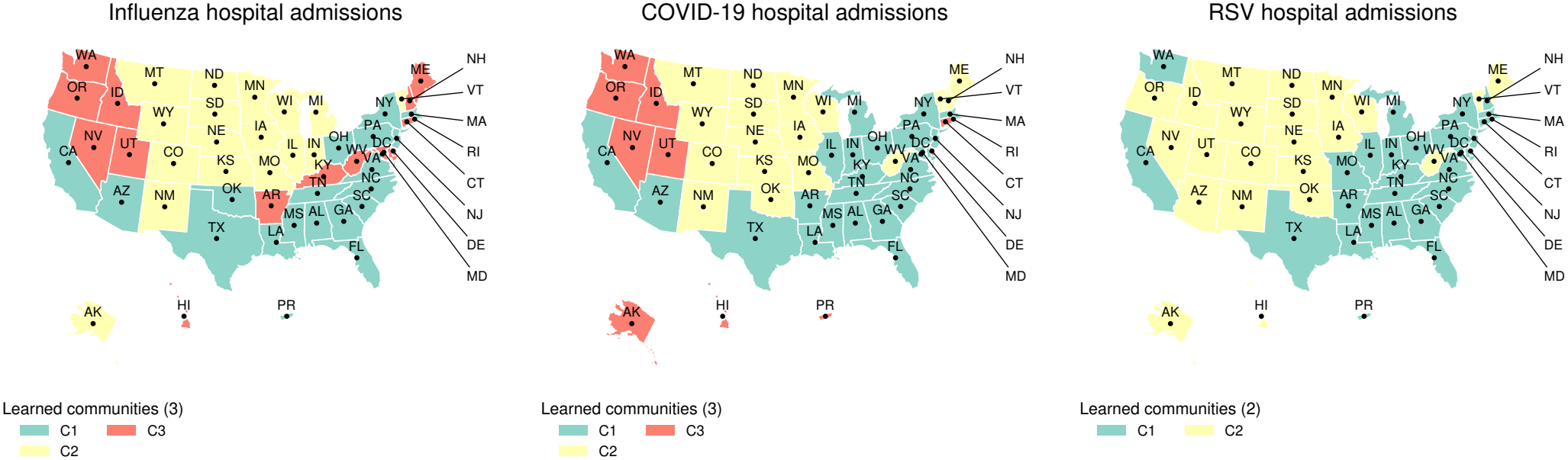
Learned aggregation communities for the modern NHSN admission targets. Community membership for influenza (left), COVID-19 (middle), and RSV (right), derived by Leiden clustering of each time-averaged, symmetrized graph of normalized aggregation shares after removing self-loops. The displayed partitions contain three, three, and two communities, respectively. Colors and labels are assigned separately within each disease and do not denote corresponding communities across diseases. These maps summarize display shares; predictions use the original raw blended weights. The graphs describe historical model borrowing and do not identify transmission pathways or provide independent validation.

